# Representation learning based on proteomic profiles uncovers key cell types and biological processes contributing to the plasma proteome

**DOI:** 10.1101/2024.12.16.24319106

**Authors:** Jiali Zhuang, Erin N. Smith, Dorothée Diogo

## Abstract

The plasma proteome is a potential source of information on health status and physiological condition and holds great potential as candidate biomarkers for diagnosis, prognosis, intervention response monitoring, and patient stratification. As proteins in the plasma can be derived from numerous cellular and tissue sources, and their levels influenced by diverse mechanisms, a comprehensive assessment of patterns of protein variation could provide insight into mechanisms driving health and disease. By applying neural network-based representation learning and unsupervised clustering to the plasma proteomic profiles of 51,180 participants in the UK biobank, we identified 36 protein modules representing major cell types and biological processes present in the plasma proteome. We discovered that the overall abundances of proteins belonging to certain modules are associated with disease status and genetic variants. Those associations reflect complex and multi-faceted mechanisms that regulate protein abundances in circulation. An investigation into the protein modules associated with disease variants uncovered both known disease biology and novel findings that may translate into testable hypotheses. Our approach generates biologically relevant groupings of plasma proteins that can be deployed to inform the design of more predictive biomarker panels and shed new light on the effects of disease-associated genetic variants.

## Introduction

Plasma protein concentrations have long been used as biomarkers for diagnosis and patient monitoring. Recent technological advances now permit accurate measurement of thousands of proteins simultaneously and offer a unique opportunity to comprehensively study the plasma proteome. UK Biobank Pharma Proteomics Project (PPP) employed the Olink proximity extension assay to profile the plasma proteomes of 54,219 UKB participants and identified over 12,000 significant associations between genetic variants and plasma protein levels [1]. The extensive genetic, phenotypic and clinical information available for the same participants allows for the discovery of genetic factors affecting plasma protein abundances as well as biomarkers for diagnosis and disease progression monitoring [2–4]. The relative contribution of a given cell type or tissue to the plasma proteome depends on multiple factors including cell counts, secretory activity level, metabolic rate, vasculature density and local endothelial permeability.

The identification of the cell types and biological processes well represented in the circulating proteome informs the selection of disease areas that are most amenable to non-invasive monitoring using blood protein biomarkers. Ascertaining the tissue/cell type origin of a plasma protein based solely on expression enrichment can be challenging, considering the limited number of highly cell type specific proteins and the wide range of relative contribution levels across cell types. Proteins that originate from the same cell type or participate in the same biological process are more likely to share similar expression patterns across participants in the cohort. A data driven approach that starts with identifying co-expressed protein modules could therefore better unravel the cell types and biological processes represented in the plasma proteome and assign individual proteins to their source. Linear decomposition algorithms such as principal component analysis (PCA) and non-negative matrix factorization (NMF) have been applied to high-dimensional omics data for deconvolution purposes [5, 6]. However, the underlying biological factors do not always follow linear relationships. A variational autoencoder (VAE) model includes an encoder and a decoder and learns a lower dimensional representation of the input data while minimizing information loss incurred by the dimension reduction. The encoder and decoder of VAEs are implemented with neural network architecture and are therefore able to model non-linear relationships. Here we present a computational framework that learns a low dimensional representation of proteins based on the plasma proteomic profiles of 51,180 UK Biobank participants with VAE and derives biologically relevant protein modules from unsupervised clustering in the latent space. We showed that most of those modules represent distinct cell types or biological processes and discovered associations with disease status and genetic variants. We demonstrated that dysregulation of protein modules reflects known biology of diseases and could provide additional insight into key aspects of complex diseases.

## Results

### Dimension reduction followed by unsupervised clustering reveals protein modules that represent key cell types and biological processes in the plasma proteome

We identified major cell types and biological processes represented in the plasma proteome using a computational framework that consists of two sequential steps: dimension reduction and clustering (**Figure 1**). In this framework, a vector of 51,180 elements is assigned to each protein, representing the normalized expression levels of the plasma protein measured in 51,180 participants. To derive an unentangled representation that captures the main sources of variation of protein abundances in the cohort, we built a variational autoencoder (VAE) that projected the protein vector into a lower dimensional latent space while minimizing the reconstruction loss relative to the original vector (for details see Methods). Next, we performed unsupervised clustering of proteins with Leiden algorithm [7] based on their proximity within the derived latent space. Proteins that are close to each other in the latent space embedding share similar expression profiles and are therefore more likely to originate from the same cell type/tissue or participate in the same biological process. The advantage of carrying out clustering on the latent space instead of the original input space is twofold: 1) recently developed non-linear clustering algorithms such as Leiden and Louvain are computationally expensive, therefore reducing the number of data dimension alleviates the computation burden; and 2) it mitigates the biases introduced by the presence of extensive correlations among the original features as VAE is designed to minimize correlation among the derived latent dimensions. With this computational framework, we assigned each plasma protein to one of the 36 distinct protein modules (**Supplementary Table S1**).

**Figure 1.**
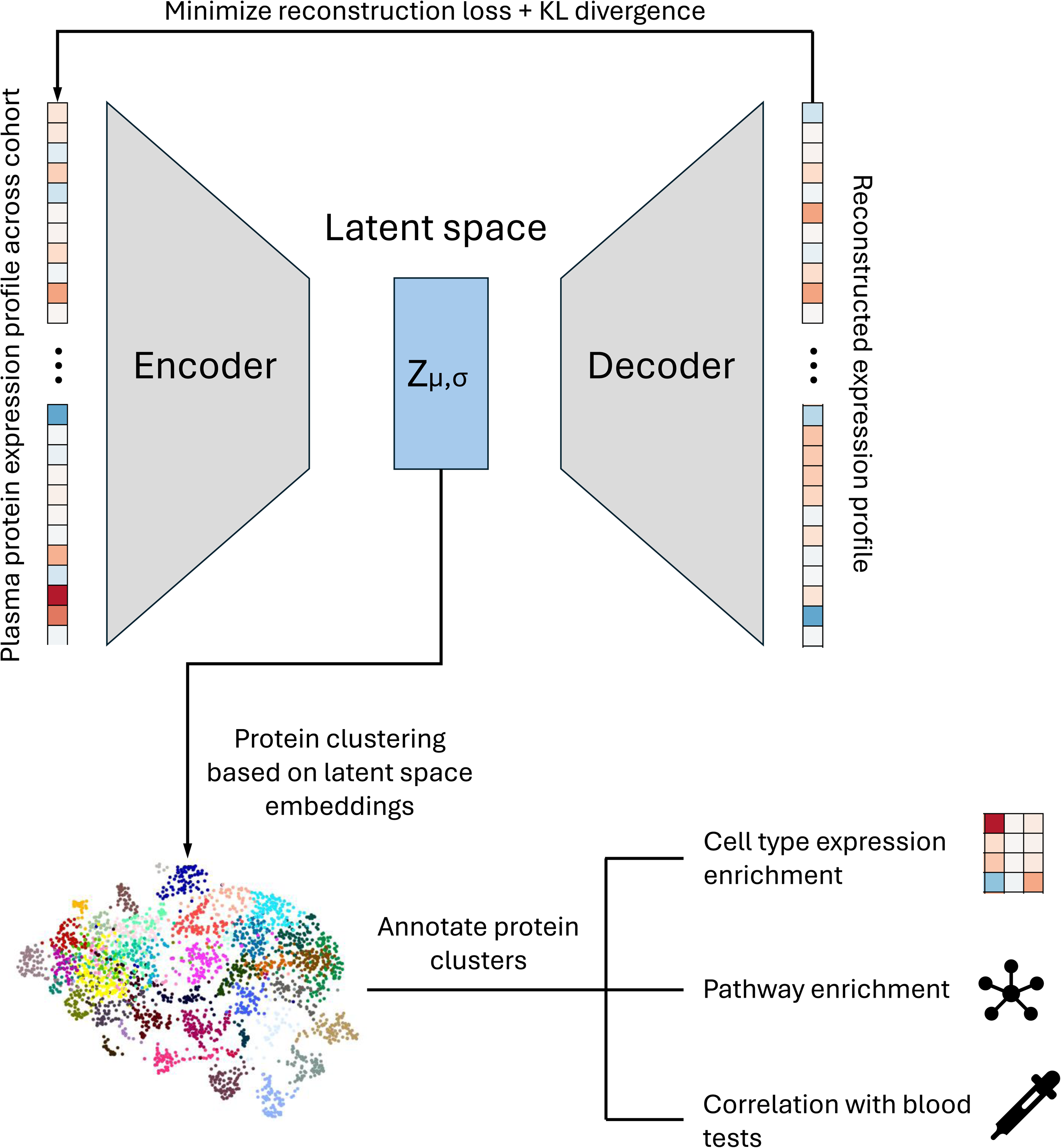
A schematic representation of the workflow that derives and annotates biologically relevant protein modules based on Olink proteomics measurements from the UKB cohort.

To test whether proteins in those modules are enriched in any cell type or biological process, we leveraged information from three orthogonal sources (for detail see Methods): 1) transcriptomic profiles of 54 major human tissues from GTEx and 116 cell types from Tabular Sapiens consortium [8, 9] (**Supplementary Figure S1 & S2**); 2) Biological processes and pathways annotated in Gene Ontology, KEGG and REACTOME (**Supplementary Table S2**); and 3) Blood test results including major blood cell counts and percentages generated in the same UK Biobank participants (**Supplementary Figure S3**). Information regarding preferential expression in tissues or cell types, enrichment in annotated biological processes and pathways, and correlation with blood cell/metabolite measurements were comprehensively evaluated before we manually assigned the protein modules to the cell type or biological process that they most likely represent. With this approach, we were able to annotate 32 of the 36 modules (**Figure 2a**).

**Figure 2.**
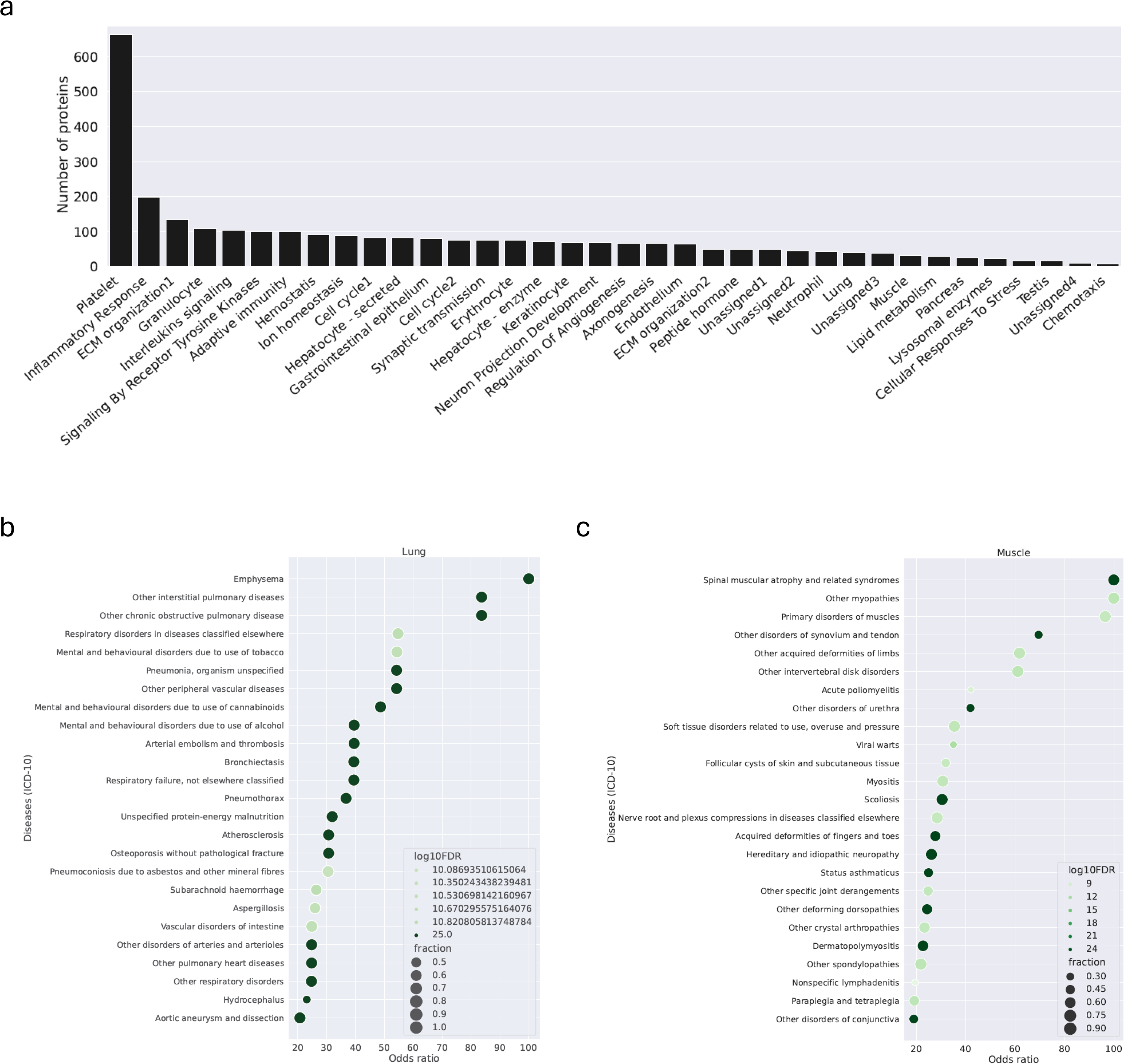
**a)** Number of member proteins in each of the identified protein modules. b) & c) Diseases in which up-regulated plasma proteins are enriched in the lung (b) and muscle (c) modules. Shades of green represents the logarithm of false discovery rate. Sizes of the dots are proportional to the fraction of up-regulated proteins that belong to the module.

### Protein modules dysregulated in disease

We examined how those protein module levels vary by disease conditions. We used 3-digit ICD-10 codes in the UKB clinical record to determine the participants’ disease status and identified plasma proteins that are associated with diseases by fitting a logistic regression model (Methods). For each module – disease pair, we tested if the proteins up- or down-regulated in the disease significantly overlap with those belonging to the module (for details see Methods).

We uncovered many associations that match the tissue or known biology of the diseases (**Supplementary Table S3**). For instance, proteins up regulated in lung diseases including emphysema, interstitial pulmonary disease and chronic obstructive pulmonary disease are significantly enriched in the lung module (**Figure 2b**). Similarly, among the top diseases whose up-regulated proteins are overrepresented in the muscle module are spinal muscular atrophy, myopathies and primary disorders of muscles, possibly reflecting muscle tissue damage accompanying disease process (**Figure 2c**). Lichen planus and alopecia areata, both of which are autoimmune diseases of the skin, are associated with down regulation of keratinocyte module proteins (**Supplementary Table S3**), consistent with a loss of skin or follicle cells under immune system attacks.

### Discovery of common genetic variants associated with changes in protein module levels

The UKB PPP consortium reported a comprehensive protein quantitative trait loci (pQTL) mapping that included 2,103 pleiotropic trans-pQTLs [1], where a locus is associated with the plasma abundances of multiple distal proteins. We hypothesized that one of the possible mechanisms underlying pleiotropic trans-pQTLs is when a genetic variant alters the overall contribution of a tissue/cell type or biological process, therefore affecting multiple proteins originating from the cell type or belonging to the biological process. We tested this hypothesis by testing whether proteins in the VAE-defined protein modules were systematically associated with selected genetic variants. In our analysis, we included 116,902 common genetic variants that are either significant quantitative trait loci (eQTL and pQTL) or coding variants with moderate to high impact predicted by Ensembl Variant Effect Predictor (VEP) [10] (for details see Methods). For each variant of interest, we collected the summary statistics from associations with 2,935 assays encompassing 2,917 unique plasma proteins and then for each protein module tested if the normalized effect sizes in the module significantly deviate from zero by fitting a Bayesian Regression model (for details see Methods). We identified 14,932 significant variant-module associations with false discovery rate < 1e-30 and estimated effect size > 1 (**Supplementary Table S4 & Figure S4**). Platelet and erythrocyte modules were associated with the largest number of loci, suggesting that the plasma proteome is highly sensitive to changes in these abundant blood cell types. One mechanism by which a locus associates with a protein module is via association with the cell counts. For instance, intronic SNP rs3917932 (1_36478315_C_G) is a splicing quantitative trait locus (sQTL) for the *CSF3R* gene, which encodes a receptor for granulocyte colony-stimulating factor (encoded by the *CSF3* gene). It significantly associates with elevated level of CSF3R and decreased level of CSF3 proteins in plasma, as well as reduced abundance of neutrophil module proteins and lower neutrophil counts in the blood (**Figure 3b & c**). A plausible explanation for the observed associations is that instead of localized to the plasma membrane, CSF3R proteins encoded by the splicing isoform are predominantly released into circulation, bind to and sequester CSF hormones in the blood, thus leading to suppressed neutrophil maturation, low neutrophil counts and low concentrations of neutrophil proteins. Another possible mechanism underlying the association between a locus and the abundances of a protein module is through dysregulation of protein secretion. SNP rs10745925 (12_101825121_T_C) is an eQTL for the *GNPTAB* gene, which encodes subunits of the N-acetylglucosamine-1-phosphotransferase that catalyzes the formation of mannose 6-phosphate (M6P) markers on high mannose type oligosaccharides in the Golgi apparatus [11]. M6P modification is essential for M6P-receptor-mediated transport of lysosomal enzymes into lysosomes, therefore explaining the association between the genetic variant and the dramatic change in lysosomal enzyme abundances in plasma (**Figure 3d**).

**Figure 3.**
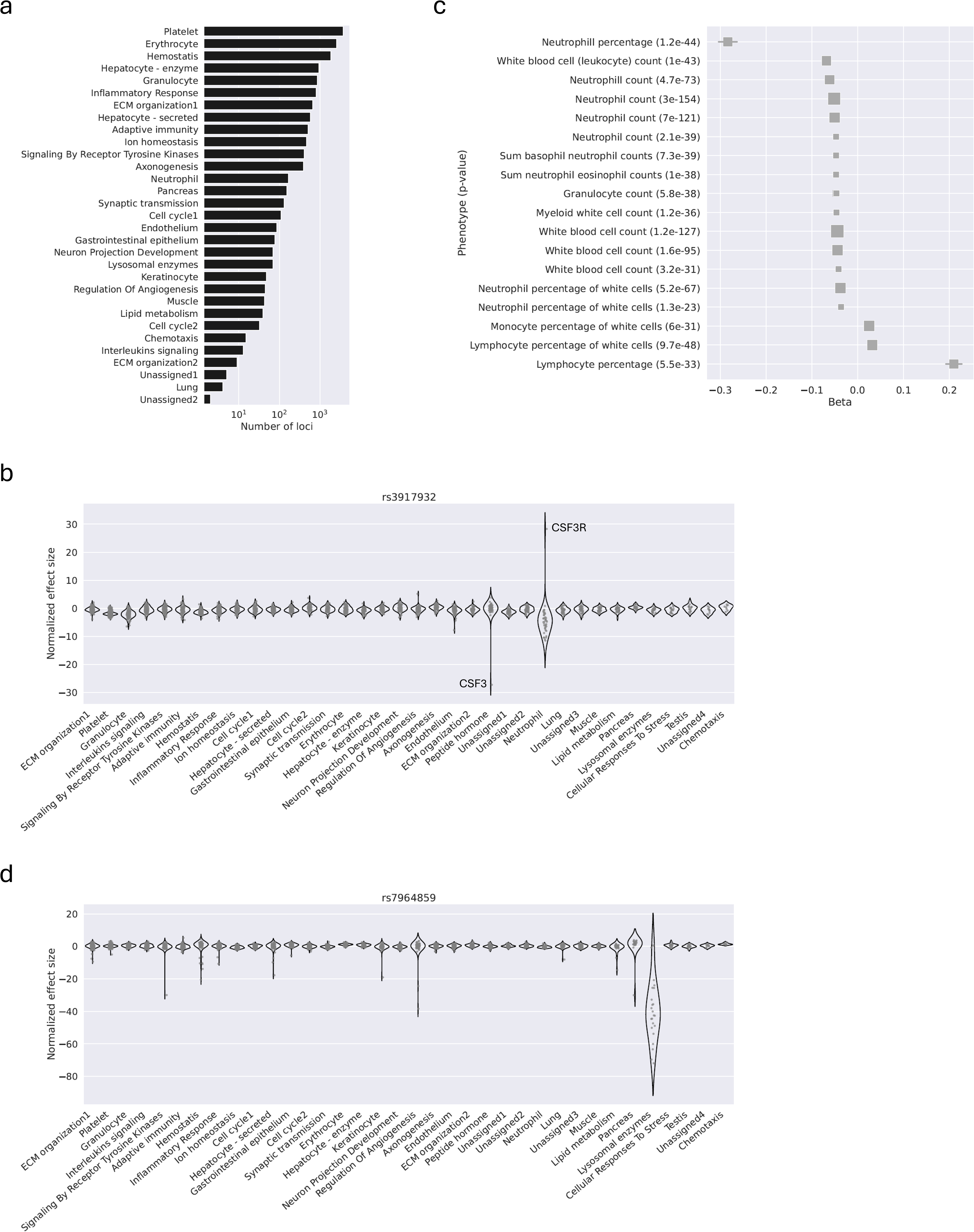
**a)** Number of loci significantly associated with the abundance of each of the protein modules. Only modules associated with at least one locus are shown. b) Distribution of normalized effect sizes of genetic variant rs3917932 (1_36478315_C_G) across protein modules. c) Forest plot showing phenotypes associated with variant rs3917932 (1_36478315_C_G). Sizes of the boxes are proportional to the study cohort sizes. Numbers in the parathesis on y-axis labels are *p*-values. All associations with *p*-value < 1e-30 are shown. d) Distribution of normalized effect sizes of genetic variant rs7964859 (12_101825121_T_C) across protein modules.

Gene-level collapsing analysis based on rare coding variants had previously revealed significant associations between loss of *GNPTAB* gene function and elevated lysosomal enzyme concentrations in plasma [12]. It is consistent with our observation that rs10745925, an eQTL associated with increased expression of *GNPTAB*, is also associated with decreased plasma abundances of lysosomal enzymes.

### Protein modules help interpret effects of genetic variants associated with complex diseases

Plasma protein modules representing key cell types and biological processes may also be employed to analyze and interpret disease-associated genetic variants and to potentially shed new light on disease mechanisms. We searched published GWAS results hosted on Open Targets Genetics [13] and discovered that 354 of the identified module-modifying loci were associated with diseases (168 unique diseases) across multiple organ systems (for details, see Methods; **Figure 4a**). Examining the protein modules (and the cell types or pathways they represent) dysregulated by the disease-associated variants offers a unique opportunity to better understand their downstream effects. For instance, SNP rs738409, a missense variant in the *PNPLA3* gene that encodes a triacylglycerol lipase and regulates liver fat metabolism, has been found to strongly associate with liver cirrhosis and non-alcoholic fatty liver disease [14].

**Figure 4.**
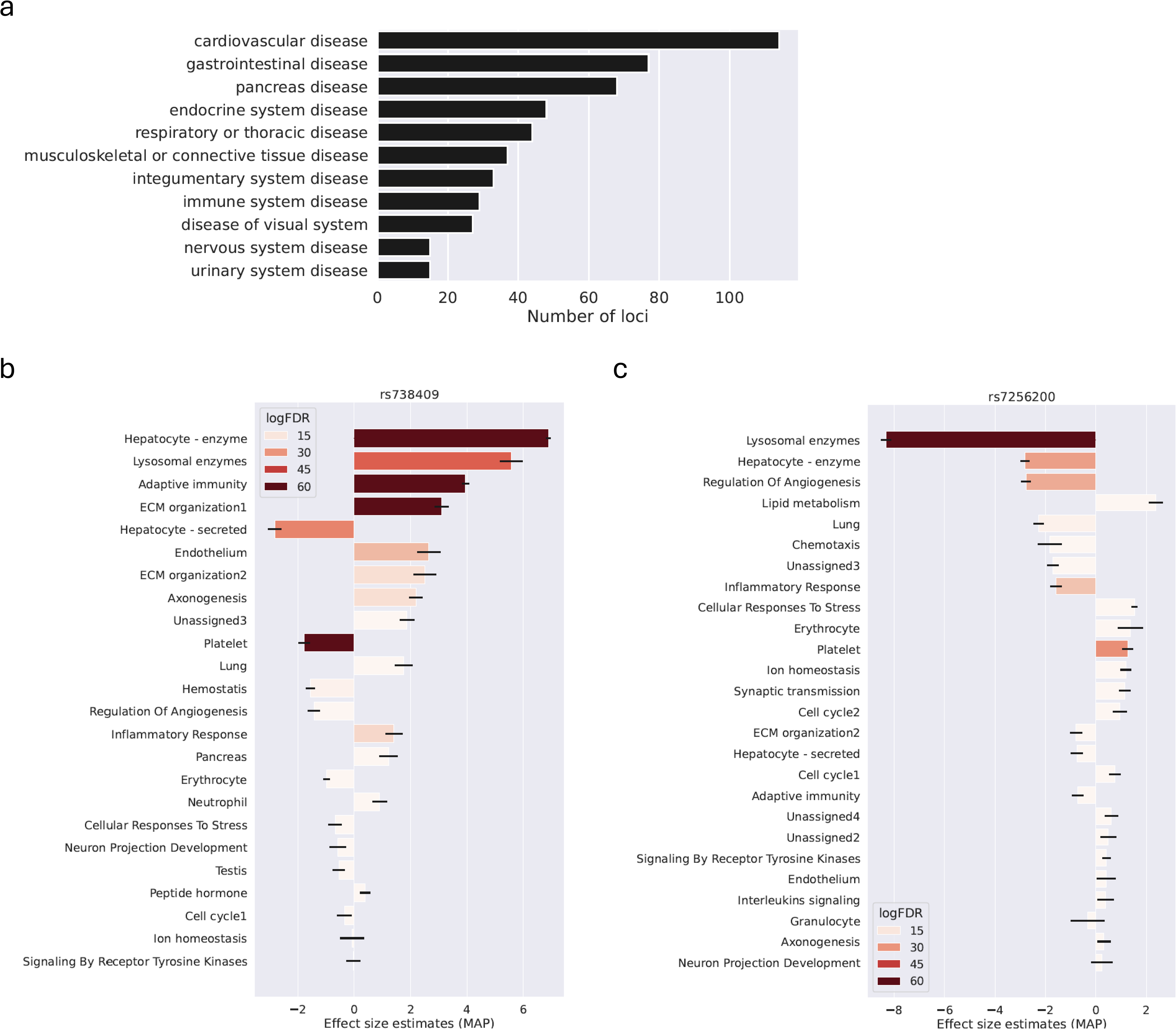
**a)** Number of module-modifying loci associated with trait categories. b) & c) Distribution of estimated effect sizes (MAP) of genetic variants rs738409 (22_43928847_C_G) (b) and rs7256200 (19_44912678_G_T) (c) on inferred protein modules. Shades of red are proportional to the logarithm of false discovery rate. Error bars represent posterior standard deviation.

Consistent with previously published findings [15], the alternative allele of rs738409 (G) was associated with increased abundances of liver enzymes in our analysis, possibly reflecting hepatocyte damage incurred by the accumulation of lipid droplets (**Figure 4b**). Additionally, proteins in the lysosome, adaptive immunity, extracellular matrix and inflammation modules were also positively associated with rs738409. On the other hand, liver secretory proteins abundances were negatively associated with rs738409. These additional associations represent either secondary effects downstream of the primary effect (in this case the accumulation of lipid droplets in hepatocytes) or novel, independent events occurring in other cell types/tissues. SNP rs7256200 is a known risk variant for late onset Alzheimer’s Disease (AD) located near the *APOC1* and *APOE* genes [16]. It is strongly associated with decreased levels of lysosomal enzymes in addition to lower APOE protein level in plasma (**Figure 4c**). Interestingly, significantly higher plasma abundance of myeloid and erythroid nuclear termination stage-specific protein (MENT), a protease inhibitor that inhibits several lysosomal enzymes including cathepsin K, cathepsin L, and cathepsin V [17], is also associated with the SNP. Dysregulation of lysosomal function and related autophagy process has been thought to play a key role in AD and age-related dementia [18]. Although by itself not a proof of causal relationship, the fact that an AD risk variant is simultaneously associated with lower plasma abundance of lysosomal proteins lends more weight to the hypothesis that lysosomal dysfunction could be one of the potential causes of AD and dementia.

Many complex diseases are associated with multiple genetic loci, which may constitute different components of the same pathway or reflect the presence of multiple molecular mechanisms underlying disease risks, possibly with distinct etiologies or progression trajectories. Disease variants’ impacts on the plasma proteome may help reveal potential disease subtypes. Type 1 diabetes is an autoimmune disease in which the immune system attacks the secretory cells in pancreas and affects insulin secretion. We found 34 module-modifying loci that were also associated with type 1 diabetes and by comparing their impacts on the protein modules we identified several variant clusters (**Figure 5a**). We also examined other diseases associated with these loci, hoping that additional phenotypic data may aid the interpretation of derived variant clusters. We found 6 loci significantly associated with pancreatic enzyme levels (labels colored in blue) and none of them is associated with additional autoimmune diseases besides type 1 diabetes. Whereas for loci that do not affect pancreas protein abundances, 60.7% (17 out of 28) of them are associated with additional autoimmune diseases besides type 1 diabetes (**Figure 5a**). The depletion (p-value = 0.024, chi-square test) of associations with additional autoimmune diseases for the pancreatic protein perturbing loci suggests that their association with type 1 diabetes may be mediated by a dysregulation in the pancreas that engenders autoimmune attacks against it. On the other hand, other variants likely act through pancreas-independent mechanisms that modulate the risk for not only type 1 diabetes but potentially other autoimmune diseases as well. Moreover, we found 14 genetic loci associated with dysregulation of proteins in the lipid metabolism module (**Figure 5b**). We observed that most (12 out of 14) of those loci were significantly associated with cardiovascular and metabolic diseases, which is consistent with the role that dysregulated lipid metabolism and obesity play in cardiovascular diseases and metabolic diseases such as type 2 diabetes [19, 20]. The direction of the association between a module-modifying variant and plasma protein levels may also be predictive of the direction of the variant’s effect on disease risks. We analyzed 175 genetic associations from a GWAS on coronary artery disease (CAD) [21] and found 21 loci are significantly associated with an extracellular matrix organization module (FDR < 1e-20). We noticed that variants associated with decreased levels of ECM organization module proteins are predominantly (9 out of 10) risk variants for CAD while most of the variants associated with increased levels of ECM organization module (10 out of 11) are protective against CAD (**Figure 5c**, p-value = 0.00076, chi-square test).

**Figure 5.**
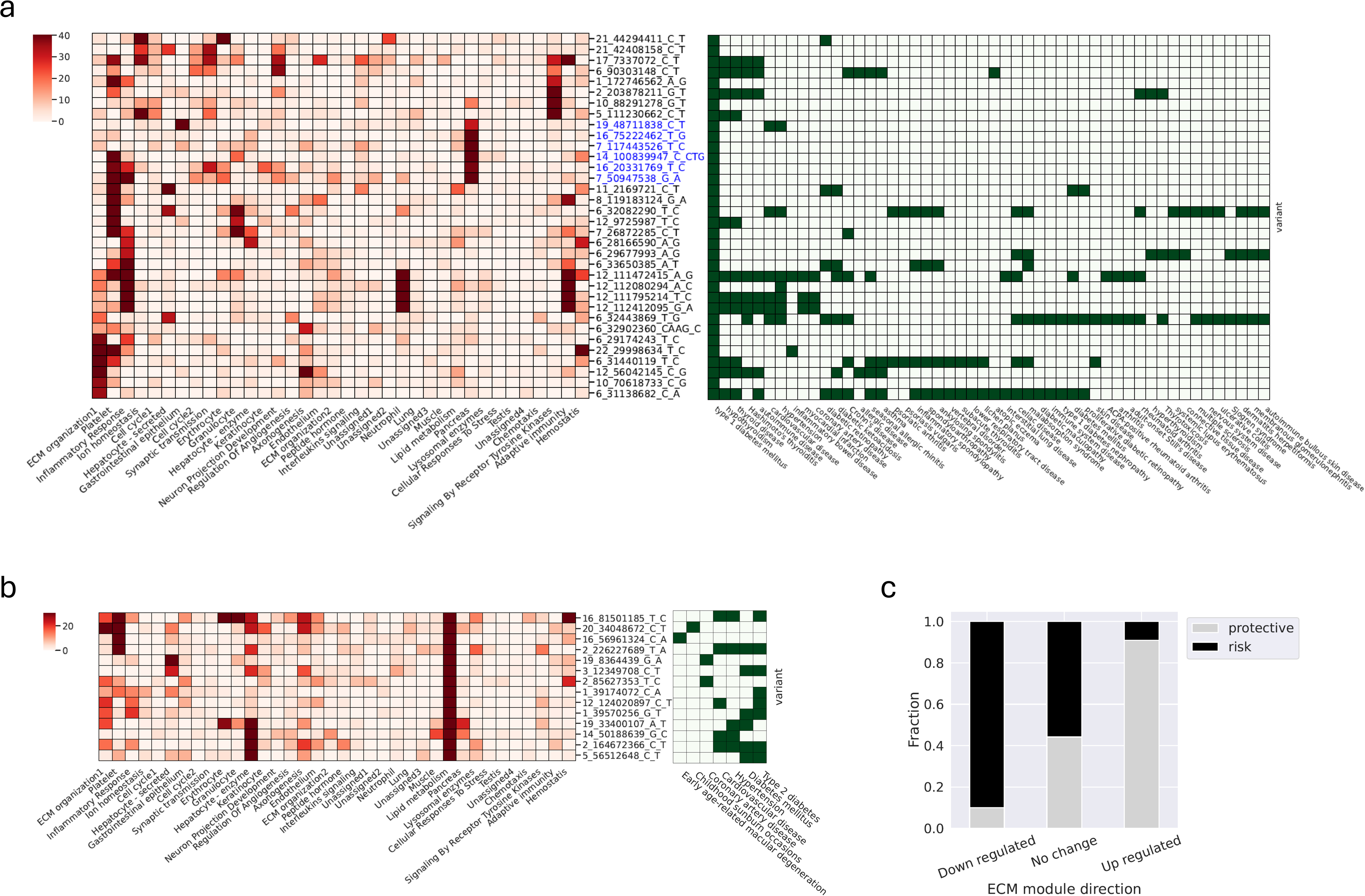
**a)** The heatmap on the left depicts the significance of associations between variants (rows) and protein modules (columns) for module-modifying variants that are also associated with type 1 diabetes. Shades of red are proportional to the logarithm of false discovery rate of the association. Variants are clustered using hierarchical clustering. Variants with blue labels belong to a cluster that show strong association with the pancreas module. The heatmap on the right depicts association between the same variants (in the same order as the left heatmap) and diseases. Green grids indicate the presence of significant association while white grids indicate the absence of association. b) Same as a) except for variants that are associated with changes in the lipid metabolism module. c) Fraction of variants that confer protection versus risk for coronary artery disease grouped by the direction of their effects on the extracellular matrix organization module.

## Discussion

In this report, we applied a data-driven approach to deconvolute 2,935 plasma proteins into modules that likely represent major cell types and biological pathways present in the plasma proteome. The protein modules we found cover a wide range of cell types and tissues, supporting the notion that plasma proteome could become a useful tool for monitoring the health status of multiple organ systems and tissues. It comes as no surprise that several protein modules are assigned to blood cell types including platelets, erythrocytes, neutrophils and immune cells, given their direct access to the circulation. The platelet module was the largest module encompassing more than a fifth of all proteins in the Olink Explore 3072 panel. It is worth noting that many of the proteins belonging to the platelet module are not platelet specific and some of them even have enriched expression in other tissues/cell types (**Supplementary Figure S1 & S2**). They were assigned to the platelet module because they share similar expression profiles with platelet-specific proteins such as Glycoprotein V Platelet (GP5) and Glycoprotein VI Platelet (GP6) and their abundance correlates with platelet counts. A likely explanation is that although those proteins are only moderately expressed in platelets, the proportion of the platelet contribution to the plasma proteome is so high that it constitutes a major source for those proteins in blood. This illustrates the potential pitfalls of relying solely on the expression levels measured in independent tissues/cell types and highlights the merit of taking a more data-driven approach to assign tissue/cell type origin for plasma proteins. A more reliable inference of tissue/cell type origin for plasma proteins will better inform panel design and protein selection for future biomarkers. With more proteins being added to the high-throughput assay panels, our approach may reveal more cell types or biological processes reflected in the plasma proteome in future studies.

Modulation of gene expression at the transcription level is merely one of the many possible mechanisms by which a genetic variant may alter the abundance of a protein in circulation. The discovery of numerous trans-pQTL loci from the original UKB-PPP report indicates extensive downstream effects beyond transcriptional modulation of the nearby *cis* gene that may include changes in cell counts, protein subcellular location (e.g., intracellular/membrane-bound versus secreted), protein leakage due to cell necrosis or apoptosis, and effects on transcription factor target gene expression. Those secondary effects often impact multiple proteins with relatively moderate effect sizes. Conventional pQTL analysis treats each protein as independent and adopts stringent *p*-value threshold to account for potential false discoveries due to multiple testing. The identification of protein modules allows us to consider a group of functionally related proteins as a whole and increases the statistical power of detecting associations with smaller effect sizes that would have been missed in conventional single protein pQTL analysis. For instance, rs1126313 (1_39174072_C_A) is significantly associated with decreased lipid metabolism module level (False Discovery Rate = 1.72e-39) while none of the individual proteins belonging to this module passed the significance threshold (nominal p-value < 1.7e-11) used in the pQTL analysis [1]. We highlighted examples of variant – protein module associations with known mechanisms and reported novel ones discovered in our analysis. Our findings suggested that genetic factors play a role in determining the relative contributions of many cell types and biological processes to the plasma proteome, and it is therefore crucial to incorporate genotypic information as a covariate in the training and classification of plasma protein-based models to adjust for variations in baseline levels across genetically diverse cohorts.

Genome-wide association studies have uncovered numerous genetic loci that are associated with predisposition to complex diseases. However, it is extremely challenging to unravel the molecular mechanisms by which genetic variation modulates the risk for a disease. Changes in plasma protein abundances associated with a disease variant may offer additional insight into the cell types and/or biological processes affected by the variant and lead to new hypothesis about the disease mechanisms. Data-driven grouping of plasma proteins into functional modules enables intuitive interpretation of disease variant effects and facilitates the identification of key cell types and biological processes for a given disease. We demonstrated examples where we use this approach to pinpoint the known biological process crucial to certain diseases (lipid metabolism in cardiovascular diseases and type 2 diabetes) and to reveal multiple potential etiologies (pancreas specific versus general autoimmune activity in the case of type 1 diabetes). We note that not all proteomic changes associated with a disease variant are necessarily relevant to the disease and a holistic evaluation of additional evidence and known biology of the disease is essential. Nonetheless, the ability to link a disease variant to alterations in certain cell types and biological processes will provide valuable clues for a better understanding of disease mechanisms and empower target discovery for future therapeutics.

## Methods

### UKB data

UK Biobank is a longitudinal cohort study of >500,000 participants aged 40-69 years enrolled in study centers across England, Wales and Scotland between 2006 and 2019 [22]. In addition to extensive baseline data about medical histories, physical measurements and health behavior of participants, biosamples were collected from the participants (e.g., saliva, urine and blood).

Ethical approval was granted for the UK Biobank by the North West Multi-Centre Research Ethics Committee and the National Health Service (NHS) National Research Ethics Service (ref: 11/NW/0382), all participants provided written informed consent, and all experiments were conducted in accordance to relevant regulations and guidelines. Additional information, including details on study protocol, are available online at https://biobank.ctsu.ox.ac.uk/. For a subset of 54,219 UKB participants, proteomic measurements were conducted by the UKB PPP consortium as described elsewhere [1]. Plasma proteomics data, genetic data and health data from the UKB participants were accessed under UK Biobank applications 65851 and 26041.

### Representation learning with variational autoencoder and unsupervised clustering of proteins

Inverse rank normalized protein abundance measurements were used for representation learning. Proteins with missing values in >= 10% of the subjects were excluded, with 2,916 proteins (2,935 assays) retained for subsequent analysis. Subjects with missing values in >= 20% of the remaining proteins were excluded, with 51,180 subjects retained in the subsequent analysis. Both the encoder and decoder in the VAE are multilayer perceptron (MLP) with 2 hidden layers. The latent space has 10 dimensions. The layer sizes of the encoder network were 51,180 (input layer) – 512 (hidden layer) – 64 (hidden layer) – 20 (output layer, 10 for mean and 10 for variance). The layer sizes of the decoder were 10 (input layer) – 64 (hidden layer) – 512 (hidden layer) – 51,180 (output layer). Since the inverse rank normalized values approximately followed normal distribution, we defined the reconstruction loss as the difference of cumulative distribution function between the input and decoder output. The regularization term added to the loss function is the *KL* divergence as in standard VAE. The model was trained in 150 epochs, with batch size of 32 and learning rate 0.0001. The training of the VAE model was carried out with the PyTorch library (version 1.13.1). Leiden algorithm implemented in the Scanpy (version 1.10.1) package was used to perform protein clustering based on their coordinates in the latent embedding, with resolution parameter being set to 2.5. For each of the 45 initial modules identified, we calculated the coefficient on the 1^st^ principal component as an estimate of the average level of that module across the cohort and computed pairwise correlations among the 45 modules. We then merged modules that are highly correlated across the subjects and reduced the number of modules to 36.

### Annotation of protein modules and inference of tissue/cell type origin

For each of the 36 protein modules, we tested expression enrichment for the transcripts encoding member proteins across human tissues (GTEx RNA-seq) [8] and cell types (Tabular Sapiens scRNA-seq) [9] (**Supplementary Figure S1 & S2**). We also performed pathway enrichment analysis against annotated pathways/biological processes in Gene Ontology, KEGG and Reactome databases for each protein module using the enrichR package [23] (**Supplementary Table S2**). Furthermore, we correlated the 1^st^ principal component coefficients of each protein module with demographic information and blood assay measurements across the cohort (**Supplementary Figure S3**) using the Pearson correlation function (scipy.stats.pearsonr) in the SciPy library (version 1.7.3). The cell type/biological process assignment was made after manual review of the results derived from analyses described above.

### Assessing protein modules dysregulation in diseases

Case-control logistic regression analysis was performed on first occurrences data (Category 1712, which included hospital inpatient data, self-reports, primary care data and death register records) defined by UK Biobank for all 3-digit ICD-10 codes. Covariates included in the regression are: time between plasma sample and assay (tbms_covar) + age + sex + age_sex + age2 + age2_sex + factor(Proteomics Batch) + factor(ukb_centre_fct) + factor(array_fct) + 20 genetic principal components. We defined up- and down-regulated proteins for each ICD-10 code with a loose threshold of false discovery rate (FDR) < 0.1. Next, for each disease (3-digit ICD-10 code)-module pair we tested if the proteins up-/down-regulated in the disease are significantly enriched in the module with hypergeometric test, which was implemented using “scipy.stats.hypergeom.cdf” function in the Scipy library (version 1.7.3). False discovery rate (“Benjamini-hochberg” method) based adjustment for multiple testing correction was carried out using the “statsmodels.stats.multitest” function in the statsmodels library (version 0.13.5). Significant associations with FDR < 1e-5 and number of overlapped proteins > = 25 and fraction of overlapped proteins > 0.05 are included in Supplementary Table S3.

### Identify associations between selected genomic loci and protein module levels

Full summary statistics from genome-wide pQTL analysis generated by the UKB PPP consortium was downloaded from Synapse (https://doi.org/10.7303/syn51364943). We selected genetic variants from 3 sources for the subsequent analysis: 1) exon variants reported in gnomAD v4.1 with minor allele frequency >= 1% in at least one ancestry group and predicted impact >= MODERATE by the Variant Effect Predictor (VEP) [10, 24]; 2) significant eQTL loci (nominal *p*-value < 1e-15) from eQTL Catalogue [25] (with linkage disequilibrium clumping for variants within 500kb window and r^2^ >= 0.8); 3) significant pQTL loci reported by the UKB PPP consortium [1]. The combination of the above three sources led to a total of 116,902 variants with summary statistics. For each variant, we calculated the normalized effect size on each protein by dividing the estimated coefficient (β) by the standard error. To minimize the impact of outliers, normalized effect sizes that are more than 5 standard deviations (s.d.) away from the mean are fixed at mean ± 5 s.d. We then fitted a Bayesian linear regression model with a centered elliptic Gaussian distribution as the prior on the normalized effect sizes of proteins belonging to a protein module. It is also known as Automatic Relevance Determination (ARD) model which automatically prunes features that are estimated to have no effects and hence leads to sparser solutions. The fitting was carried out using the “sklearn.linear_model.ARDRegression” function in the scikit-learn library (version 1.0.2). The maximum a posteriori (MAP) values of the coefficients were taken as the estimates for the effect sizes of the modules. P-values were calculated from the posterior (normal) distributions of the coefficients. False discovery rate (“Benjamini-hochberg” method) based adjustment for multiple testing was carried out using the “statsmodels.stats.multitest” function. LD clumping was performed within 500kb windows to retain only the most significant variant to represent the locus. Significant associations with FDR < 1e-30 and absolute MAP > 1 are included in Supplementary Table S4.

### Disease associations for module-modifying variants

Disease associations for module-modifying variants were obtained from Open Targets Genetics platform via the “pheWAS” API endpoint. Only associations with *p*-value < 5e-8 were retained in subsequent analysis. Diseases belonging to the “cell proliferation disorder” category were not shown in Figure 5b & c. The summary statistics for the coronary artery disease study (GCST010866) were downloaded from the GWAS Catalog.

## Data Availability

Data from UK biobank cannot be shared openly and are subject to UK Biobank ethical approval. Further information about applying for UK Biobank data access can be obtained from the UK Biobank website (https://www.ukbiobank.ac.uk) or by emailing UK Biobank.

## Acknowledgements

This research has been conducted using the UK Biobank Resource under application numbers 65851 and 26041. We thank the UK Biobank team and participants for making this valuable resource possible. We thank the research and development teams at the 13 participating UKB-PPP companies (Alnylam Pharmaceuticals, Amgen, AstraZeneca, Biogen, Calico, Bristol-Myers Squibb, Genentech, GlaxoSmithKlein (GSK), Janssen Pharmaceuticals, Novo Nordisk, Pfizer, Regeneron and Takeda) for funding the proteomics measurements.

## Author contributions

J.Z. conceived and designed the analysis. J.Z. and E.N.S. conducted the analysis. J.Z., E.N.S. and D.D wrote the manuscript.

