## Supplementary material for "Representation learning based on proteomic profiles uncovers key cell types and biological processes contributing to the plasma proteome": FigureS2.pdf

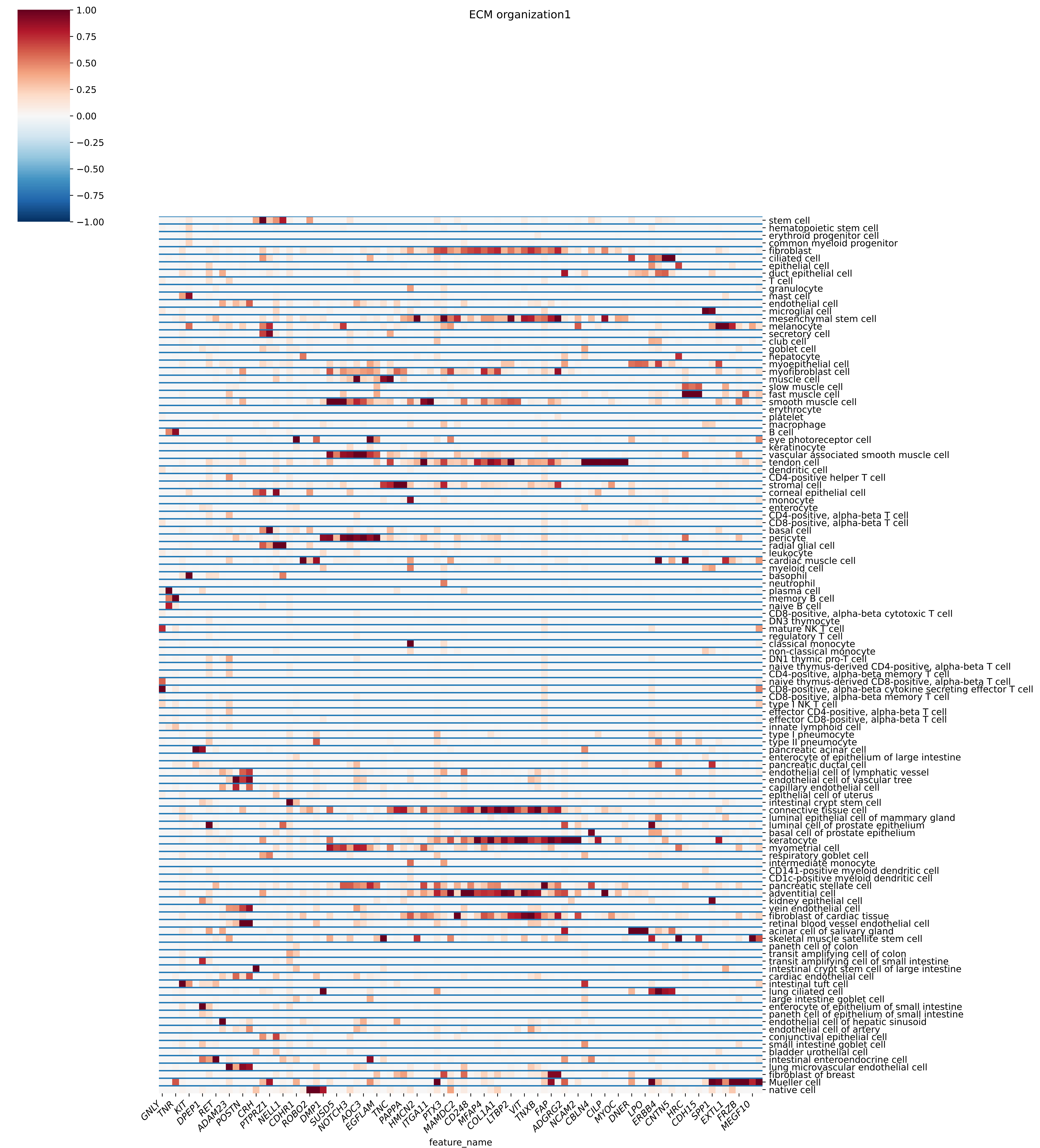
ECM organization1
1.00
0.75
0.50
0.25
0.00
−0.25
−0.50
−0.75
−1.00
stem cell
hematopoietic stem cell
erythroid progenitor cell
common myeloid progenitor
fibroblast
ciliated cell
epithelial cell
duct epithelial cell
T cell
granulocyte
mast cell
endothelial cell
microglial cell
mesenchymal stem cell
melanocyte
secretory cell
club cell
goblet cell
hepatocyte
myoepithelial cell
myofibroblast cell
muscle cell
slow muscle cell
fast muscle cell
smooth muscle cell
erythrocyte
platelet
macrophage
B cell
eye photoreceptor cell
keratinocyte
vascular associated smooth muscle cell
tendon cell
dendritic cell
CD4-positive helper T cell
stromal cell
corneal epithelial cell
monocyte
enterocyte
CD4-positive, alpha-beta T cell
CD8-positive, alpha-beta T cell
basal cell
pericyte
radial glial cell
leukocyte
cardiac muscle cell
myeloid cell
basophil
neutrophil
plasma cell
memory B cell
naive B cell
CD8-positive, alpha-beta cytotoxic T cell
DN3 thymocyte
mature NK T cell
regulatory T cell
classical monocyte
non-classical monocyte
DN1 thymic pro-T cell
naive thymus-derived CD4-positive, alpha-beta T cell
CD4-positive, alpha-beta memory T cell
naive thymus-derived CD8-positive, alpha-beta T cell
CD8-positive, alpha-beta cytokine secreting effector T cell
CD8-positive, alpha-beta memory T cell
type I NK T cell
effector CD4-positive, alpha-beta T cell
effector CD8-positive, alpha-beta T cell
innate lymphoid cell
type I pneumocyte
type II pneumocyte
pancreatic acinar cell
enterocyte of epithelium of large intestine
pancreatic ductal cell
endothelial cell of lymphatic vessel
endothelial cell of vascular tree
capillary endothelial cell
epithelial cell of uterus
intestinal crypt stem cell
connective tissue cell
luminal epithelial cell of mammary gland
luminal cell of prostate epithelium
basal cell of prostate epithelium
keratocyte
myometrial cell
respiratory goblet cell
intermediate monocyte
CD141-positive myeloid dendritic cell
CD1c-positive myeloid dendritic cell
pancreatic stellate cell
adventitial cell
kidney epithelial cell
vein endothelial cell
fibroblast of cardiac tissue
retinal blood vessel endothelial cell
acinar cell of salivary gland
skeletal muscle satellite stem cell
paneth cell of colon
transit amplifying cell of colon
transit amplifying cell of small intestine
intestinal crypt stem cell of large intestine
cardiac endothelial cell
intestinal tuft cell
lung ciliated cell
large intestine goblet cell
enterocyte of epithelium of small intestine
paneth cell of epithelium of small intestine
endothelial cell of hepatic sinusoid
endothelial cell of artery
conjunctival epithelial cell
small intestine goblet cell
bladder urothelial cell
intestinal enteroendocrine cell
lung microvascular endothelial cell
fibroblast of breast
Mueller cell
native cell
GNLY
TNR
KIT
DPEP1
RET
ADAM23
POSTN
CRH
PTPRZ1
NELL1
CDHR1
ROBO2
DMP1
SUSD5
NOTCH3
AOC3
EGFLAM
TNC
PAPPA
HMCN2
ITGA11
PTX3
MAMDC2
CD248
MFAP4
COL1A1
LTBP2
VIT
TNXB
FAP
ADGRG2
NCAM2
CBLN4
CILP
MYOC
DNER
LPO
ERBB4
CNTN5
HRC
CDH15
SPP1
EXTL1
FRZB
MEGF10
feature_name

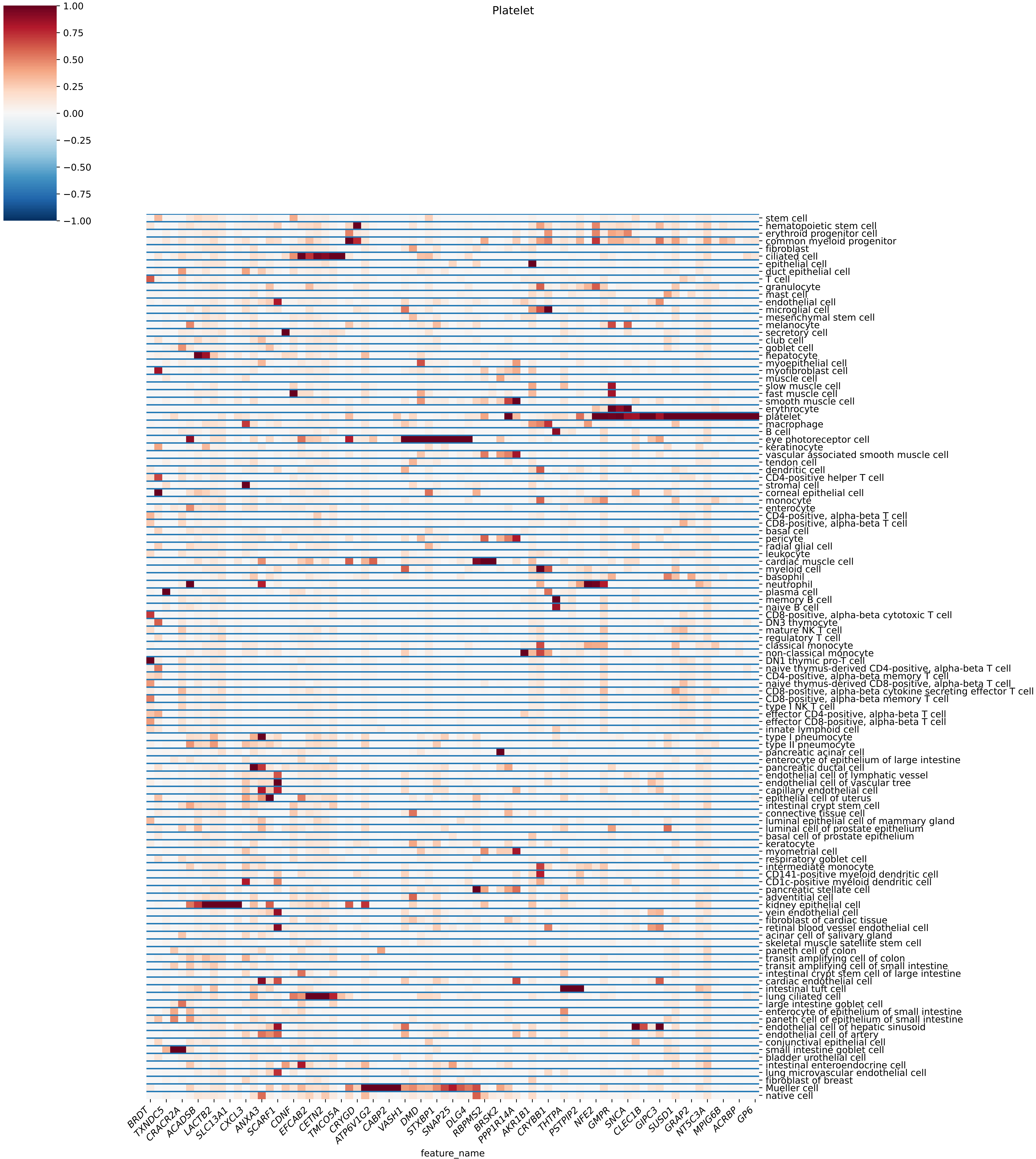

Platelet
1.00
0.75
0.50
0.25
0.00
−0.25
−0.50
−0.75
−1.00
stem cell
hematopoietic stem cell
erythroid progenitor cell
common myeloid progenitor
fibroblast
ciliated cell
epithelial cell
duct epithelial cell
T cell
granulocyte
mast cell
endothelial cell
microglial cell
mesenchymal stem cell
melanocyte
secretory cell
club cell
goblet cell
hepatocyte
myoepithelial cell
myofibroblast cell
muscle cell
slow muscle cell
fast muscle cell
smooth muscle cell
erythrocyte
platelet
macrophage
B cell
eye photoreceptor cell
keratinocyte
vascular associated smooth muscle cell
tendon cell
dendritic cell
CD4-positive helper T cell
stromal cell
corneal epithelial cell
monocyte
enterocyte
CD4-positive, alpha-beta T cell
CD8-positive, alpha-beta T cell
basal cell
pericyte
radial glial cell
leukocyte
cardiac muscle cell
myeloid cell
basophil
neutrophil
plasma cell
memory B cell
naive B cell
CD8-positive, alpha-beta cytotoxic T cell
DN3 thymocyte
mature NK T cell
regulatory T cell
classical monocyte
non-classical monocyte
DN1 thymic pro-T cell
naive thymus-derived CD4-positive, alpha-beta T cell
CD4-positive, alpha-beta memory T cell
naive thymus-derived CD8-positive, alpha-beta T cell
CD8-positive, alpha-beta cytokine secreting effector T cell
CD8-positive, alpha-beta memory T cell
type I NK T cell
effector CD4-positive, alpha-beta T cell
effector CD8-positive, alpha-beta T cell
innate lymphoid cell
type I pneumocyte
type II pneumocyte
pancreatic acinar cell
enterocyte of epithelium of large intestine
pancreatic ductal cell
endothelial cell of lymphatic vessel
endothelial cell of vascular tree
capillary endothelial cell
epithelial cell of uterus
intestinal crypt stem cell
connective tissue cell
luminal epithelial cell of mammary gland
luminal cell of prostate epithelium
basal cell of prostate epithelium
keratocyte
myometrial cell
respiratory goblet cell
intermediate monocyte
CD141-positive myeloid dendritic cell
CD1c-positive myeloid dendritic cell
pancreatic stellate cell
adventitial cell
kidney epithelial cell
vein endothelial cell
fibroblast of cardiac tissue
retinal blood vessel endothelial cell
acinar cell of salivary gland
skeletal muscle satellite stem cell
paneth cell of colon
transit amplifying cell of colon
transit amplifying cell of small intestine
intestinal crypt stem cell of large intestine
cardiac endothelial cell
intestinal tuft cell
lung ciliated cell
large intestine goblet cell
enterocyte of epithelium of small intestine
paneth cell of epithelium of small intestine
endothelial cell of hepatic sinusoid
endothelial cell of artery
conjunctival epithelial cell
small intestine goblet cell
bladder urothelial cell
intestinal enteroendocrine cell
lung microvascular endothelial cell
fibroblast of breast
Mueller cell
native cell
BRDT
TXNDC5
CRACR2A
ACADSB
LACTB2
SLC13A1
CXCL3
ANXA3
SCARF1
CDNF
EFCAB2
CETN2
TMCO5A
CRYGD
ATP6V1G2
CABP2
VASH1
DMD
STXBP1
SNAP25
DLG4
RBPMS2
BRSK2
PPP1R14A
AKR1B1
CRYBB1
THTPA
PSTPIP2
NFE2
GMPR
SNCA
CLEC1B
GIPC3
SUSD1
GRAP2
NT5C3A
MPIG6B
ACRBP
GP6
feature_name

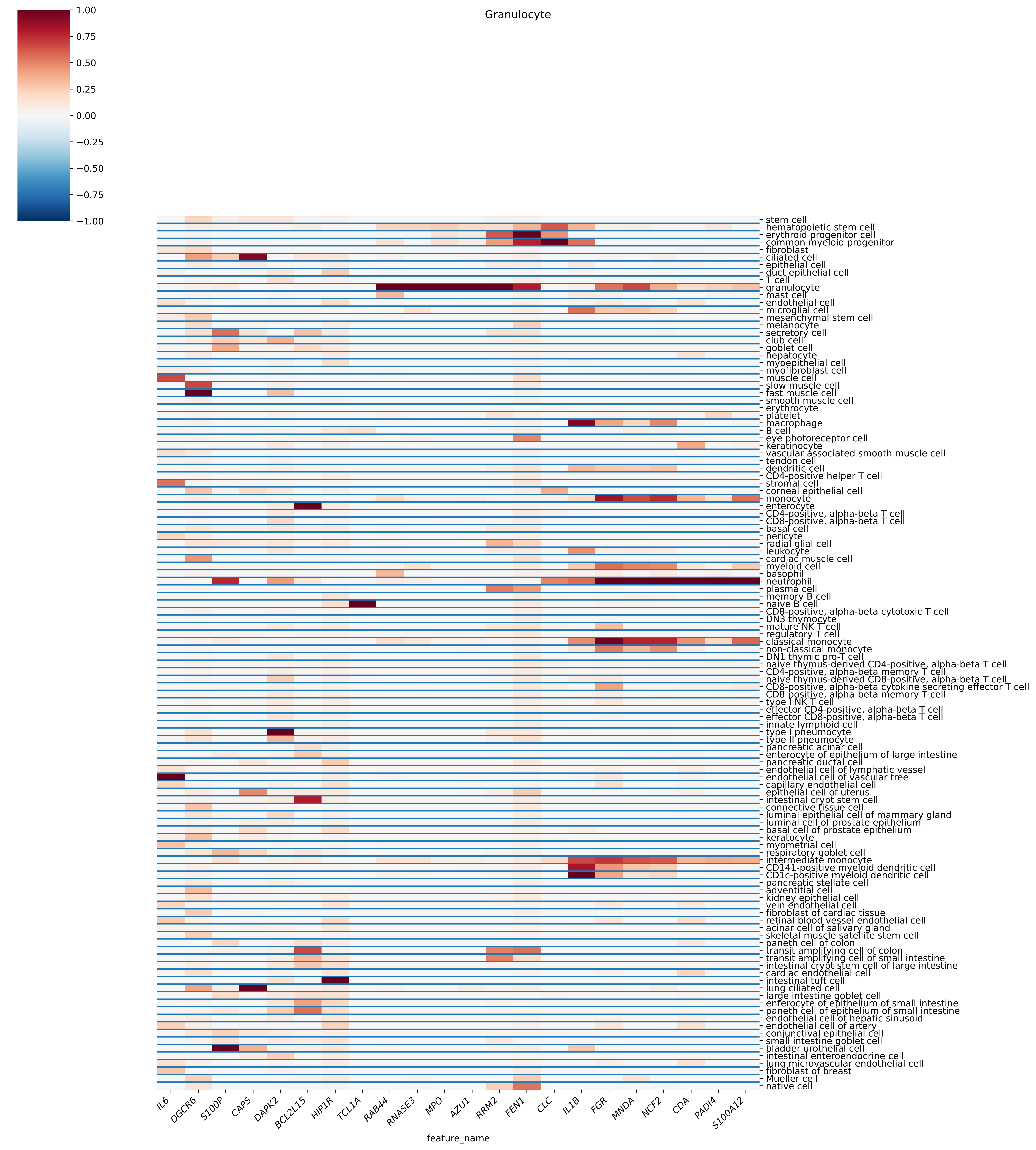

Granulocyte
1.00
0.75
0.50
0.25
0.00
−0.25
−0.50
−0.75
−1.00
stem cell
hematopoietic stem cell
erythroid progenitor cell
common myeloid progenitor
fibroblast
ciliated cell
epithelial cell
duct epithelial cell
T cell
granulocyte
mast cell
endothelial cell
microglial cell
mesenchymal stem cell
melanocyte
secretory cell
club cell
goblet cell
hepatocyte
myoepithelial cell
myofibroblast cell
muscle cell
slow muscle cell
fast muscle cell
smooth muscle cell
erythrocyte
platelet
macrophage
B cell
eye photoreceptor cell
keratinocyte
vascular associated smooth muscle cell
tendon cell
dendritic cell
CD4-positive helper T cell
stromal cell
corneal epithelial cell
monocyte
enterocyte
CD4-positive, alpha-beta T cell
CD8-positive, alpha-beta T cell
basal cell
pericyte
radial glial cell
leukocyte
cardiac muscle cell
myeloid cell
basophil
neutrophil
plasma cell
memory B cell
naive B cell
CD8-positive, alpha-beta cytotoxic T cell
DN3 thymocyte
mature NK T cell
regulatory T cell
classical monocyte
non-classical monocyte
DN1 thymic pro-T cell
naive thymus-derived CD4-positive, alpha-beta T cell
CD4-positive, alpha-beta memory T cell
naive thymus-derived CD8-positive, alpha-beta T cell
CD8-positive, alpha-beta cytokine secreting effector T cell
CD8-positive, alpha-beta memory T cell
type I NK T cell
effector CD4-positive, alpha-beta T cell
effector CD8-positive, alpha-beta T cell
innate lymphoid cell
type I pneumocyte
type II pneumocyte
pancreatic acinar cell
enterocyte of epithelium of large intestine
pancreatic ductal cell
endothelial cell of lymphatic vessel
endothelial cell of vascular tree
capillary endothelial cell
epithelial cell of uterus
intestinal crypt stem cell
connective tissue cell
luminal epithelial cell of mammary gland
luminal cell of prostate epithelium
basal cell of prostate epithelium
keratocyte
myometrial cell
respiratory goblet cell
intermediate monocyte
CD141-positive myeloid dendritic cell
CD1c-positive myeloid dendritic cell
pancreatic stellate cell
adventitial cell
kidney epithelial cell
vein endothelial cell
fibroblast of cardiac tissue
retinal blood vessel endothelial cell
acinar cell of salivary gland
skeletal muscle satellite stem cell
paneth cell of colon
transit amplifying cell of colon
transit amplifying cell of small intestine
intestinal crypt stem cell of large intestine
cardiac endothelial cell
intestinal tuft cell
lung ciliated cell
large intestine goblet cell
enterocyte of epithelium of small intestine
paneth cell of epithelium of small intestine
endothelial cell of hepatic sinusoid
endothelial cell of artery
conjunctival epithelial cell
small intestine goblet cell
bladder urothelial cell
intestinal enteroendocrine cell
lung microvascular endothelial cell
fibroblast of breast
Mueller cell
native cell
IL6
DGCR6
S100P
CAPS
DAPK2
BCL2L15
HIP1R
TCL1A
RAB44
RNASE3
MPO
AZU1
RRM2
FEN1
CLC
IL1B
FGR
MNDA
NCF2
CDA
PADI4
S100A12
feature_name

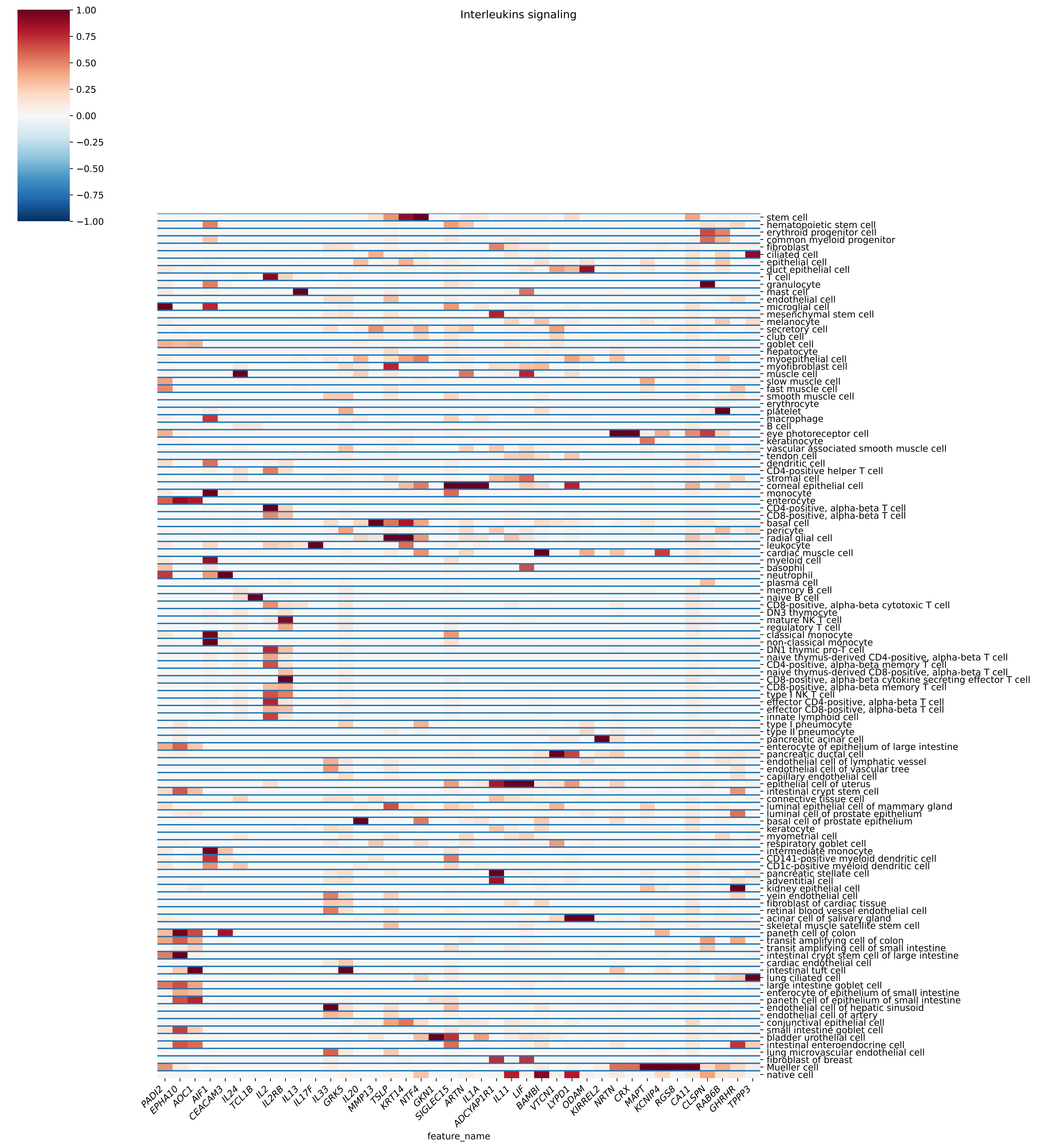

Interleukins signaling
1.00
0.75
0.50
0.25
0.00
−0.25
−0.50
−0.75
−1.00
stem cell
hematopoietic stem cell
erythroid progenitor cell
common myeloid progenitor
fibroblast
ciliated cell
epithelial cell
duct epithelial cell
T cell
granulocyte
mast cell
endothelial cell
microglial cell
mesenchymal stem cell
melanocyte
secretory cell
club cell
goblet cell
hepatocyte
myoepithelial cell
myofibroblast cell
muscle cell
slow muscle cell
fast muscle cell
smooth muscle cell
erythrocyte
platelet
macrophage
B cell
eye photoreceptor cell
keratinocyte
vascular associated smooth muscle cell
tendon cell
dendritic cell
CD4-positive helper T cell
stromal cell
corneal epithelial cell
monocyte
enterocyte
CD4-positive, alpha-beta T cell
CD8-positive, alpha-beta T cell
basal cell
pericyte
radial glial cell
leukocyte
cardiac muscle cell
myeloid cell
basophil
neutrophil
plasma cell
memory B cell
naive B cell
CD8-positive, alpha-beta cytotoxic T cell
DN3 thymocyte
mature NK T cell
regulatory T cell
classical monocyte
non-classical monocyte
DN1 thymic pro-T cell
naive thymus-derived CD4-positive, alpha-beta T cell
CD4-positive, alpha-beta memory T cell
naive thymus-derived CD8-positive, alpha-beta T cell
CD8-positive, alpha-beta cytokine secreting effector T cell
CD8-positive, alpha-beta memory T cell
type I NK T cell
effector CD4-positive, alpha-beta T cell
effector CD8-positive, alpha-beta T cell
innate lymphoid cell
type I pneumocyte
type II pneumocyte
pancreatic acinar cell
enterocyte of epithelium of large intestine
pancreatic ductal cell
endothelial cell of lymphatic vessel
endothelial cell of vascular tree
capillary endothelial cell
epithelial cell of uterus
intestinal crypt stem cell
connective tissue cell
luminal epithelial cell of mammary gland
luminal cell of prostate epithelium
basal cell of prostate epithelium
keratocyte
myometrial cell
respiratory goblet cell
intermediate monocyte
CD141-positive myeloid dendritic cell
CD1c-positive myeloid dendritic cell
pancreatic stellate cell
adventitial cell
kidney epithelial cell
vein endothelial cell
fibroblast of cardiac tissue
retinal blood vessel endothelial cell
acinar cell of salivary gland
skeletal muscle satellite stem cell
paneth cell of colon
transit amplifying cell of colon
transit amplifying cell of small intestine
intestinal crypt stem cell of large intestine
cardiac endothelial cell
intestinal tuft cell
lung ciliated cell
large intestine goblet cell
enterocyte of epithelium of small intestine
paneth cell of epithelium of small intestine
endothelial cell of hepatic sinusoid
endothelial cell of artery
conjunctival epithelial cell
small intestine goblet cell
bladder urothelial cell
intestinal enteroendocrine cell
lung microvascular endothelial cell
fibroblast of breast
Mueller cell
native cell
PADI2
EPHA10
AOC1
AIF1
CEACAM3
IL24
TCL1B
IL2
IL2RB
IL13
IL17F
IL33
GRK5
IL20
MMP13
TSLP
KRT14
NTF4
GKN1
SIGLEC15
ARTN
IL1A
ADCYAP1R1
IL11
LIF
BAMBI
VTCN1
LYPD1
ODAM
KIRREL2
NRTN
CRX
MAPT
KCNIP4
RGS8
CA11
CLSPN
RAB6B
GHRHR
TPPP3
feature_name

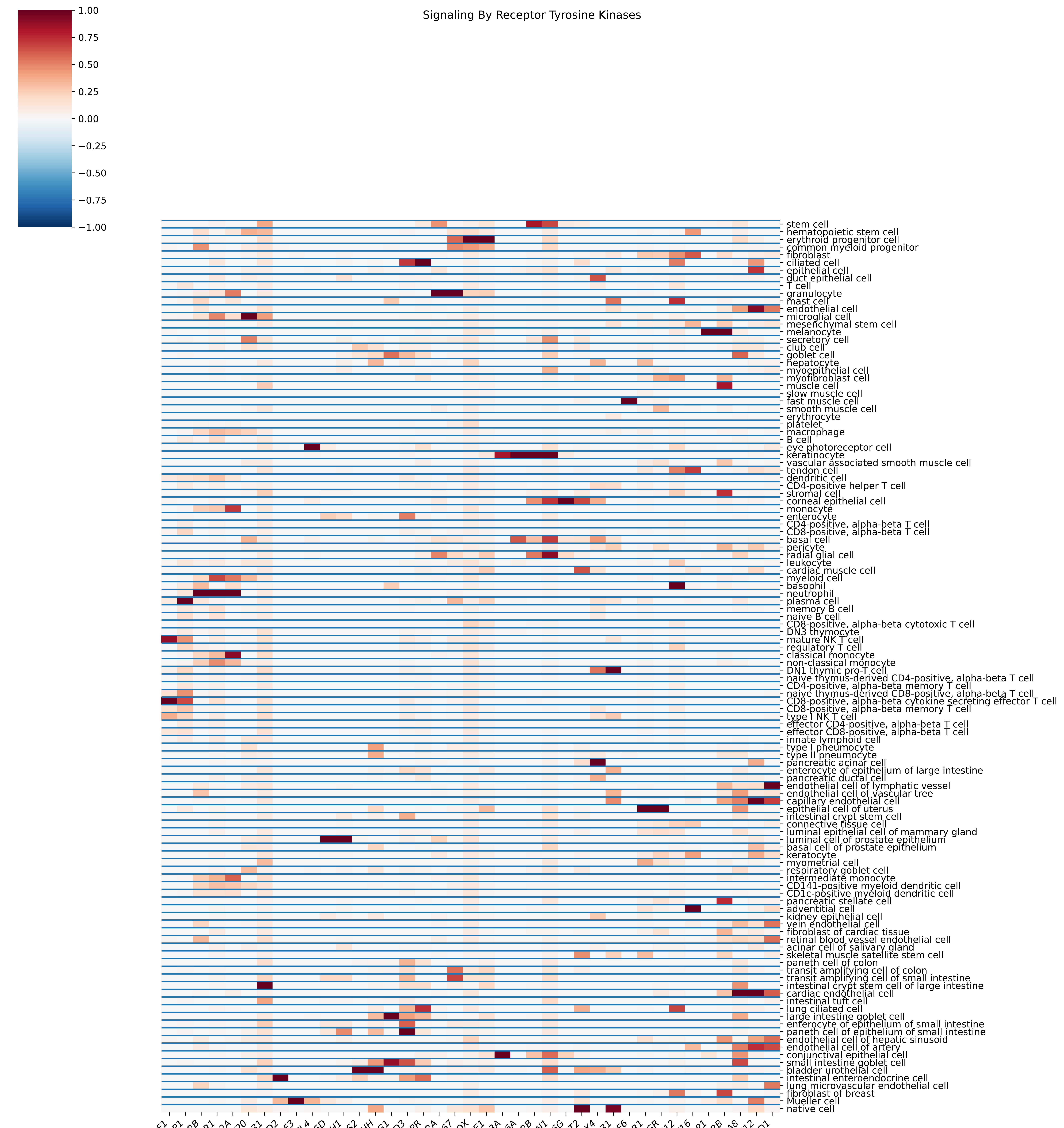

Signaling By Receptor Tyrosine Kinases
1.00
0.75
0.50
0.25
0.00
−0.25
−0.50
−0.75
−1.00
stem cell
hematopoietic stem cell
erythroid progenitor cell
common myeloid progenitor
fibroblast
ciliated cell
epithelial cell
duct epithelial cell
T cell
granulocyte
mast cell
endothelial cell
microglial cell
mesenchymal stem cell
melanocyte
secretory cell
club cell
goblet cell
hepatocyte
myoepithelial cell
myofibroblast cell
muscle cell
slow muscle cell
fast muscle cell
smooth muscle cell
erythrocyte
platelet
macrophage
B cell
eye photoreceptor cell
keratinocyte
vascular associated smooth muscle cell
tendon cell
dendritic cell
CD4-positive helper T cell
stromal cell
corneal epithelial cell
monocyte
enterocyte
CD4-positive, alpha-beta T cell
CD8-positive, alpha-beta T cell
basal cell
pericyte
radial glial cell
leukocyte
cardiac muscle cell
myeloid cell
basophil
neutrophil
plasma cell
memory B cell
naive B cell
CD8-positive, alpha-beta cytotoxic T cell
DN3 thymocyte
mature NK T cell
regulatory T cell
classical monocyte
non-classical monocyte
DN1 thymic pro-T cell
naive thymus-derived CD4-positive, alpha-beta T cell
CD4-positive, alpha-beta memory T cell
naive thymus-derived CD8-positive, alpha-beta T cell
CD8-positive, alpha-beta cytokine secreting effector T cell
CD8-positive, alpha-beta memory T cell
type I NK T cell
effector CD4-positive, alpha-beta T cell
effector CD8-positive, alpha-beta T cell
innate lymphoid cell
type I pneumocyte
type II pneumocyte
pancreatic acinar cell
enterocyte of epithelium of large intestine
pancreatic ductal cell
endothelial cell of lymphatic vessel
endothelial cell of vascular tree
capillary endothelial cell
epithelial cell of uterus
intestinal crypt stem cell
connective tissue cell
luminal epithelial cell of mammary gland
luminal cell of prostate epithelium
basal cell of prostate epithelium
keratocyte
myometrial cell
respiratory goblet cell
intermediate monocyte
CD141-positive myeloid dendritic cell
CD1c-positive myeloid dendritic cell
pancreatic stellate cell
adventitial cell
kidney epithelial cell
vein endothelial cell
fibroblast of cardiac tissue
retinal blood vessel endothelial cell
acinar cell of salivary gland
skeletal muscle satellite stem cell
paneth cell of colon
transit amplifying cell of colon
transit amplifying cell of small intestine
intestinal crypt stem cell of large intestine
cardiac endothelial cell
intestinal tuft cell
lung ciliated cell
large intestine goblet cell
enterocyte of epithelium of small intestine
paneth cell of epithelium of small intestine
endothelial cell of hepatic sinusoid
endothelial cell of artery
conjunctival epithelial cell
small intestine goblet cell
bladder urothelial cell
intestinal enteroendocrine cell
lung microvascular endothelial cell
fibroblast of breast
Mueller cell
native cell
KLRF1
ZBP1
CSF2RB
TLR1
CLEC12A
FGF20
RNF31
GAD2
FGF3
IGFL4
UNC5D
FOLH1
NPHS2
SHH
TPSG1
SMPD3
GIPR
IL31RA
MKI67
CPOX
CCNE1
DEFB103A
IL36A
IL20RB
NECTIN1
IL36G
HS6ST2
DDX4
IL31
FGF6
ESR1
PGR
ADAM12
FGF16
TYRP1
EDNRB
CA8
FGF12
THSD1
feature_name

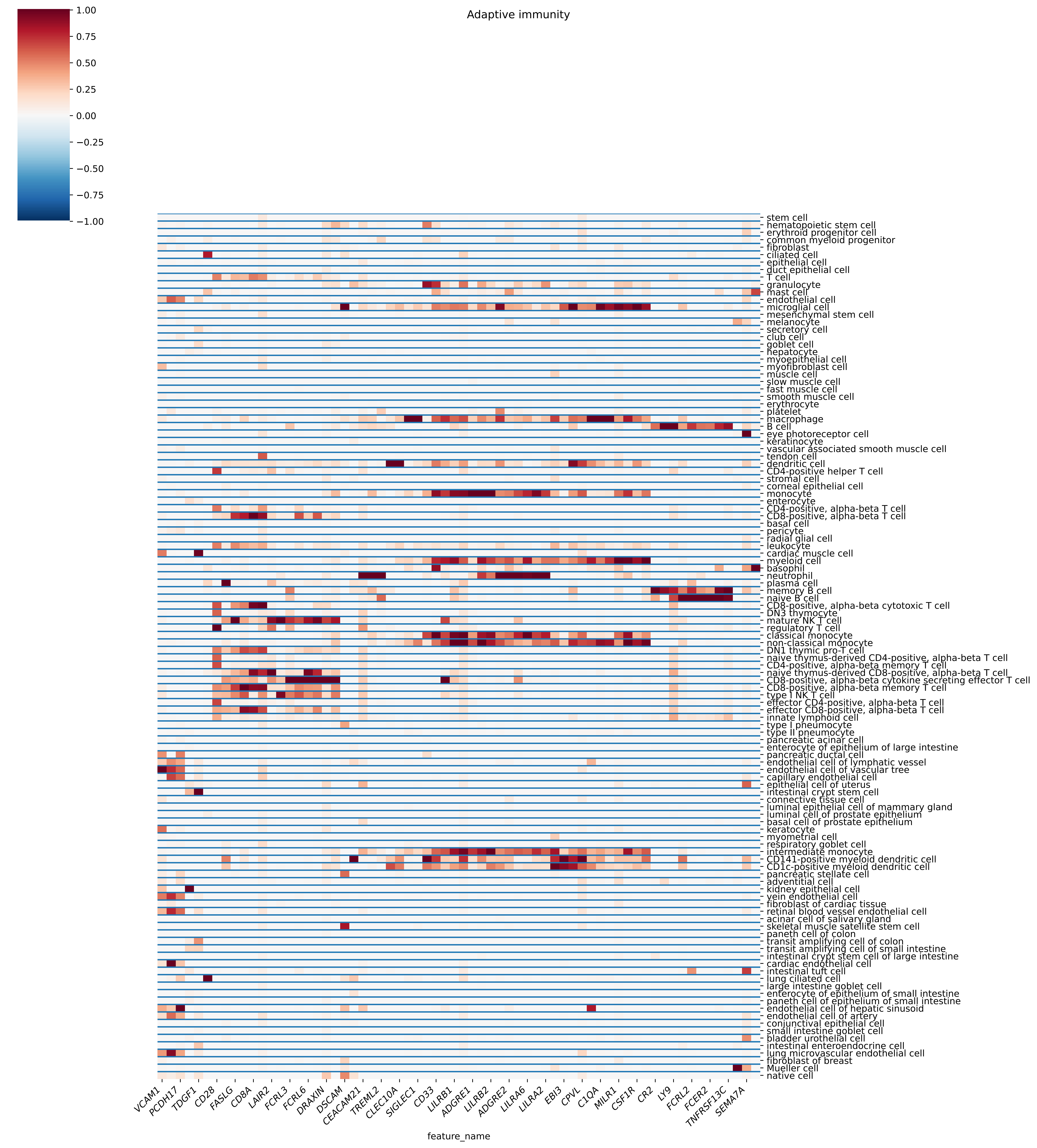

Adaptive immunity
1.00
0.75
0.50
0.25
0.00
−0.25
−0.50
−0.75
−1.00
stem cell
hematopoietic stem cell
erythroid progenitor cell
common myeloid progenitor
fibroblast
ciliated cell
epithelial cell
duct epithelial cell
T cell
granulocyte
mast cell
endothelial cell
microglial cell
mesenchymal stem cell
melanocyte
secretory cell
club cell
goblet cell
hepatocyte
myoepithelial cell
myofibroblast cell
muscle cell
slow muscle cell
fast muscle cell
smooth muscle cell
erythrocyte
platelet
macrophage
B cell
eye photoreceptor cell
keratinocyte
vascular associated smooth muscle cell
tendon cell
dendritic cell
CD4-positive helper T cell
stromal cell
corneal epithelial cell
monocyte
enterocyte
CD4-positive, alpha-beta T cell
CD8-positive, alpha-beta T cell
basal cell
pericyte
radial glial cell
leukocyte
cardiac muscle cell
myeloid cell
basophil
neutrophil
plasma cell
memory B cell
naive B cell
CD8-positive, alpha-beta cytotoxic T cell
DN3 thymocyte
mature NK T cell
regulatory T cell
classical monocyte
non-classical monocyte
DN1 thymic pro-T cell
naive thymus-derived CD4-positive, alpha-beta T cell
CD4-positive, alpha-beta memory T cell
naive thymus-derived CD8-positive, alpha-beta T cell
CD8-positive, alpha-beta cytokine secreting effector T cell
CD8-positive, alpha-beta memory T cell
type I NK T cell
effector CD4-positive, alpha-beta T cell
effector CD8-positive, alpha-beta T cell
innate lymphoid cell
type I pneumocyte
type II pneumocyte
pancreatic acinar cell
enterocyte of epithelium of large intestine
pancreatic ductal cell
endothelial cell of lymphatic vessel
endothelial cell of vascular tree
capillary endothelial cell
epithelial cell of uterus
intestinal crypt stem cell
connective tissue cell
luminal epithelial cell of mammary gland
luminal cell of prostate epithelium
basal cell of prostate epithelium
keratocyte
myometrial cell
respiratory goblet cell
intermediate monocyte
CD141-positive myeloid dendritic cell
CD1c-positive myeloid dendritic cell
pancreatic stellate cell
adventitial cell
kidney epithelial cell
vein endothelial cell
fibroblast of cardiac tissue
retinal blood vessel endothelial cell
acinar cell of salivary gland
skeletal muscle satellite stem cell
paneth cell of colon
transit amplifying cell of colon
transit amplifying cell of small intestine
intestinal crypt stem cell of large intestine
cardiac endothelial cell
intestinal tuft cell
lung ciliated cell
large intestine goblet cell
enterocyte of epithelium of small intestine
paneth cell of epithelium of small intestine
endothelial cell of hepatic sinusoid
endothelial cell of artery
conjunctival epithelial cell
small intestine goblet cell
bladder urothelial cell
intestinal enteroendocrine cell
lung microvascular endothelial cell
fibroblast of breast
Mueller cell
native cell
VCAM1
PCDH17
TDGF1
CD28
FASLG
CD8A
LAIR2
FCRL3
FCRL6
DRAXIN
DSCAM
CEACAM21
TREML2
CLEC10A
SIGLEC1
CD33
LILRB1
ADGRE1
LILRB2
ADGRE2
LILRA6
LILRA2
EBI3
CPVL
C1QA
MILR1
CSF1R
CR2
LY9
FCRL2
FCER2
TNFRSF13C
SEMA7A
feature_name

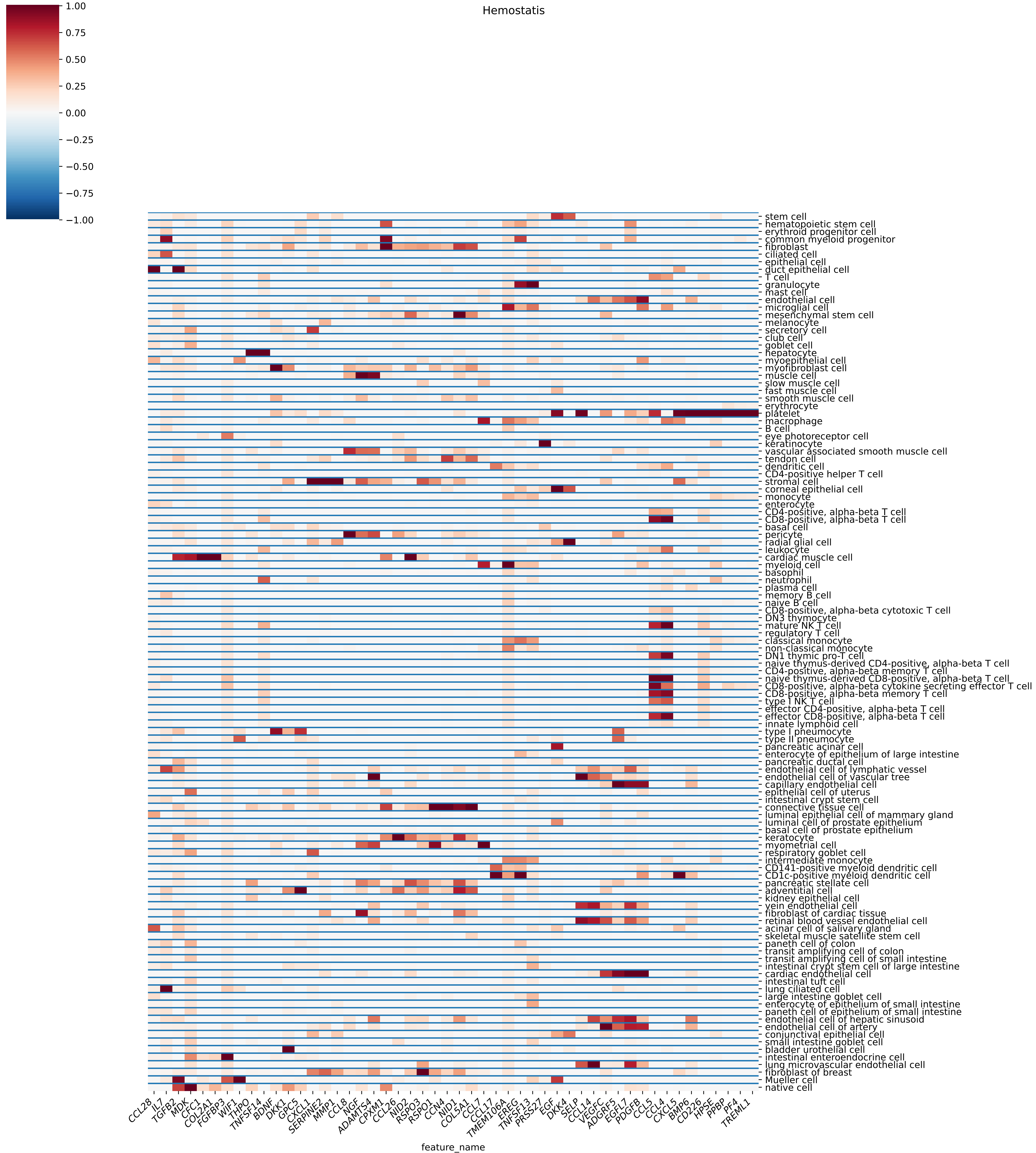
Hemostatis
1.00
0.75
0.50
0.25
0.00
−0.25
−0.50
−0.75
−1.00
stem cell
hematopoietic stem cell
erythroid progenitor cell
common myeloid progenitor
fibroblast
ciliated cell
epithelial cell
duct epithelial cell
T cell
granulocyte
mast cell
endothelial cell
microglial cell
mesenchymal stem cell
melanocyte
secretory cell
club cell
goblet cell
hepatocyte
myoepithelial cell
myofibroblast cell
muscle cell
slow muscle cell
fast muscle cell
smooth muscle cell
erythrocyte
platelet
macrophage
B cell
eye photoreceptor cell
keratinocyte
vascular associated smooth muscle cell
tendon cell
dendritic cell
CD4-positive helper T cell
stromal cell
corneal epithelial cell
monocyte
enterocyte
CD4-positive, alpha-beta T cell
CD8-positive, alpha-beta T cell
basal cell
pericyte
radial glial cell
leukocyte
cardiac muscle cell
myeloid cell
basophil
neutrophil
plasma cell
memory B cell
naive B cell
CD8-positive, alpha-beta cytotoxic T cell
DN3 thymocyte
mature NK T cell
regulatory T cell
classical monocyte
non-classical monocyte
DN1 thymic pro-T cell
naive thymus-derived CD4-positive, alpha-beta T cell
CD4-positive, alpha-beta memory T cell
naive thymus-derived CD8-positive, alpha-beta T cell
CD8-positive, alpha-beta cytokine secreting effector T cell
CD8-positive, alpha-beta memory T cell
type I NK T cell
effector CD4-positive, alpha-beta T cell
effector CD8-positive, alpha-beta T cell
innate lymphoid cell
type I pneumocyte
type II pneumocyte
pancreatic acinar cell
enterocyte of epithelium of large intestine
pancreatic ductal cell
endothelial cell of lymphatic vessel
endothelial cell of vascular tree
capillary endothelial cell
epithelial cell of uterus
intestinal crypt stem cell
connective tissue cell
luminal epithelial cell of mammary gland
luminal cell of prostate epithelium
basal cell of prostate epithelium
keratocyte
myometrial cell
respiratory goblet cell
intermediate monocyte
CD141-positive myeloid dendritic cell
CD1c-positive myeloid dendritic cell
pancreatic stellate cell
adventitial cell
kidney epithelial cell
vein endothelial cell
fibroblast of cardiac tissue
retinal blood vessel endothelial cell
acinar cell of salivary gland
skeletal muscle satellite stem cell
paneth cell of colon
transit amplifying cell of colon
transit amplifying cell of small intestine
intestinal crypt stem cell of large intestine
cardiac endothelial cell
intestinal tuft cell
lung ciliated cell
large intestine goblet cell
enterocyte of epithelium of small intestine
paneth cell of epithelium of small intestine
endothelial cell of hepatic sinusoid
endothelial cell of artery
conjunctival epithelial cell
small intestine goblet cell
bladder urothelial cell
intestinal enteroendocrine cell
lung microvascular endothelial cell
fibroblast of breast
Mueller cell
native cell
CCL28
IL7
TGFB2
MDK
CFC1
COL2A1
FGFBP3
WIF1
THPO
TNFSF14
BDNF
DKK1
GPC5
CXCL1
SERPINE2
MMP1
CCL8
NGF
ADAMTS4
CPXM1
CCL26
NID2
RSPO3
RSPO1
CCN4
NID1
COL5A1
CCL7
CCL17
TMEM106A
EREG
TNFSF13
PRSS27
EGF
DKK4
SELP
CCL14
VEGFC
ADGRF5
EGFL7
PDGFB
CCL5
CCL4
CXCL5
BMP6
CD226
HPSE
PPBP
PF4
TREML1
feature_name

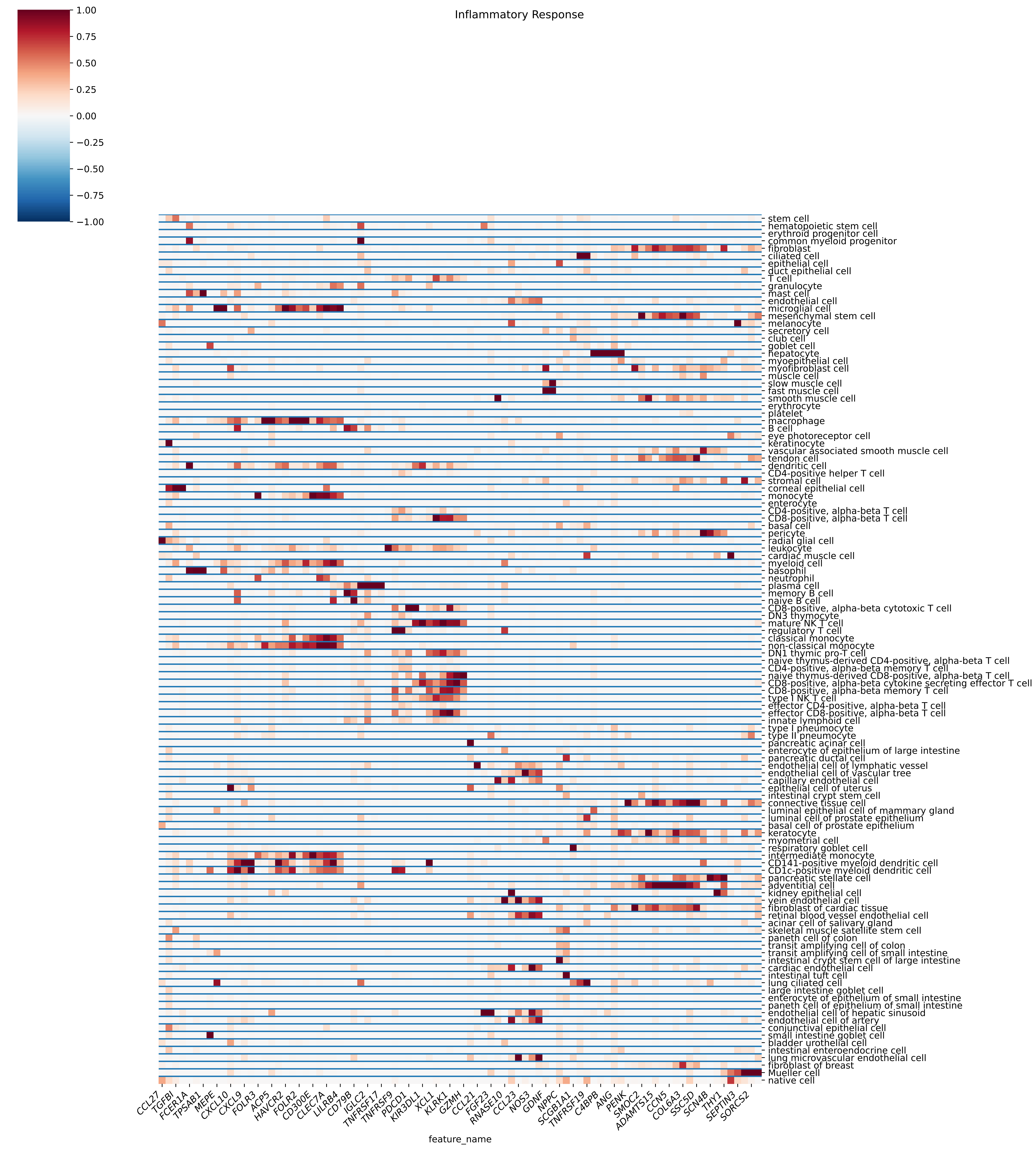
Inflammatory Response
1.00
0.75
0.50
0.25
0.00
−0.25
−0.50
−0.75
−1.00
feature_name
CCL27
TGFBI
FCER1A
TPSAB1
MEPE
CXCL10
CXCL9
FOLR3
ACP5
HAVCR2
FOLR2
CD300E
CLEC7A
LILRB4
CD79B
IGLC2
TNFRSF17
TNFRSF9
PDCD1
KIR3DL1
XCL1
KLRK1
GZMH
CCL21
FGF23
RNASE10
CCL23
NOS3
GDNF
NPPC
SCGB1A1
TNFRSF19
C4BPB
ANG
PENK
SMOC2
ADAMTS15
CCN5
COL6A3
SSC5D
SCN4B
THY1
SEPTIN3
SORCS2
stem cell
hematopoietic stem cell
erythroid progenitor cell
common myeloid progenitor
fibroblast
ciliated cell
epithelial cell
duct epithelial cell
T cell
granulocyte
mast cell
endothelial cell
microglial cell
mesenchymal stem cell
melanocyte
secretory cell
club cell
goblet cell
hepatocyte
myoepithelial cell
myofibroblast cell
muscle cell
slow muscle cell
fast muscle cell
smooth muscle cell
erythrocyte
platelet
macrophage
B cell
eye photoreceptor cell
keratinocyte
vascular associated smooth muscle cell
tendon cell
dendritic cell
CD4-positive helper T cell
stromal cell
corneal epithelial cell
monocyte
enterocyte
CD4-positive, alpha-beta T cell
CD8-positive, alpha-beta T cell
basal cell
pericyte
radial glial cell
leukocyte
cardiac muscle cell
myeloid cell
basophil
neutrophil
plasma cell
memory B cell
naive B cell
CD8-positive, alpha-beta cytotoxic T cell
DN3 thymocyte
mature NK T cell
regulatory T cell
classical monocyte
non-classical monocyte
DN1 thymic pro-T cell
naive thymus-derived CD4-positive, alpha-beta T cell
CD4-positive, alpha-beta memory T cell
naive thymus-derived CD8-positive, alpha-beta T cell
CD8-positive, alpha-beta cytokine secreting effector T cell
CD8-positive, alpha-beta memory T cell
type I NK T cell
effector CD4-positive, alpha-beta T cell
effector CD8-positive, alpha-beta T cell
innate lymphoid cell
type I pneumocyte
type II pneumocyte
pancreatic acinar cell
enterocyte of epithelium of large intestine
pancreatic ductal cell
endothelial cell of lymphatic vessel
endothelial cell of vascular tree
capillary endothelial cell
epithelial cell of uterus
intestinal crypt stem cell
connective tissue cell
luminal epithelial cell of mammary gland
luminal cell of prostate epithelium
basal cell of prostate epithelium
keratocyte
myometrial cell
respiratory goblet cell
intermediate monocyte
CD141-positive myeloid dendritic cell
CD1c-positive myeloid dendritic cell
pancreatic stellate cell
adventitial cell
kidney epithelial cell
vein endothelial cell
fibroblast of cardiac tissue
retinal blood vessel endothelial cell
acinar cell of salivary gland
skeletal muscle satellite stem cell
paneth cell of colon
transit amplifying cell of colon
transit amplifying cell of small intestine
intestinal crypt stem cell of large intestine
cardiac endothelial cell
intestinal tuft cell
lung ciliated cell
large intestine goblet cell
enterocyte of epithelium of small intestine
paneth cell of epithelium of small intestine
endothelial cell of hepatic sinusoid
endothelial cell of artery
conjunctival epithelial cell
small intestine goblet cell
bladder urothelial cell
intestinal enteroendocrine cell
lung microvascular endothelial cell
fibroblast of breast
Mueller cell
native cell

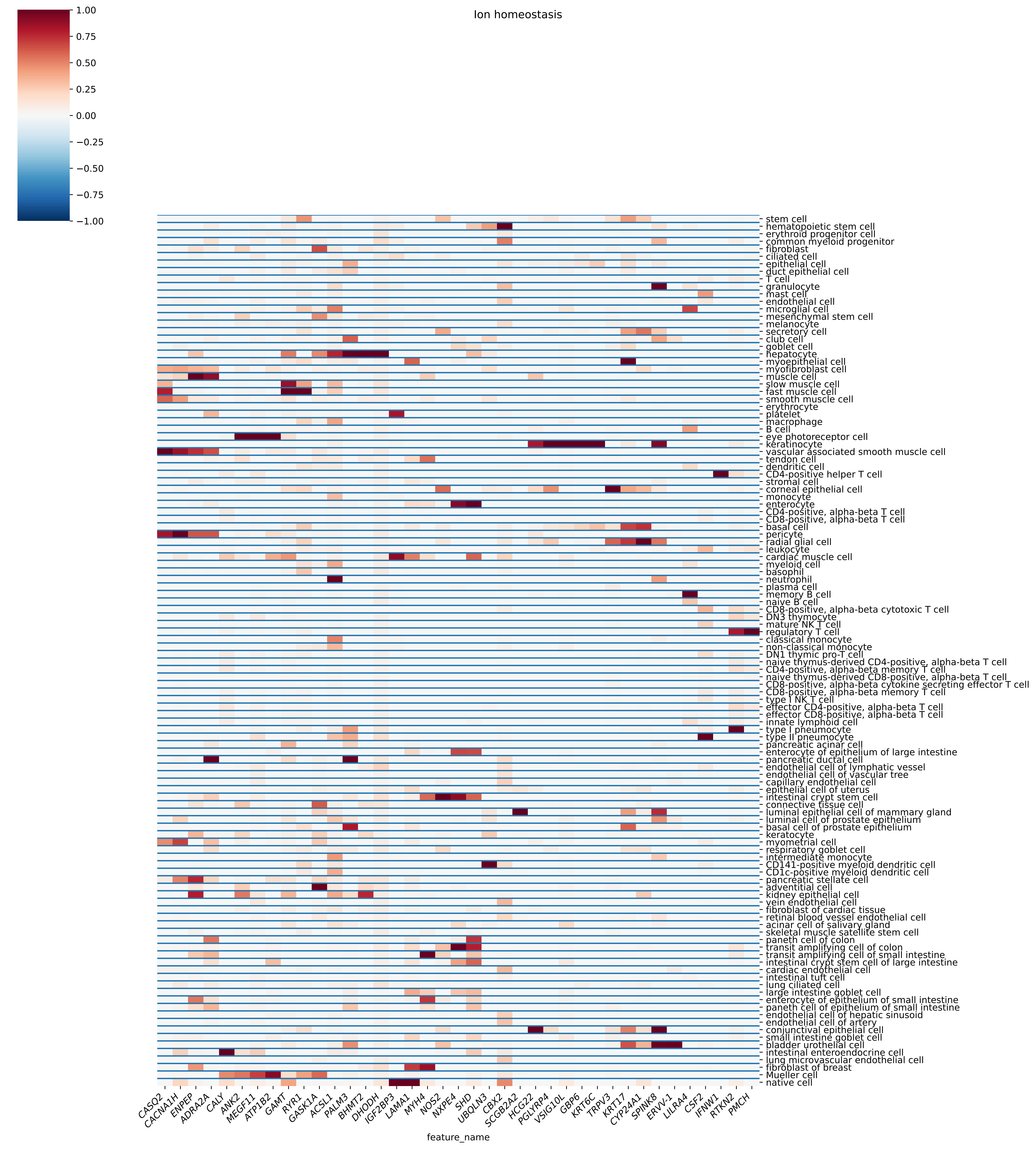

Ion homeostasis
1.00
0.75
0.50
0.25
0.00
−0.25
−0.50
−0.75
−1.00
stem cell
hematopoietic stem cell
erythroid progenitor cell
common myeloid progenitor
fibroblast
ciliated cell
epithelial cell
duct epithelial cell
T cell
granulocyte
mast cell
endothelial cell
microglial cell
mesenchymal stem cell
melanocyte
secretory cell
club cell
goblet cell
hepatocyte
myoepithelial cell
myofibroblast cell
muscle cell
slow muscle cell
fast muscle cell
smooth muscle cell
erythrocyte
platelet
macrophage
B cell
eye photoreceptor cell
keratinocyte
vascular associated smooth muscle cell
tendon cell
dendritic cell
CD4-positive helper T cell
stromal cell
corneal epithelial cell
monocyte
enterocyte
CD4-positive, alpha-beta T cell
CD8-positive, alpha-beta T cell
basal cell
pericyte
radial glial cell
leukocyte
cardiac muscle cell
myeloid cell
basophil
neutrophil
plasma cell
memory B cell
naive B cell
CD8-positive, alpha-beta cytotoxic T cell
DN3 thymocyte
mature NK T cell
regulatory T cell
classical monocyte
non-classical monocyte
DN1 thymic pro-T cell
naive thymus-derived CD4-positive, alpha-beta T cell
CD4-positive, alpha-beta memory T cell
naive thymus-derived CD8-positive, alpha-beta T cell
CD8-positive, alpha-beta cytokine secreting effector T cell
CD8-positive, alpha-beta memory T cell
type I NK T cell
effector CD4-positive, alpha-beta T cell
effector CD8-positive, alpha-beta T cell
innate lymphoid cell
type I pneumocyte
type II pneumocyte
pancreatic acinar cell
enterocyte of epithelium of large intestine
pancreatic ductal cell
endothelial cell of lymphatic vessel
endothelial cell of vascular tree
capillary endothelial cell
epithelial cell of uterus
intestinal crypt stem cell
connective tissue cell
luminal epithelial cell of mammary gland
luminal cell of prostate epithelium
basal cell of prostate epithelium
keratocyte
myometrial cell
respiratory goblet cell
intermediate monocyte
CD141-positive myeloid dendritic cell
CD1c-positive myeloid dendritic cell
pancreatic stellate cell
adventitial cell
kidney epithelial cell
vein endothelial cell
fibroblast of cardiac tissue
retinal blood vessel endothelial cell
acinar cell of salivary gland
skeletal muscle satellite stem cell
paneth cell of colon
transit amplifying cell of colon
transit amplifying cell of small intestine
intestinal crypt stem cell of large intestine
cardiac endothelial cell
intestinal tuft cell
lung ciliated cell
large intestine goblet cell
enterocyte of epithelium of small intestine
paneth cell of epithelium of small intestine
endothelial cell of hepatic sinusoid
endothelial cell of artery
conjunctival epithelial cell
small intestine goblet cell
bladder urothelial cell
intestinal enteroendocrine cell
lung microvascular endothelial cell
fibroblast of breast
Mueller cell
native cell
CASQ2
CACNA1H
ENPEP
ADRA2A
CALY
ANK2
MEGF11
ATP1B2
GAMT
RYR1
GASK1A
ACSL1
PALM3
BHMT2
DHODH
IGF2BP3
LAMA1
MYH4
NOS2
NXPE4
SHD
UBQLN3
CBX2
SCGB2A2
HCG22
PGLYRP4
VSIG10L
GBP6
KRT6C
TRPV3
KRT17
CYP24A1
SPINK8
ERVV-1
LILRA4
CSF2
IFNW1
RTKN2
PMCH
feature_name

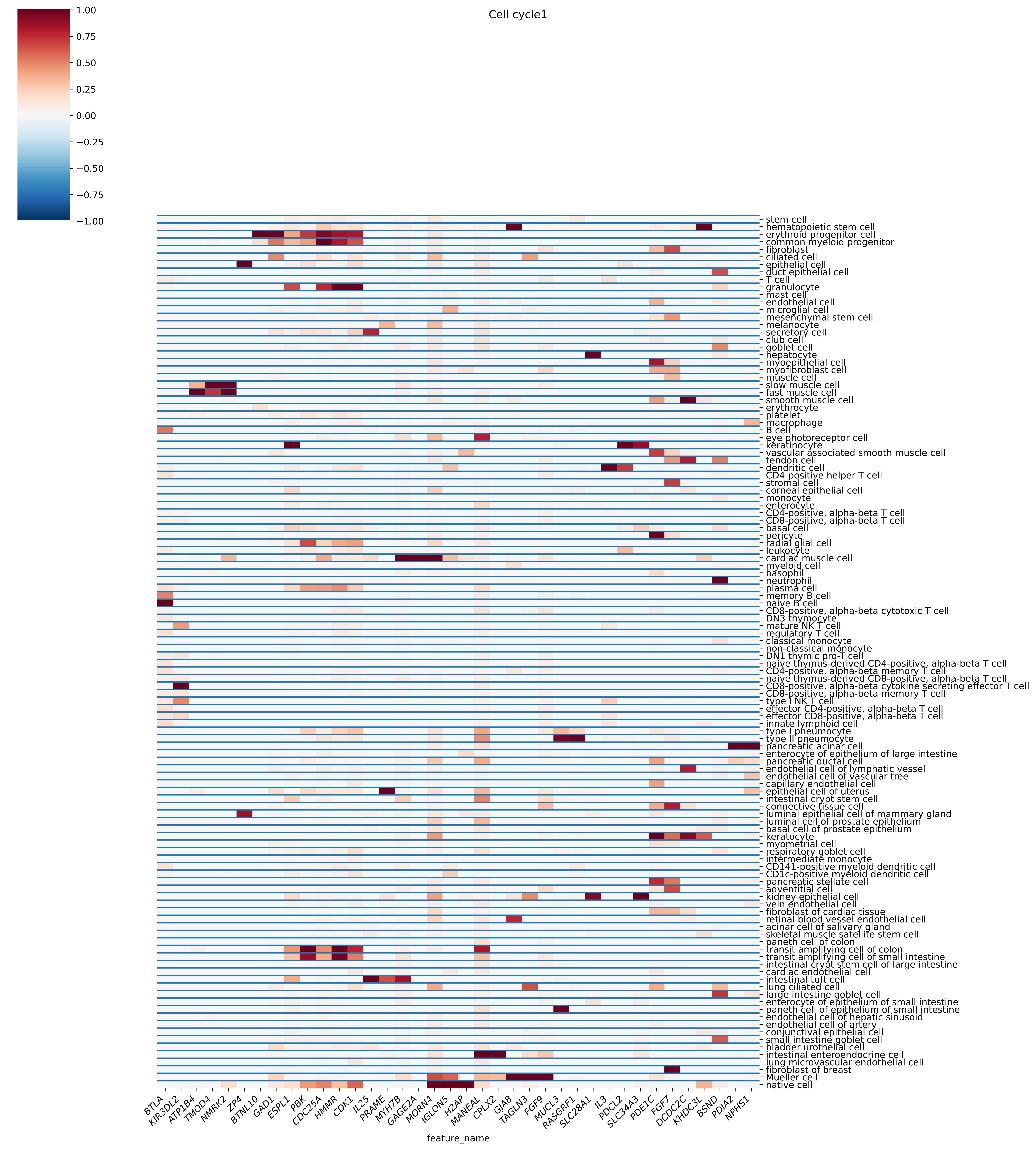

Cell cycle1
1.00
0.75
0.50
0.25
0.00
−0.25
−0.50
−0.75
−1.00
stem cell
hematopoietic stem cell
erythroid progenitor cell
common myeloid progenitor
fibroblast
ciliated cell
epithelial cell
duct epithelial cell
T cell
granulocyte
mast cell
endothelial cell
microglial cell
mesenchymal stem cell
melanocyte
secretory cell
club cell
goblet cell
hepatocyte
myoepithelial cell
myofibroblast cell
muscle cell
slow muscle cell
fast muscle cell
smooth muscle cell
erythrocyte
platelet
macrophage
B cell
eye photoreceptor cell
keratinocyte
vascular associated smooth muscle cell
tendon cell
dendritic cell
CD4-positive helper T cell
stromal cell
corneal epithelial cell
monocyte
enterocyte
CD4-positive, alpha-beta T cell
CD8-positive, alpha-beta T cell
basal cell
pericyte
radial glial cell
leukocyte
cardiac muscle cell
myeloid cell
basophil
neutrophil
plasma cell
memory B cell
naive B cell
CD8-positive, alpha-beta cytotoxic T cell
DN3 thymocyte
mature NK T cell
regulatory T cell
classical monocyte
non-classical monocyte
DN1 thymic pro-T cell
naive thymus-derived CD4-positive, alpha-beta T cell
CD4-positive, alpha-beta memory T cell
naive thymus-derived CD8-positive, alpha-beta T cell
CD8-positive, alpha-beta cytokine secreting effector T cell
CD8-positive, alpha-beta memory T cell
type I NK T cell
effector CD4-positive, alpha-beta T cell
effector CD8-positive, alpha-beta T cell
innate lymphoid cell
type I pneumocyte
type II pneumocyte
pancreatic acinar cell
enterocyte of epithelium of large intestine
pancreatic ductal cell
endothelial cell of lymphatic vessel
endothelial cell of vascular tree
capillary endothelial cell
epithelial cell of uterus
intestinal crypt stem cell
connective tissue cell
luminal epithelial cell of mammary gland
luminal cell of prostate epithelium
basal cell of prostate epithelium
keratocyte
myometrial cell
respiratory goblet cell
intermediate monocyte
CD141-positive myeloid dendritic cell
CD1c-positive myeloid dendritic cell
pancreatic stellate cell
adventitial cell
kidney epithelial cell
vein endothelial cell
fibroblast of cardiac tissue
retinal blood vessel endothelial cell
acinar cell of salivary gland
skeletal muscle satellite stem cell
paneth cell of colon
transit amplifying cell of colon
transit amplifying cell of small intestine
intestinal crypt stem cell of large intestine
cardiac endothelial cell
intestinal tuft cell
lung ciliated cell
large intestine goblet cell
enterocyte of epithelium of small intestine
paneth cell of epithelium of small intestine
endothelial cell of hepatic sinusoid
endothelial cell of artery
conjunctival epithelial cell
small intestine goblet cell
bladder urothelial cell
intestinal enteroendocrine cell
lung microvascular endothelial cell
fibroblast of breast
Mueller cell
native cell
BTLA
KIR3DL2
ATP1B4
TMOD4
NMRK2
ZP4
BTNL10
GAD1
ESPL1
PBK
CDC25A
HMMR
CDK1
IL25
PRAME
MYH7B
GAGE2A
MORN4
IGLON5
H2AP
MANEAL
CPLX2
GJA8
TAGLN3
FGF9
MUCL3
RASGRF1
SLC28A1
IL3
PDCL2
SLC34A3
PDE1C
FGF7
DCDC2C
KHDC3L
BSND
PDIA2
NPHS1
feature_name

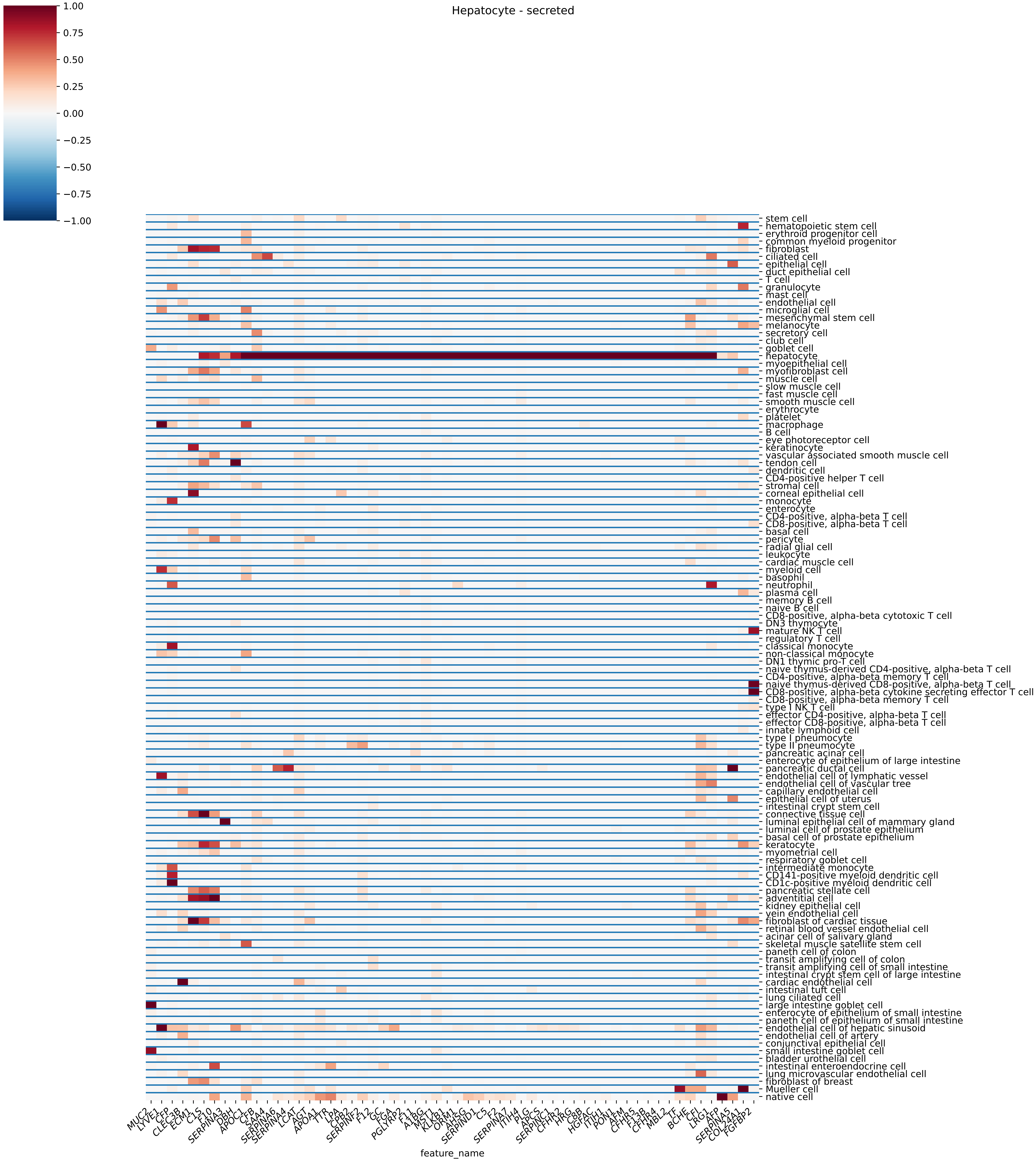
Hepatocyte - secreted
1.00
0.75
0.50
0.25
0.00
−0.25
−0.50
−0.75
−1.00
stem cell
hematopoietic stem cell
erythroid progenitor cell
common myeloid progenitor
fibroblast
ciliated cell
epithelial cell
duct epithelial cell
T cell
granulocyte
mast cell
endothelial cell
microglial cell
mesenchymal stem cell
melanocyte
secretory cell
club cell
goblet cell
hepatocyte
myoepithelial cell
myofibroblast cell
muscle cell
slow muscle cell
fast muscle cell
smooth muscle cell
erythrocyte
platelet
macrophage
B cell
eye photoreceptor cell
keratinocyte
vascular associated smooth muscle cell
tendon cell
dendritic cell
CD4-positive helper T cell
stromal cell
corneal epithelial cell
monocyte
enterocyte
CD4-positive, alpha-beta T cell
CD8-positive, alpha-beta T cell
basal cell
pericyte
radial glial cell
leukocyte
cardiac muscle cell
myeloid cell
basophil
neutrophil
plasma cell
memory B cell
naive B cell
CD8-positive, alpha-beta cytotoxic T cell
DN3 thymocyte
mature NK T cell
regulatory T cell
classical monocyte
non-classical monocyte
DN1 thymic pro-T cell
naive thymus-derived CD4-positive, alpha-beta T cell
CD4-positive, alpha-beta memory T cell
naive thymus-derived CD8-positive, alpha-beta T cell
CD8-positive, alpha-beta cytokine secreting effector T cell
CD8-positive, alpha-beta memory T cell
type I NK T cell
effector CD4-positive, alpha-beta T cell
effector CD8-positive, alpha-beta T cell
innate lymphoid cell
type I pneumocyte
type II pneumocyte
pancreatic acinar cell
enterocyte of epithelium of large intestine
pancreatic ductal cell
endothelial cell of lymphatic vessel
endothelial cell of vascular tree
capillary endothelial cell
epithelial cell of uterus
intestinal crypt stem cell
connective tissue cell
luminal epithelial cell of mammary gland
luminal cell of prostate epithelium
basal cell of prostate epithelium
keratocyte
myometrial cell
respiratory goblet cell
intermediate monocyte
CD141-positive myeloid dendritic cell
CD1c-positive myeloid dendritic cell
pancreatic stellate cell
adventitial cell
kidney epithelial cell
vein endothelial cell
fibroblast of cardiac tissue
retinal blood vessel endothelial cell
acinar cell of salivary gland
skeletal muscle satellite stem cell
paneth cell of colon
transit amplifying cell of colon
transit amplifying cell of small intestine
intestinal crypt stem cell of large intestine
cardiac endothelial cell
intestinal tuft cell
lung ciliated cell
large intestine goblet cell
enterocyte of epithelium of small intestine
paneth cell of epithelium of small intestine
endothelial cell of hepatic sinusoid
endothelial cell of artery
conjunctival epithelial cell
small intestine goblet cell
bladder urothelial cell
intestinal enteroendocrine cell
lung microvascular endothelial cell
fibroblast of breast
Mueller cell
native cell
MUC2
LYVE1
CFP
CLEC3B
ECM1
C1S
F10
SERPINA3
DBH
APOC1
CFB
SAA4
SERPINA6
SERPINA4
LCAT
AGT
APOA1
TTR
LPA
CPB2
SERPINF2
F12
GC
FGA
PGLYRP2
F11
A1BG
MST1
KLKB1
ORM1
AHSG
SERPIND1
C5
F2
SERPINA7
ITIH4
PLG
APCS
SERPINC1
CFHR2
HRG
C8B
HGFAC
ITIH1
PON1
AFM
CFHR5
F13B
CFHR4
MBL2
TF
BCHE
CFI
LRG1
AFP
SERPINA5
COL24A1
FGFBP2
feature_name

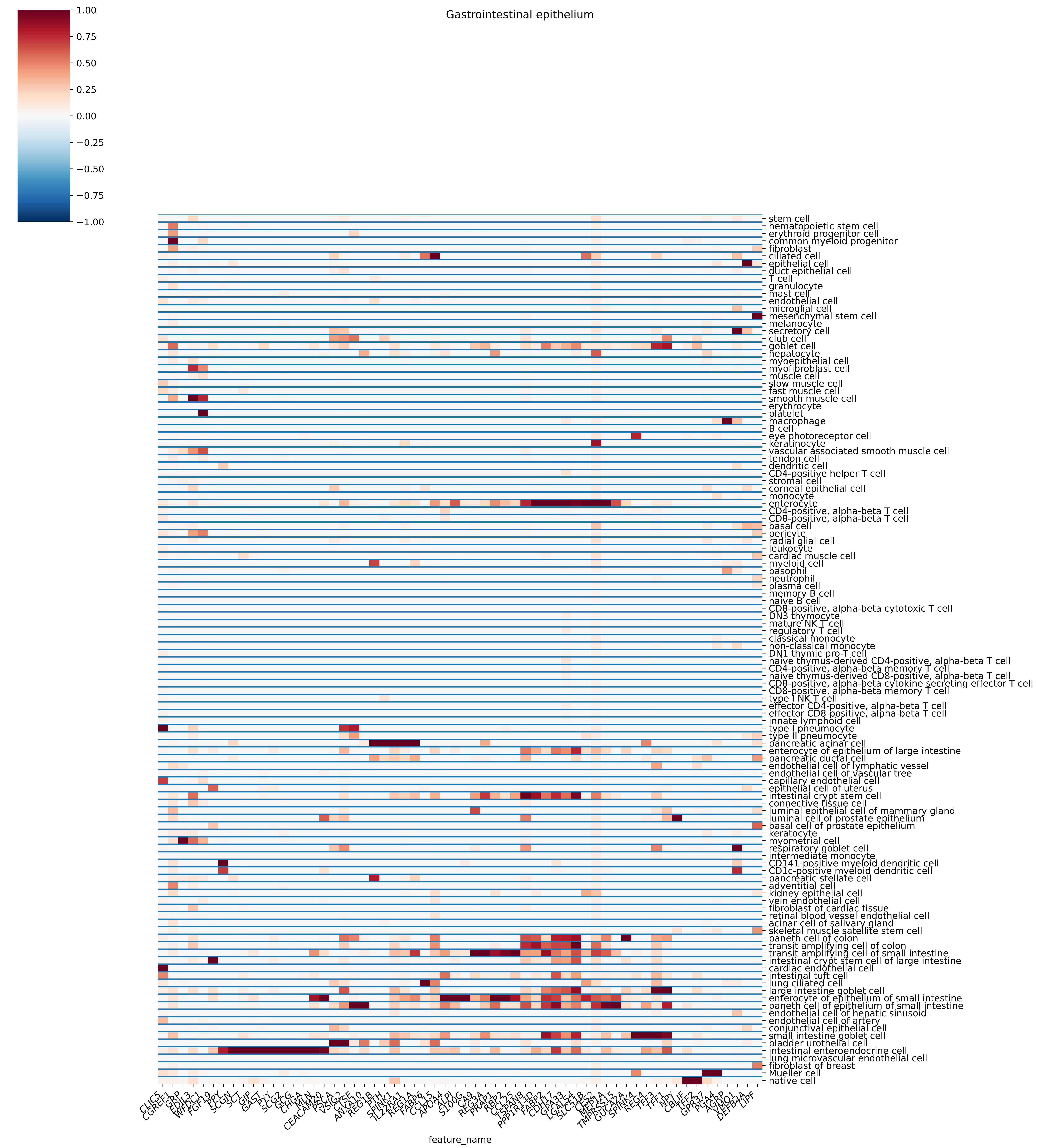
Gastrointestinal epithelium
1.00
0.75
0.50
0.25
0.00
−0.25
−0.50
−0.75
−1.00
stem cell
hematopoietic stem cell
erythroid progenitor cell
common myeloid progenitor
fibroblast
ciliated cell
epithelial cell
duct epithelial cell
T cell
granulocyte
mast cell
endothelial cell
microglial cell
mesenchymal stem cell
melanocyte
secretory cell
club cell
goblet cell
hepatocyte
myoepithelial cell
myofibroblast cell
muscle cell
slow muscle cell
fast muscle cell
smooth muscle cell
erythrocyte
platelet
macrophage
B cell
eye photoreceptor cell
keratinocyte
vascular associated smooth muscle cell
tendon cell
dendritic cell
CD4-positive helper T cell
stromal cell
corneal epithelial cell
monocyte
enterocyte
CD4-positive, alpha-beta T cell
CD8-positive, alpha-beta T cell
basal cell
pericyte
radial glial cell
leukocyte
cardiac muscle cell
myeloid cell
basophil
neutrophil
plasma cell
memory B cell
naive B cell
CD8-positive, alpha-beta cytotoxic T cell
DN3 thymocyte
mature NK T cell
regulatory T cell
classical monocyte
non-classical monocyte
DN1 thymic pro-T cell
naive thymus-derived CD4-positive, alpha-beta T cell
CD4-positive, alpha-beta memory T cell
naive thymus-derived CD8-positive, alpha-beta T cell
CD8-positive, alpha-beta cytokine secreting effector T cell
CD8-positive, alpha-beta memory T cell
type I NK T cell
effector CD4-positive, alpha-beta T cell
effector CD8-positive, alpha-beta T cell
innate lymphoid cell
type I pneumocyte
type II pneumocyte
pancreatic acinar cell
enterocyte of epithelium of large intestine
pancreatic ductal cell
endothelial cell of lymphatic vessel
endothelial cell of vascular tree
capillary endothelial cell
epithelial cell of uterus
intestinal crypt stem cell
connective tissue cell
luminal epithelial cell of mammary gland
luminal cell of prostate epithelium
basal cell of prostate epithelium
keratocyte
myometrial cell
respiratory goblet cell
intermediate monocyte
CD141-positive myeloid dendritic cell
CD1c-positive myeloid dendritic cell
pancreatic stellate cell
adventitial cell
kidney epithelial cell
vein endothelial cell
fibroblast of cardiac tissue
retinal blood vessel endothelial cell
acinar cell of salivary gland
skeletal muscle satellite stem cell
paneth cell of colon
transit amplifying cell of colon
transit amplifying cell of small intestine
intestinal crypt stem cell of large intestine
cardiac endothelial cell
intestinal tuft cell
lung ciliated cell
large intestine goblet cell
enterocyte of epithelium of small intestine
paneth cell of epithelium of small intestine
endothelial cell of hepatic sinusoid
endothelial cell of artery
conjunctival epithelial cell
small intestine goblet cell
bladder urothelial cell
intestinal enteroendocrine cell
lung microvascular endothelial cell
fibroblast of breast
Mueller cell
native cell
CLIC5
CGREF1
GRP
EDIL3
WFDC1
FGF19
PPY
SCGN
SCT
GIP
GAST
PYY
SCG2
GCG
CHGA
MLN
CEACAM20
PSCA
VSIG2
CTSE
ANXA10
REG1B
PTH
SPINK1
IL22RA1
REG1A
FABP6
CCL15
APOA4
ALPI
S100G
CA9
REG3A
PRAP1
RBP2
CCL25
TSPAN8
PPP1R14D
FABP2
CDH17
GPA33
LGALS4
SLC51B
CES2
MEP1A
TMPRSS15
GUCA2A
SPINK4
REG4
TFF3
TFF1
NPY
CBLIF
TFF2
GPR37
PGA4
AGRP
VMO1
DEFB4A
LIPF
feature_name

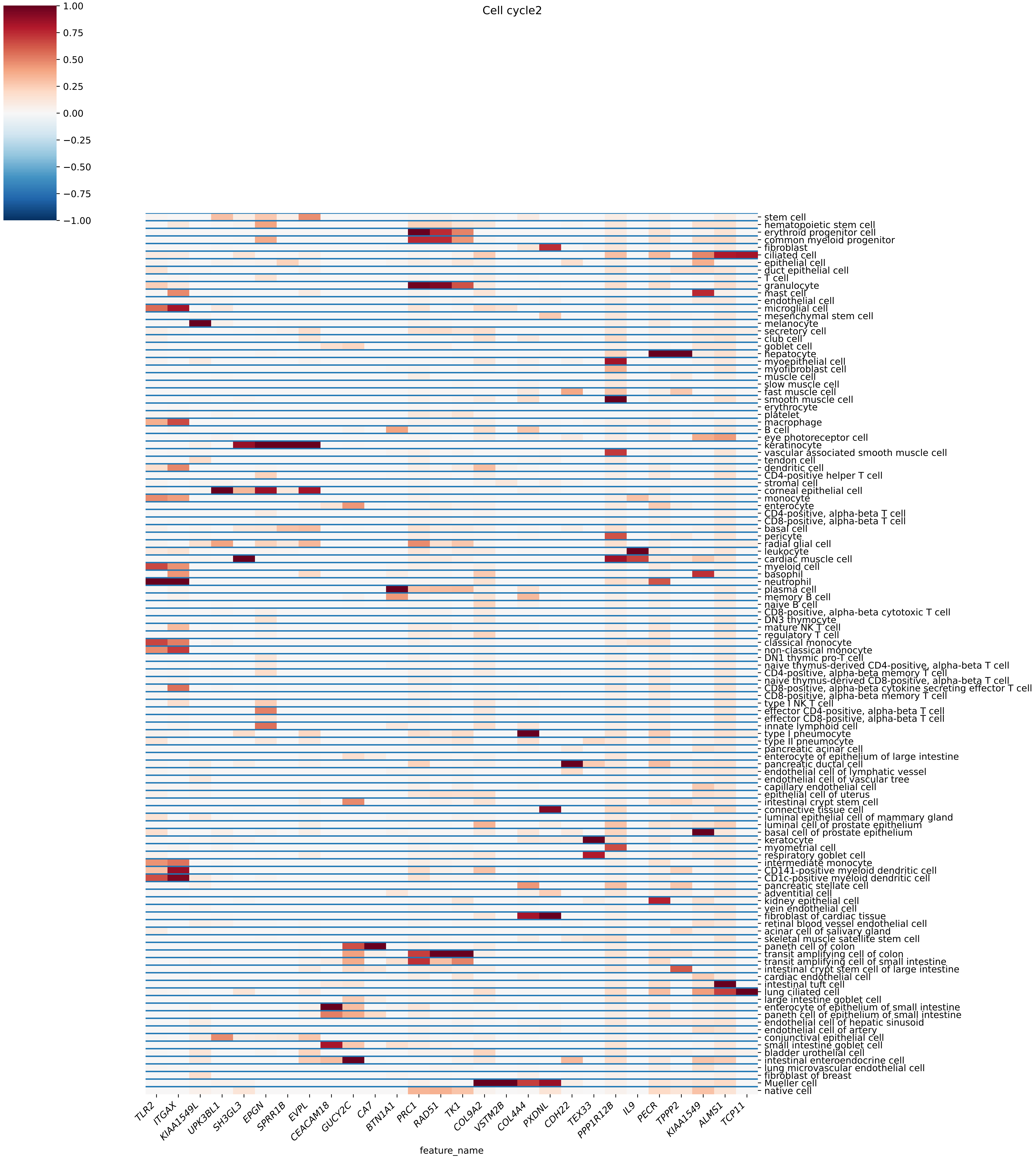
Cell cycle2
1.00
0.75
0.50
0.25
0.00
−0.25
−0.50
−0.75
−1.00
stem cell
hematopoietic stem cell
erythroid progenitor cell
common myeloid progenitor
fibroblast
ciliated cell
epithelial cell
duct epithelial cell
T cell
granulocyte
mast cell
endothelial cell
microglial cell
mesenchymal stem cell
melanocyte
secretory cell
club cell
goblet cell
hepatocyte
myoepithelial cell
myofibroblast cell
muscle cell
slow muscle cell
fast muscle cell
smooth muscle cell
erythrocyte
platelet
macrophage
B cell
eye photoreceptor cell
keratinocyte
vascular associated smooth muscle cell
tendon cell
dendritic cell
CD4-positive helper T cell
stromal cell
corneal epithelial cell
monocyte
enterocyte
CD4-positive, alpha-beta T cell
CD8-positive, alpha-beta T cell
basal cell
pericyte
radial glial cell
leukocyte
cardiac muscle cell
myeloid cell
basophil
neutrophil
plasma cell
memory B cell
naive B cell
CD8-positive, alpha-beta cytotoxic T cell
DN3 thymocyte
mature NK T cell
regulatory T cell
classical monocyte
non-classical monocyte
DN1 thymic pro-T cell
naive thymus-derived CD4-positive, alpha-beta T cell
CD4-positive, alpha-beta memory T cell
naive thymus-derived CD8-positive, alpha-beta T cell
CD8-positive, alpha-beta cytokine secreting effector T cell
CD8-positive, alpha-beta memory T cell
type I NK T cell
effector CD4-positive, alpha-beta T cell
effector CD8-positive, alpha-beta T cell
innate lymphoid cell
type I pneumocyte
type II pneumocyte
pancreatic acinar cell
enterocyte of epithelium of large intestine
pancreatic ductal cell
endothelial cell of lymphatic vessel
endothelial cell of vascular tree
capillary endothelial cell
epithelial cell of uterus
intestinal crypt stem cell
connective tissue cell
luminal epithelial cell of mammary gland
luminal cell of prostate epithelium
basal cell of prostate epithelium
keratocyte
myometrial cell
respiratory goblet cell
intermediate monocyte
CD141-positive myeloid dendritic cell
CD1c-positive myeloid dendritic cell
pancreatic stellate cell
adventitial cell
kidney epithelial cell
vein endothelial cell
fibroblast of cardiac tissue
retinal blood vessel endothelial cell
acinar cell of salivary gland
skeletal muscle satellite stem cell
paneth cell of colon
transit amplifying cell of colon
transit amplifying cell of small intestine
intestinal crypt stem cell of large intestine
cardiac endothelial cell
intestinal tuft cell
lung ciliated cell
large intestine goblet cell
enterocyte of epithelium of small intestine
paneth cell of epithelium of small intestine
endothelial cell of hepatic sinusoid
endothelial cell of artery
conjunctival epithelial cell
small intestine goblet cell
bladder urothelial cell
intestinal enteroendocrine cell
lung microvascular endothelial cell
fibroblast of breast
Mueller cell
native cell
TLR2
ITGAX
KIAA1549L
UPK3BL1
SH3GL3
EPGN
SPRR1B
EVPL
CEACAM18
GUCY2C
CA7
BTN1A1
PRC1
RAD51
TK1
COL9A2
VSTM2B
COL4A4
PXDNL
CDH22
TEX33
PPP1R12B
IL9
PECR
TPPP2
KIAA1549
ALMS1
TCP11
feature_name

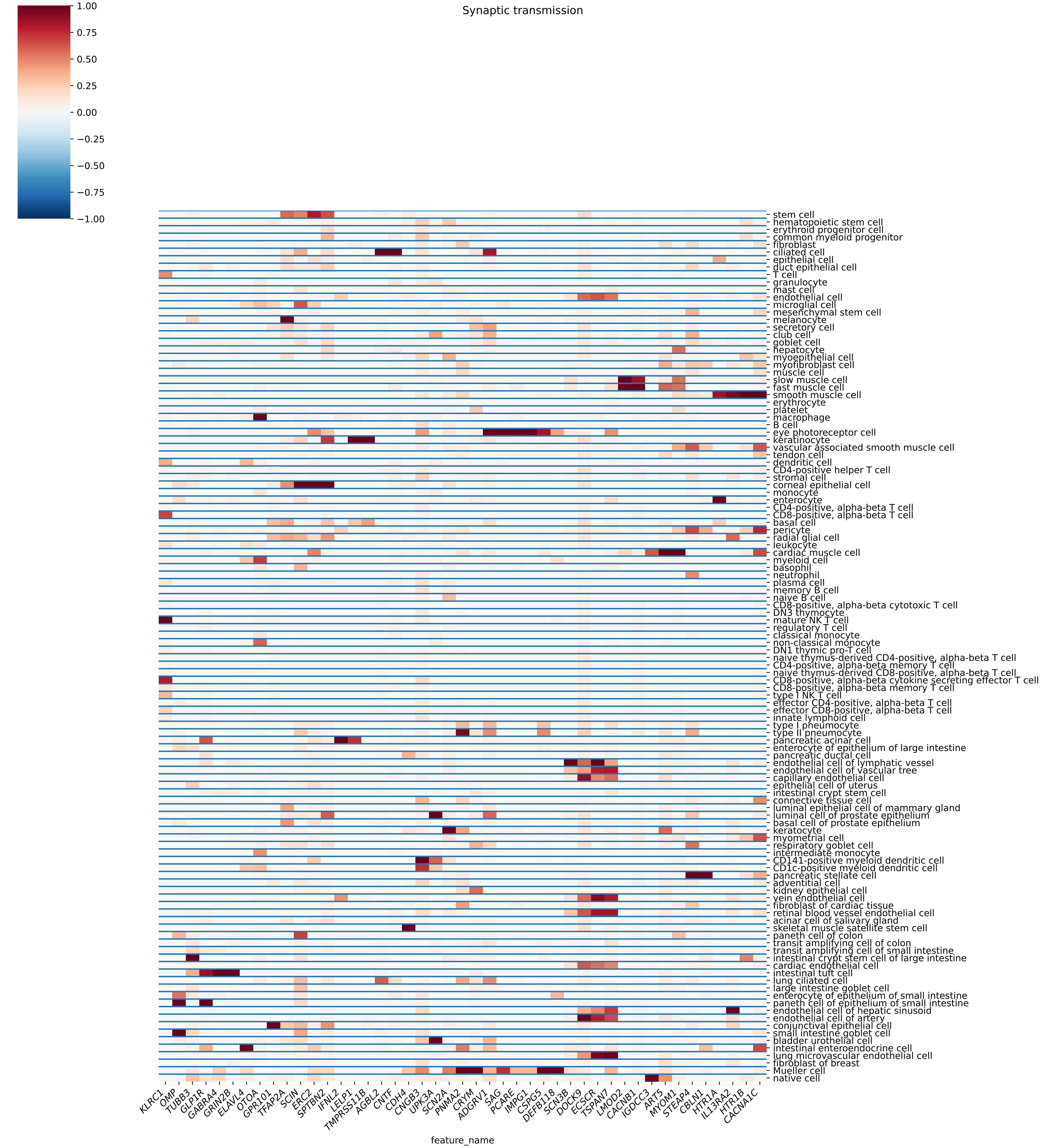
Synaptic transmission
1.00
0.75
0.50
0.25
0.00
−0.25
−0.50
−0.75
−1.00
stem cell
hematopoietic stem cell
erythroid progenitor cell
common myeloid progenitor
fibroblast
ciliated cell
epithelial cell
duct epithelial cell
T cell
granulocyte
mast cell
endothelial cell
microglial cell
mesenchymal stem cell
melanocyte
secretory cell
club cell
goblet cell
hepatocyte
myoepithelial cell
myofibroblast cell
muscle cell
slow muscle cell
fast muscle cell
smooth muscle cell
erythrocyte
platelet
macrophage
B cell
eye photoreceptor cell
keratinocyte
vascular associated smooth muscle cell
tendon cell
dendritic cell
CD4-positive helper T cell
stromal cell
corneal epithelial cell
monocyte
enterocyte
CD4-positive, alpha-beta T cell
CD8-positive, alpha-beta T cell
basal cell
pericyte
radial glial cell
leukocyte
cardiac muscle cell
myeloid cell
basophil
neutrophil
plasma cell
memory B cell
naive B cell
CD8-positive, alpha-beta cytotoxic T cell
DN3 thymocyte
mature NK T cell
regulatory T cell
classical monocyte
non-classical monocyte
DN1 thymic pro-T cell
naive thymus-derived CD4-positive, alpha-beta T cell
CD4-positive, alpha-beta memory T cell
naive thymus-derived CD8-positive, alpha-beta T cell
CD8-positive, alpha-beta cytokine secreting effector T cell
CD8-positive, alpha-beta memory T cell
type I NK T cell
effector CD4-positive, alpha-beta T cell
effector CD8-positive, alpha-beta T cell
innate lymphoid cell
type I pneumocyte
type II pneumocyte
pancreatic acinar cell
enterocyte of epithelium of large intestine
pancreatic ductal cell
endothelial cell of lymphatic vessel
endothelial cell of vascular tree
capillary endothelial cell
epithelial cell of uterus
intestinal crypt stem cell
connective tissue cell
luminal epithelial cell of mammary gland
luminal cell of prostate epithelium
basal cell of prostate epithelium
keratocyte
myometrial cell
respiratory goblet cell
intermediate monocyte
CD141-positive myeloid dendritic cell
CD1c-positive myeloid dendritic cell
pancreatic stellate cell
adventitial cell
kidney epithelial cell
vein endothelial cell
fibroblast of cardiac tissue
retinal blood vessel endothelial cell
acinar cell of salivary gland
skeletal muscle satellite stem cell
paneth cell of colon
transit amplifying cell of colon
transit amplifying cell of small intestine
intestinal crypt stem cell of large intestine
cardiac endothelial cell
intestinal tuft cell
lung ciliated cell
large intestine goblet cell
enterocyte of epithelium of small intestine
paneth cell of epithelium of small intestine
endothelial cell of hepatic sinusoid
endothelial cell of artery
conjunctival epithelial cell
small intestine goblet cell
bladder urothelial cell
intestinal enteroendocrine cell
lung microvascular endothelial cell
fibroblast of breast
Mueller cell
native cell
KLRC1
OMP
TUBB3
GLP1R
GABRA4
GRIN2B
ELAVL4
OTOA
GPR101
TFAP2A
SCIN
ERC2
SPTBN2
IFNL2
LELP1
TMPRSS11B
AGBL2
CNTF
CDH4
CNGB3
UPK3A
SCN2A
PNMA2
CRYM
ADGRV1
SAG
PCARE
IMPG1
CSPG5
DEFB118
SCN3B
DOCK9
ECSCR
TSPAN7
LMOD2
CACNB1
IGDCC3
ART5
MYOM1
STEAP4
CBLN1
HTR1A
IL13RA2
HTR1B
CACNA1C
feature_name

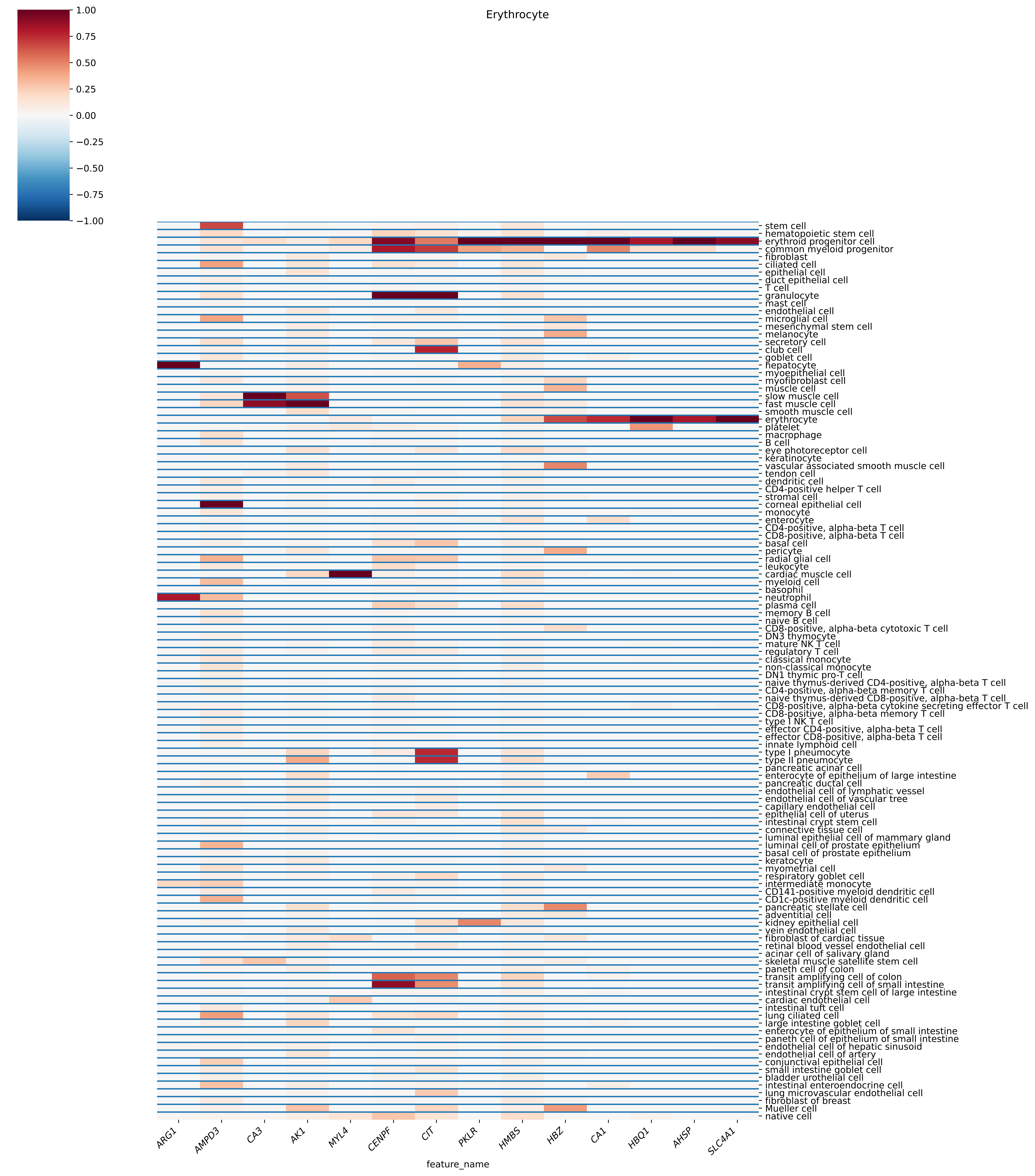
Erythrocyte
1.00
0.75
0.50
0.25
0.00
−0.25
−0.50
−0.75
−1.00
stem cell
hematopoietic stem cell
erythroid progenitor cell
common myeloid progenitor
fibroblast
ciliated cell
epithelial cell
duct epithelial cell
T cell
granulocyte
mast cell
endothelial cell
microglial cell
mesenchymal stem cell
melanocyte
secretory cell
club cell
goblet cell
hepatocyte
myoepithelial cell
myofibroblast cell
muscle cell
slow muscle cell
fast muscle cell
smooth muscle cell
erythrocyte
platelet
macrophage
B cell
eye photoreceptor cell
keratinocyte
vascular associated smooth muscle cell
tendon cell
dendritic cell
CD4-positive helper T cell
stromal cell
corneal epithelial cell
monocyte
enterocyte
CD4-positive, alpha-beta T cell
CD8-positive, alpha-beta T cell
basal cell
pericyte
radial glial cell
leukocyte
cardiac muscle cell
myeloid cell
basophil
neutrophil
plasma cell
memory B cell
naive B cell
CD8-positive, alpha-beta cytotoxic T cell
DN3 thymocyte
mature NK T cell
regulatory T cell
classical monocyte
non-classical monocyte
DN1 thymic pro-T cell
naive thymus-derived CD4-positive, alpha-beta T cell
CD4-positive, alpha-beta memory T cell
naive thymus-derived CD8-positive, alpha-beta T cell
CD8-positive, alpha-beta cytokine secreting effector T cell
CD8-positive, alpha-beta memory T cell
type I NK T cell
effector CD4-positive, alpha-beta T cell
effector CD8-positive, alpha-beta T cell
innate lymphoid cell
type I pneumocyte
type II pneumocyte
pancreatic acinar cell
enterocyte of epithelium of large intestine
pancreatic ductal cell
endothelial cell of lymphatic vessel
endothelial cell of vascular tree
capillary endothelial cell
epithelial cell of uterus
intestinal crypt stem cell
connective tissue cell
luminal epithelial cell of mammary gland
luminal cell of prostate epithelium
basal cell of prostate epithelium
keratocyte
myometrial cell
respiratory goblet cell
intermediate monocyte
CD141-positive myeloid dendritic cell
CD1c-positive myeloid dendritic cell
pancreatic stellate cell
adventitial cell
kidney epithelial cell
vein endothelial cell
fibroblast of cardiac tissue
retinal blood vessel endothelial cell
acinar cell of salivary gland
skeletal muscle satellite stem cell
paneth cell of colon
transit amplifying cell of colon
transit amplifying cell of small intestine
intestinal crypt stem cell of large intestine
cardiac endothelial cell
intestinal tuft cell
lung ciliated cell
large intestine goblet cell
enterocyte of epithelium of small intestine
paneth cell of epithelium of small intestine
endothelial cell of hepatic sinusoid
endothelial cell of artery
conjunctival epithelial cell
small intestine goblet cell
bladder urothelial cell
intestinal enteroendocrine cell
lung microvascular endothelial cell
fibroblast of breast
Mueller cell
native cell
ARG1
AMPD3
CA3
AK1
MYL4
CENPF
CIT
PKLR
HMBS
HBZ
CA1
HBQ1
AHSP
SLC4A1
feature_name

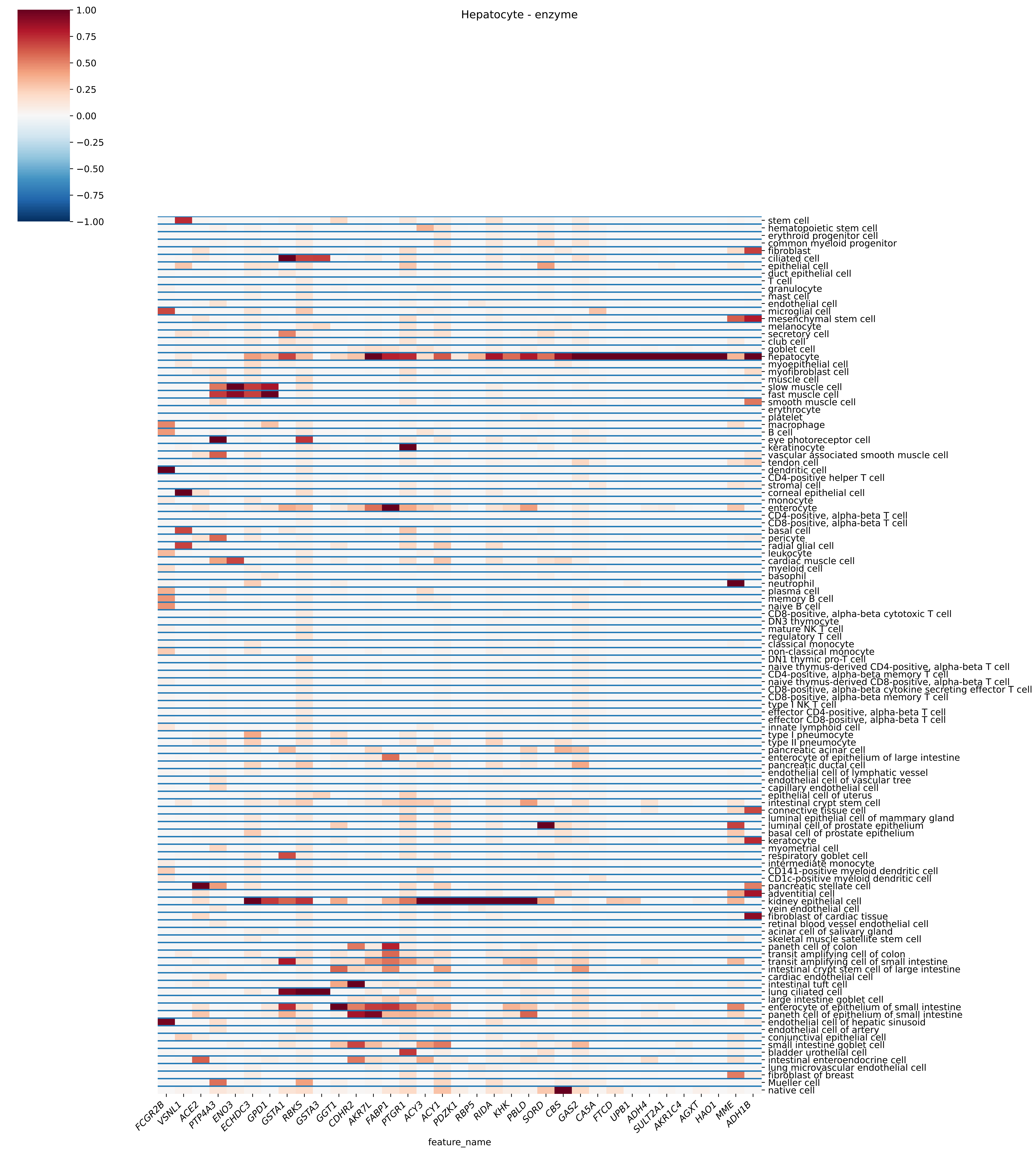
Hepatocyte - enzyme
1.00
0.75
0.50
0.25
0.00
−0.25
−0.50
−0.75
−1.00
stem cell
hematopoietic stem cell
erythroid progenitor cell
common myeloid progenitor
fibroblast
ciliated cell
epithelial cell
duct epithelial cell
T cell
granulocyte
mast cell
endothelial cell
microglial cell
mesenchymal stem cell
melanocyte
secretory cell
club cell
goblet cell
hepatocyte
myoepithelial cell
myofibroblast cell
muscle cell
slow muscle cell
fast muscle cell
smooth muscle cell
erythrocyte
platelet
macrophage
B cell
eye photoreceptor cell
keratinocyte
vascular associated smooth muscle cell
tendon cell
dendritic cell
CD4-positive helper T cell
stromal cell
corneal epithelial cell
monocyte
enterocyte
CD4-positive, alpha-beta T cell
CD8-positive, alpha-beta T cell
basal cell
pericyte
radial glial cell
leukocyte
cardiac muscle cell
myeloid cell
basophil
neutrophil
plasma cell
memory B cell
naive B cell
CD8-positive, alpha-beta cytotoxic T cell
DN3 thymocyte
mature NK T cell
regulatory T cell
classical monocyte
non-classical monocyte
DN1 thymic pro-T cell
naive thymus-derived CD4-positive, alpha-beta T cell
CD4-positive, alpha-beta memory T cell
naive thymus-derived CD8-positive, alpha-beta T cell
CD8-positive, alpha-beta cytokine secreting effector T cell
CD8-positive, alpha-beta memory T cell
type I NK T cell
effector CD4-positive, alpha-beta T cell
effector CD8-positive, alpha-beta T cell
innate lymphoid cell
type I pneumocyte
type II pneumocyte
pancreatic acinar cell
enterocyte of epithelium of large intestine
pancreatic ductal cell
endothelial cell of lymphatic vessel
endothelial cell of vascular tree
capillary endothelial cell
epithelial cell of uterus
intestinal crypt stem cell
connective tissue cell
luminal epithelial cell of mammary gland
luminal cell of prostate epithelium
basal cell of prostate epithelium
keratocyte
myometrial cell
respiratory goblet cell
intermediate monocyte
CD141-positive myeloid dendritic cell
CD1c-positive myeloid dendritic cell
pancreatic stellate cell
adventitial cell
kidney epithelial cell
vein endothelial cell
fibroblast of cardiac tissue
retinal blood vessel endothelial cell
acinar cell of salivary gland
skeletal muscle satellite stem cell
paneth cell of colon
transit amplifying cell of colon
transit amplifying cell of small intestine
intestinal crypt stem cell of large intestine
cardiac endothelial cell
intestinal tuft cell
lung ciliated cell
large intestine goblet cell
enterocyte of epithelium of small intestine
paneth cell of epithelium of small intestine
endothelial cell of hepatic sinusoid
endothelial cell of artery
conjunctival epithelial cell
small intestine goblet cell
bladder urothelial cell
intestinal enteroendocrine cell
lung microvascular endothelial cell
fibroblast of breast
Mueller cell
native cell
FCGR2B
VSNL1
ACE2
PTP4A3
ENO3
ECHDC3
GPD1
GSTA1
RBKS
GSTA3
GGT1
CDHR2
AKR7L
FABP1
PTGR1
ACY3
ACY1
PDZK1
RBP5
RIDA
KHK
PBLD
SORD
CBS
GAS2
CA5A
FTCD
UPB1
ADH4
SULT2A1
AKR1C4
AGXT
HAO1
MME
ADH1B
feature_name

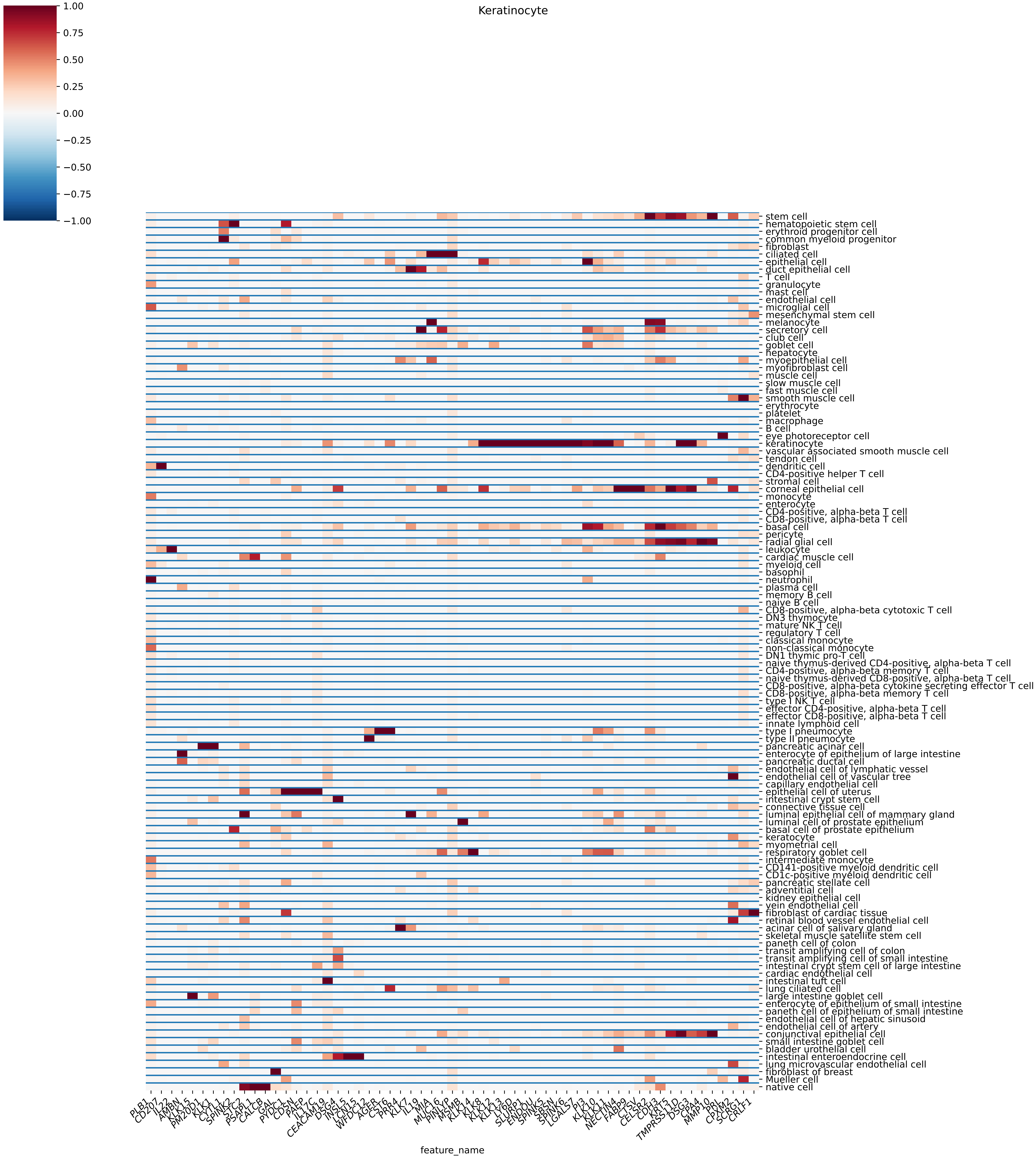
Keratinocyte
1.00
0.75
0.50
0.25
0.00
−0.25
−0.50
−0.75
−1.00
stem cell
hematopoietic stem cell
erythroid progenitor cell
common myeloid progenitor
fibroblast
ciliated cell
epithelial cell
duct epithelial cell
T cell
granulocyte
mast cell
endothelial cell
microglial cell
mesenchymal stem cell
melanocyte
secretory cell
club cell
goblet cell
hepatocyte
myoepithelial cell
myofibroblast cell
muscle cell
slow muscle cell
fast muscle cell
smooth muscle cell
erythrocyte
platelet
macrophage
B cell
eye photoreceptor cell
keratinocyte
vascular associated smooth muscle cell
tendon cell
dendritic cell
CD4-positive helper T cell
stromal cell
corneal epithelial cell
monocyte
enterocyte
CD4-positive, alpha-beta T cell
CD8-positive, alpha-beta T cell
basal cell
pericyte
radial glial cell
leukocyte
cardiac muscle cell
myeloid cell
basophil
neutrophil
plasma cell
memory B cell
naive B cell
CD8-positive, alpha-beta cytotoxic T cell
DN3 thymocyte
mature NK T cell
regulatory T cell
classical monocyte
non-classical monocyte
DN1 thymic pro-T cell
naive thymus-derived CD4-positive, alpha-beta T cell
CD4-positive, alpha-beta memory T cell
naive thymus-derived CD8-positive, alpha-beta T cell
CD8-positive, alpha-beta cytokine secreting effector T cell
CD8-positive, alpha-beta memory T cell
type I NK T cell
effector CD4-positive, alpha-beta T cell
effector CD8-positive, alpha-beta T cell
innate lymphoid cell
type I pneumocyte
type II pneumocyte
pancreatic acinar cell
enterocyte of epithelium of large intestine
pancreatic ductal cell
endothelial cell of lymphatic vessel
endothelial cell of vascular tree
capillary endothelial cell
epithelial cell of uterus
intestinal crypt stem cell
connective tissue cell
luminal epithelial cell of mammary gland
luminal cell of prostate epithelium
basal cell of prostate epithelium
keratocyte
myometrial cell
respiratory goblet cell
intermediate monocyte
CD141-positive myeloid dendritic cell
CD1c-positive myeloid dendritic cell
pancreatic stellate cell
adventitial cell
kidney epithelial cell
vein endothelial cell
fibroblast of cardiac tissue
retinal blood vessel endothelial cell
acinar cell of salivary gland
skeletal muscle satellite stem cell
paneth cell of colon
transit amplifying cell of colon
transit amplifying cell of small intestine
intestinal crypt stem cell of large intestine
cardiac endothelial cell
intestinal tuft cell
lung ciliated cell
large intestine goblet cell
enterocyte of epithelium of small intestine
paneth cell of epithelium of small intestine
endothelial cell of hepatic sinusoid
endothelial cell of artery
conjunctival epithelial cell
small intestine goblet cell
bladder urothelial cell
intestinal enteroendocrine cell
lung microvascular endothelial cell
fibroblast of breast
Mueller cell
native cell
PLB1
CD207
IL22
AMBN
KLK15
PM20D1
KLK1
CYTL1
SPINK2
STC2
PSAPL1
CALCB
GAL
PYDC1
CDSN
PAEP
IL17C
CEACAM19
DSG4
INSL5
LCN15
WFDC12
AGER
CST6
PRR4
KLK7
IL19
MIA
MUC16
PINLYP
MSMB
KLK14
KLK8
KLK12
KLK13
LY6D
SLURP1
ENDOU
SPINK5
SBSN
SPINK6
LGALS7
PI3
KLK10
KLK11
NECTIN4
FABP9
CTSV
CELSR2
CDH3
KRT5
TMPRSS11D
DSG3
CPA4
MMP10
PRL
CPXM2
SCRG1
CRLF1
feature_name

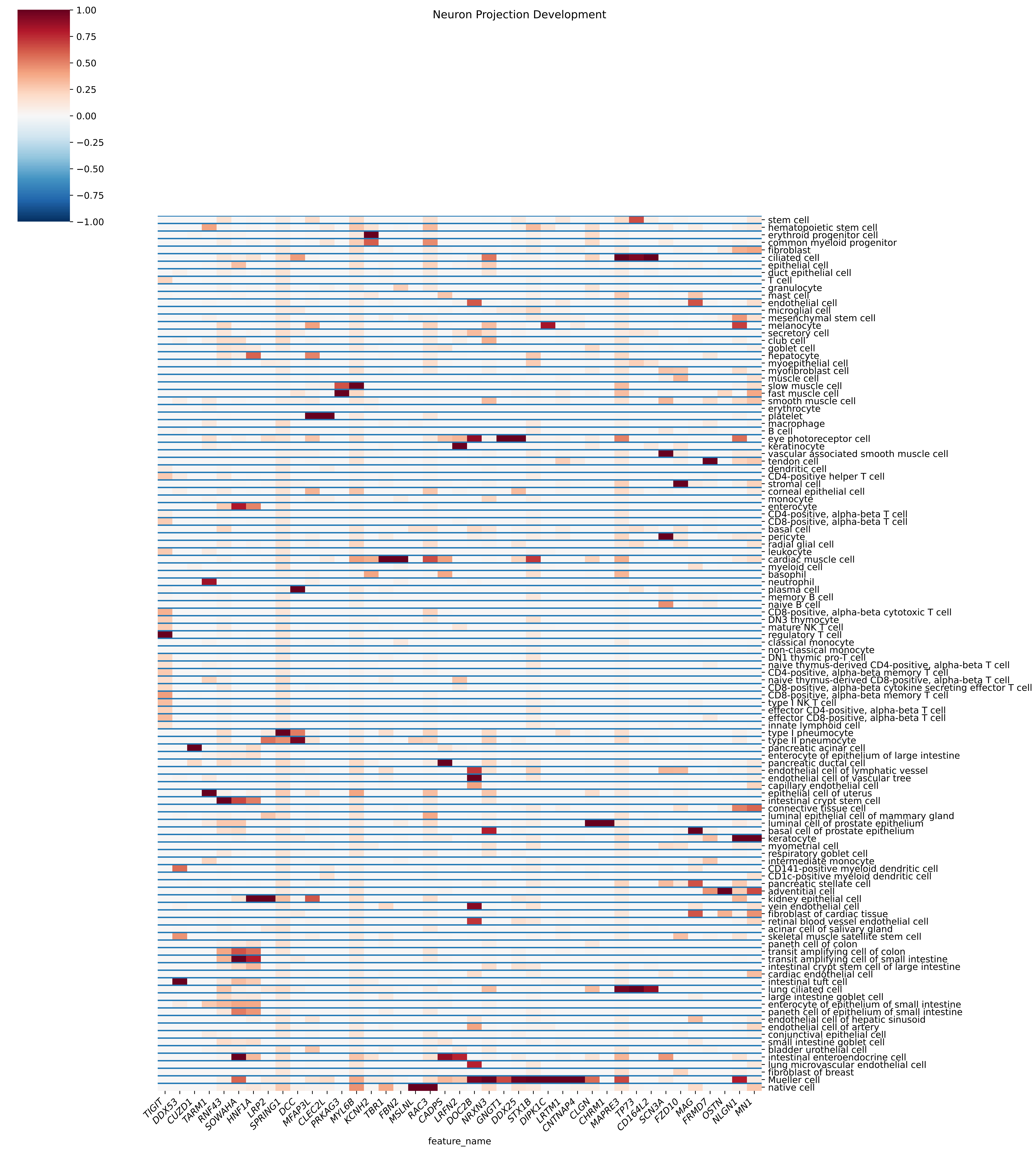
Neuron Projection Development
1.00
0.75
0.50
0.25
0.00
−0.25
−0.50
−0.75
−1.00
stem cell
hematopoietic stem cell
erythroid progenitor cell
common myeloid progenitor
fibroblast
ciliated cell
epithelial cell
duct epithelial cell
T cell
granulocyte
mast cell
endothelial cell
microglial cell
mesenchymal stem cell
melanocyte
secretory cell
club cell
goblet cell
hepatocyte
myoepithelial cell
myofibroblast cell
muscle cell
slow muscle cell
fast muscle cell
smooth muscle cell
erythrocyte
platelet
macrophage
B cell
eye photoreceptor cell
keratinocyte
vascular associated smooth muscle cell
tendon cell
dendritic cell
CD4-positive helper T cell
stromal cell
corneal epithelial cell
monocyte
enterocyte
CD4-positive, alpha-beta T cell
CD8-positive, alpha-beta T cell
basal cell
pericyte
radial glial cell
leukocyte
cardiac muscle cell
myeloid cell
basophil
neutrophil
plasma cell
memory B cell
naive B cell
CD8-positive, alpha-beta cytotoxic T cell
DN3 thymocyte
mature NK T cell
regulatory T cell
classical monocyte
non-classical monocyte
DN1 thymic pro-T cell
naive thymus-derived CD4-positive, alpha-beta T cell
CD4-positive, alpha-beta memory T cell
naive thymus-derived CD8-positive, alpha-beta T cell
CD8-positive, alpha-beta cytokine secreting effector T cell
CD8-positive, alpha-beta memory T cell
type I NK T cell
effector CD4-positive, alpha-beta T cell
effector CD8-positive, alpha-beta T cell
innate lymphoid cell
type I pneumocyte
type II pneumocyte
pancreatic acinar cell
enterocyte of epithelium of large intestine
pancreatic ductal cell
endothelial cell of lymphatic vessel
endothelial cell of vascular tree
capillary endothelial cell
epithelial cell of uterus
intestinal crypt stem cell
connective tissue cell
luminal epithelial cell of mammary gland
luminal cell of prostate epithelium
basal cell of prostate epithelium
keratocyte
myometrial cell
respiratory goblet cell
intermediate monocyte
CD141-positive myeloid dendritic cell
CD1c-positive myeloid dendritic cell
pancreatic stellate cell
adventitial cell
kidney epithelial cell
vein endothelial cell
fibroblast of cardiac tissue
retinal blood vessel endothelial cell
acinar cell of salivary gland
skeletal muscle satellite stem cell
paneth cell of colon
transit amplifying cell of colon
transit amplifying cell of small intestine
intestinal crypt stem cell of large intestine
cardiac endothelial cell
intestinal tuft cell
lung ciliated cell
large intestine goblet cell
enterocyte of epithelium of small intestine
paneth cell of epithelium of small intestine
endothelial cell of hepatic sinusoid
endothelial cell of artery
conjunctival epithelial cell
small intestine goblet cell
bladder urothelial cell
intestinal enteroendocrine cell
lung microvascular endothelial cell
fibroblast of breast
Mueller cell
native cell
TIGIT
DDX53
CUZD1
TARM1
RNF43
SOWAHA
HNF1A
LRP2
SPRING1
DCC
MFAP3L
CLEC2L
PRKAG3
MYL6B
KCNH2
TBR1
FBN2
MSLNL
RAC3
CADPS
LRFN2
DOC2B
NRXN3
GNGT1
DDX25
STX1B
DIPK1C
LRTM1
CNTNAP4
CLGN
CHRM1
MAPRE3
TP73
CD164L2
SCN3A
FZD10
MAG
FRMD7
OSTN
NLGN1
MN1
feature_name

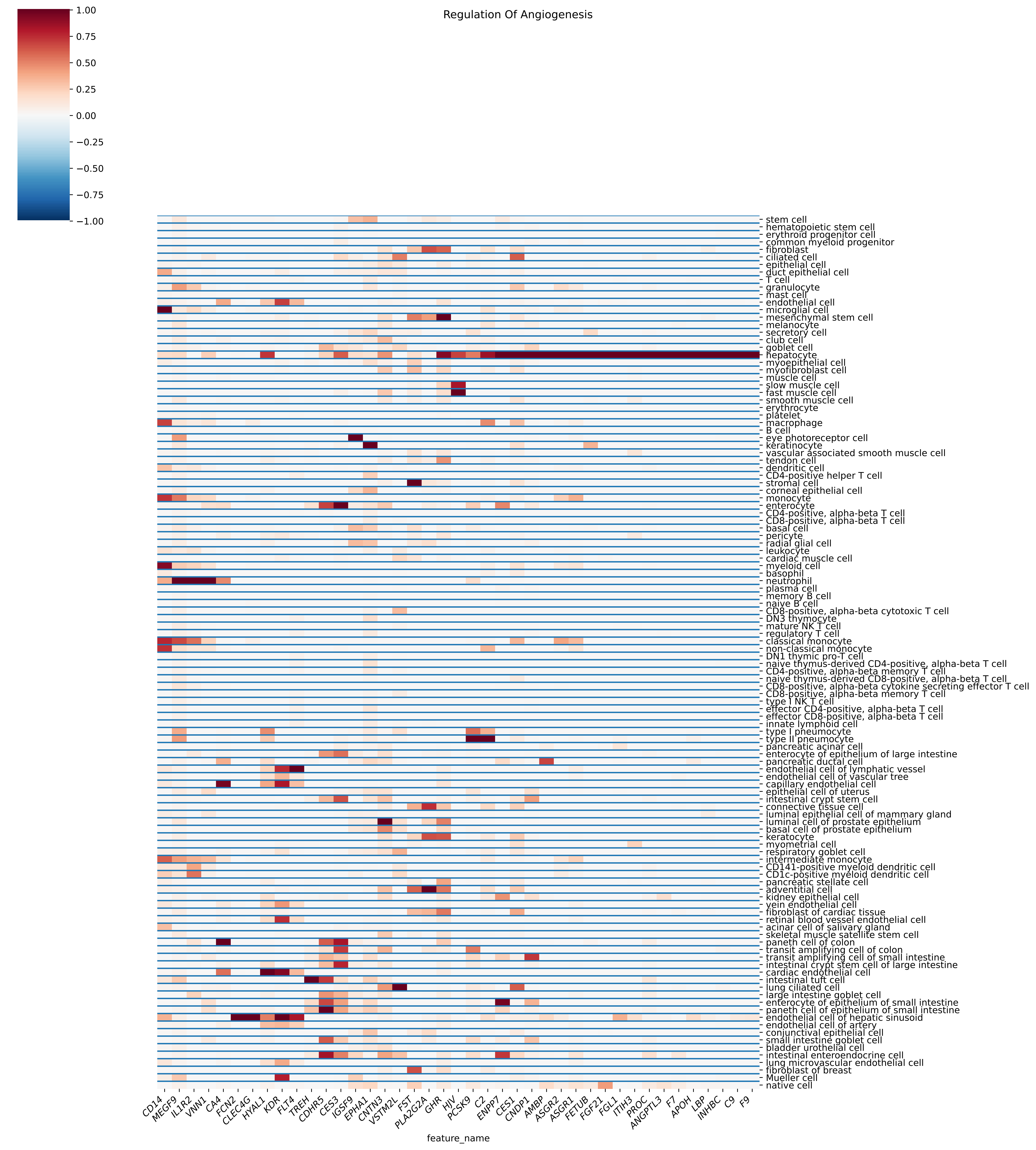

Regulation Of Angiogenesis
1.00
0.75
0.50
0.25
0.00
−0.25
−0.50
−0.75
−1.00
stem cell
hematopoietic stem cell
erythroid progenitor cell
common myeloid progenitor
fibroblast
ciliated cell
epithelial cell
duct epithelial cell
T cell
granulocyte
mast cell
endothelial cell
microglial cell
mesenchymal stem cell
melanocyte
secretory cell
club cell
goblet cell
hepatocyte
myoepithelial cell
myofibroblast cell
muscle cell
slow muscle cell
fast muscle cell
smooth muscle cell
erythrocyte
platelet
macrophage
B cell
eye photoreceptor cell
keratinocyte
vascular associated smooth muscle cell
tendon cell
dendritic cell
CD4-positive helper T cell
stromal cell
corneal epithelial cell
monocyte
enterocyte
CD4-positive, alpha-beta T cell
CD8-positive, alpha-beta T cell
basal cell
pericyte
radial glial cell
leukocyte
cardiac muscle cell
myeloid cell
basophil
neutrophil
plasma cell
memory B cell
naive B cell
CD8-positive, alpha-beta cytotoxic T cell
DN3 thymocyte
mature NK T cell
regulatory T cell
classical monocyte
non-classical monocyte
DN1 thymic pro-T cell
naive thymus-derived CD4-positive, alpha-beta T cell
CD4-positive, alpha-beta memory T cell
naive thymus-derived CD8-positive, alpha-beta T cell
CD8-positive, alpha-beta cytokine secreting effector T cell
CD8-positive, alpha-beta memory T cell
type I NK T cell
effector CD4-positive, alpha-beta T cell
effector CD8-positive, alpha-beta T cell
innate lymphoid cell
type I pneumocyte
type II pneumocyte
pancreatic acinar cell
enterocyte of epithelium of large intestine
pancreatic ductal cell
endothelial cell of lymphatic vessel
endothelial cell of vascular tree
capillary endothelial cell
epithelial cell of uterus
intestinal crypt stem cell
connective tissue cell
luminal epithelial cell of mammary gland
luminal cell of prostate epithelium
basal cell of prostate epithelium
keratocyte
myometrial cell
respiratory goblet cell
intermediate monocyte
CD141-positive myeloid dendritic cell
CD1c-positive myeloid dendritic cell
pancreatic stellate cell
adventitial cell
kidney epithelial cell
vein endothelial cell
fibroblast of cardiac tissue
retinal blood vessel endothelial cell
acinar cell of salivary gland
skeletal muscle satellite stem cell
paneth cell of colon
transit amplifying cell of colon
transit amplifying cell of small intestine
intestinal crypt stem cell of large intestine
cardiac endothelial cell
intestinal tuft cell
lung ciliated cell
large intestine goblet cell
enterocyte of epithelium of small intestine
paneth cell of epithelium of small intestine
endothelial cell of hepatic sinusoid
endothelial cell of artery
conjunctival epithelial cell
small intestine goblet cell
bladder urothelial cell
intestinal enteroendocrine cell
lung microvascular endothelial cell
fibroblast of breast
Mueller cell
native cell
CD14
MEGF9
IL1R2
VNN1
CA4
FCN2
CLEC4G
HYAL1
KDR
FLT4
TREH
CDHR5
CES3
IGSF9
EPHA1
CNTN3
VSTM2L
FST
PLA2G2A
GHR
HJV
PCSK9
C2
ENPP7
CES1
CNDP1
AMBP
ASGR2
ASGR1
FETUB
FGF21
FGL1
ITIH3
PROC
ANGPTL3
F7
APOH
LBP
INHBC
C9
F9
feature_name

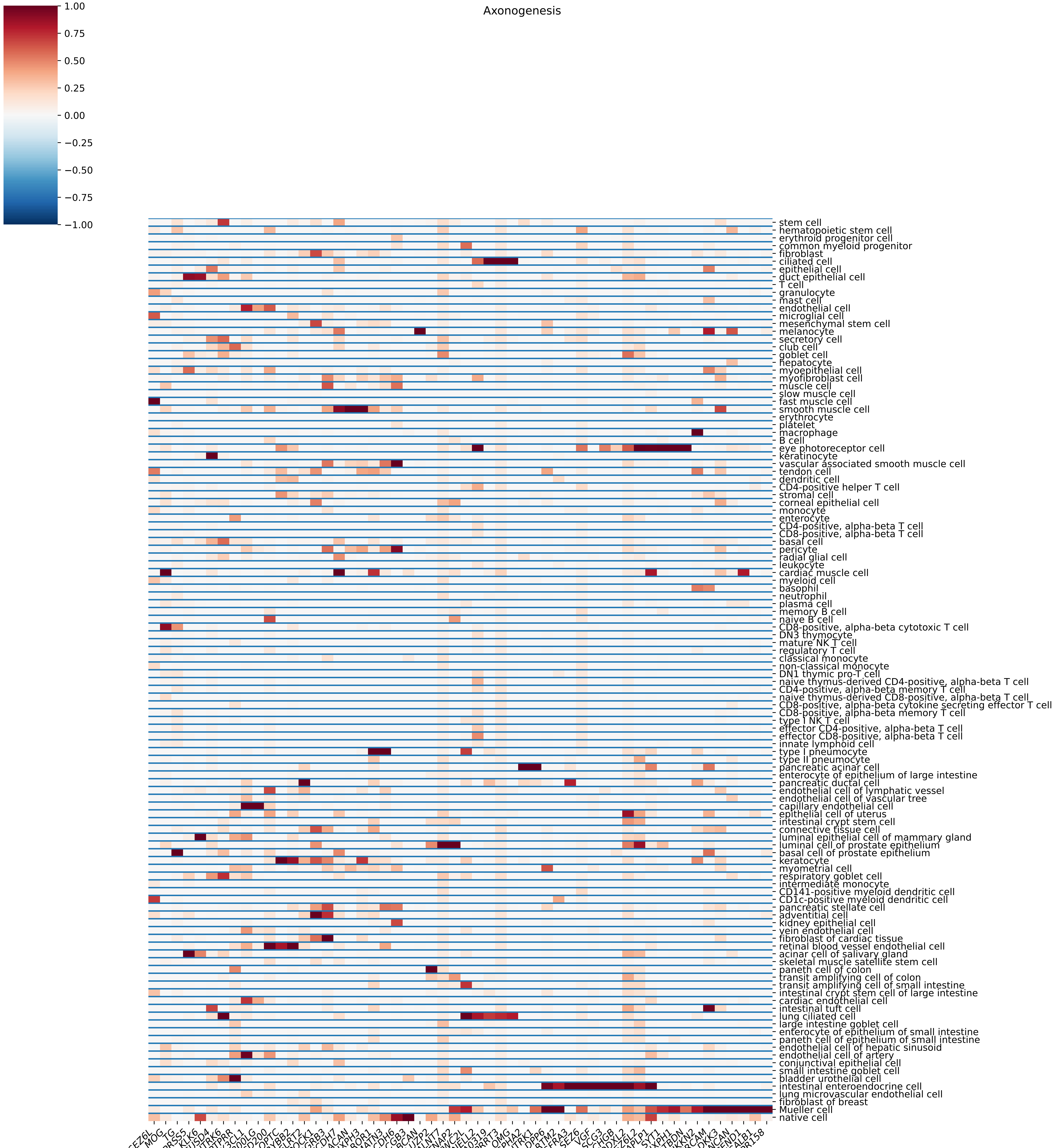

Axonogenesis
1.00
0.75
0.50
0.25
0.00
−0.25
−0.50
−0.75
−1.00
stem cell
hematopoietic stem cell
erythroid progenitor cell
common myeloid progenitor
fibroblast
ciliated cell
epithelial cell
duct epithelial cell
T cell
granulocyte
mast cell
endothelial cell
microglial cell
mesenchymal stem cell
melanocyte
secretory cell
club cell
goblet cell
hepatocyte
myoepithelial cell
myofibroblast cell
muscle cell
slow muscle cell
fast muscle cell
smooth muscle cell
erythrocyte
platelet
macrophage
B cell
eye photoreceptor cell
keratinocyte
vascular associated smooth muscle cell
tendon cell
dendritic cell
CD4-positive helper T cell
stromal cell
corneal epithelial cell
monocyte
enterocyte
CD4-positive, alpha-beta T cell
CD8-positive, alpha-beta T cell
basal cell
pericyte
radial glial cell
leukocyte
cardiac muscle cell
myeloid cell
basophil
neutrophil
plasma cell
memory B cell
naive B cell
CD8-positive, alpha-beta cytotoxic T cell
DN3 thymocyte
mature NK T cell
regulatory T cell
classical monocyte
non-classical monocyte
DN1 thymic pro-T cell
naive thymus-derived CD4-positive, alpha-beta T cell
CD4-positive, alpha-beta memory T cell
naive thymus-derived CD8-positive, alpha-beta T cell
CD8-positive, alpha-beta cytokine secreting effector T cell
CD8-positive, alpha-beta memory T cell
type I NK T cell
effector CD4-positive, alpha-beta T cell
effector CD8-positive, alpha-beta T cell
innate lymphoid cell
type I pneumocyte
type II pneumocyte
pancreatic acinar cell
enterocyte of epithelium of large intestine
pancreatic ductal cell
endothelial cell of lymphatic vessel
endothelial cell of vascular tree
capillary endothelial cell
epithelial cell of uterus
intestinal crypt stem cell
connective tissue cell
luminal epithelial cell of mammary gland
luminal cell of prostate epithelium
basal cell of prostate epithelium
keratocyte
myometrial cell
respiratory goblet cell
intermediate monocyte
CD141-positive myeloid dendritic cell
CD1c-positive myeloid dendritic cell
pancreatic stellate cell
adventitial cell
kidney epithelial cell
vein endothelial cell
fibroblast of cardiac tissue
retinal blood vessel endothelial cell
acinar cell of salivary gland
skeletal muscle satellite stem cell
paneth cell of colon
transit amplifying cell of colon
transit amplifying cell of small intestine
intestinal crypt stem cell of large intestine
cardiac endothelial cell
intestinal tuft cell
lung ciliated cell
large intestine goblet cell
enterocyte of epithelium of small intestine
paneth cell of epithelium of small intestine
endothelial cell of hepatic sinusoid
endothelial cell of artery
conjunctival epithelial cell
small intestine goblet cell
bladder urothelial cell
intestinal enteroendocrine cell
lung microvascular endothelial cell
fibroblast of breast
Mueller cell
native cell
SEZ6L
MOG
TG
TMPRSS5
KLK6
SUSD4
SLITRK6
PTPRR
CX3CL1
CD300LG
CD200
OPTC
CRYBB2
FLRT2
SPOCK1
ADGRB3
PCDH7
ACAN
NXPH3
ROR1
MATN3
CDH6
CGB3
BCAN
LUZP2
GALNT7
CNTNAP2
VWC2L
NELL2
KIAA0319
PRRT3
OMG
GPHA2
SLITRK1
DPP6
LRTM2
GFRA3
SEZ6
VGF
SCG3
CHGB
PODXL2
SEZ6L2
APLP1
SYT1
NXPH1
RTBDN
WFIKKN2
NRCAM
DKK3
NCAN
CEND1
CALB1
GPR158
feature_name

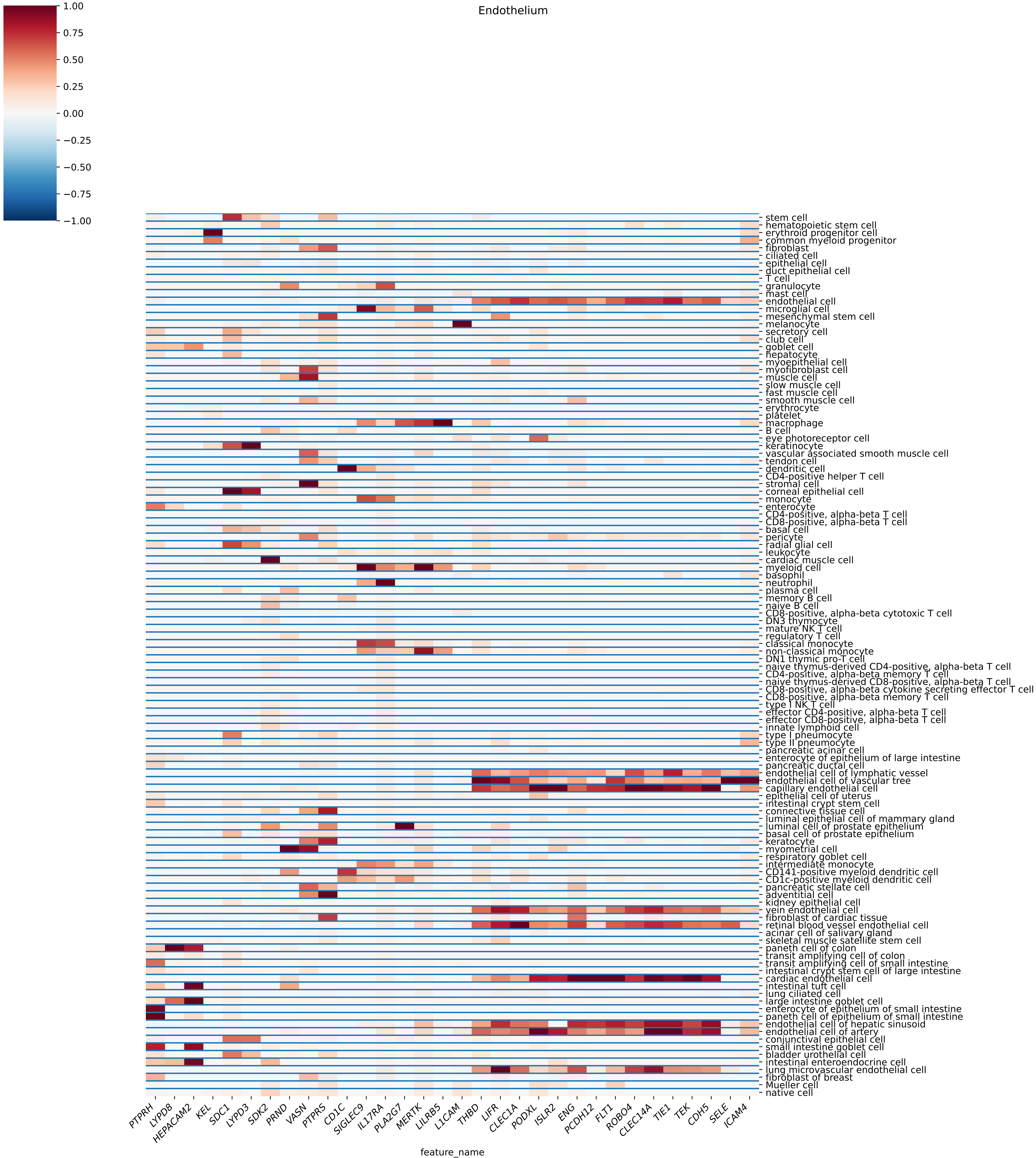
Endothelium
1.00
0.75
0.50
0.25
0.00
−0.25
−0.50
−0.75
−1.00
stem cell
hematopoietic stem cell
erythroid progenitor cell
common myeloid progenitor
fibroblast
ciliated cell
epithelial cell
duct epithelial cell
T cell
granulocyte
mast cell
endothelial cell
microglial cell
mesenchymal stem cell
melanocyte
secretory cell
club cell
goblet cell
hepatocyte
myoepithelial cell
myofibroblast cell
muscle cell
slow muscle cell
fast muscle cell
smooth muscle cell
erythrocyte
platelet
macrophage
B cell
eye photoreceptor cell
keratinocyte
vascular associated smooth muscle cell
tendon cell
dendritic cell
CD4-positive helper T cell
stromal cell
corneal epithelial cell
monocyte
enterocyte
CD4-positive, alpha-beta T cell
CD8-positive, alpha-beta T cell
basal cell
pericyte
radial glial cell
leukocyte
cardiac muscle cell
myeloid cell
basophil
neutrophil
plasma cell
memory B cell
naive B cell
CD8-positive, alpha-beta cytotoxic T cell
DN3 thymocyte
mature NK T cell
regulatory T cell
classical monocyte
non-classical monocyte
DN1 thymic pro-T cell
naive thymus-derived CD4-positive, alpha-beta T cell
CD4-positive, alpha-beta memory T cell
naive thymus-derived CD8-positive, alpha-beta T cell
CD8-positive, alpha-beta cytokine secreting effector T cell
CD8-positive, alpha-beta memory T cell
type I NK T cell
effector CD4-positive, alpha-beta T cell
effector CD8-positive, alpha-beta T cell
innate lymphoid cell
type I pneumocyte
type II pneumocyte
pancreatic acinar cell
enterocyte of epithelium of large intestine
pancreatic ductal cell
endothelial cell of lymphatic vessel
endothelial cell of vascular tree
capillary endothelial cell
epithelial cell of uterus
intestinal crypt stem cell
connective tissue cell
luminal epithelial cell of mammary gland
luminal cell of prostate epithelium
basal cell of prostate epithelium
keratocyte
myometrial cell
respiratory goblet cell
intermediate monocyte
CD141-positive myeloid dendritic cell
CD1c-positive myeloid dendritic cell
pancreatic stellate cell
adventitial cell
kidney epithelial cell
vein endothelial cell
fibroblast of cardiac tissue
retinal blood vessel endothelial cell
acinar cell of salivary gland
skeletal muscle satellite stem cell
paneth cell of colon
transit amplifying cell of colon
transit amplifying cell of small intestine
intestinal crypt stem cell of large intestine
cardiac endothelial cell
intestinal tuft cell
lung ciliated cell
large intestine goblet cell
enterocyte of epithelium of small intestine
paneth cell of epithelium of small intestine
endothelial cell of hepatic sinusoid
endothelial cell of artery
conjunctival epithelial cell
small intestine goblet cell
bladder urothelial cell
intestinal enteroendocrine cell
lung microvascular endothelial cell
fibroblast of breast
Mueller cell
native cell
PTPRH
LYPD8
HEPACAM2
KEL
SDC1
LYPD3
SDK2
PRND
VASN
PTPRS
CD1C
SIGLEC9
IL17RA
PLA2G7
MERTK
LILRB5
L1CAM
THBD
LIFR
CLEC1A
PODXL
ISLR2
ENG
PCDH12
FLT1
ROBO4
CLEC14A
TIE1
TEK
CDH5
SELE
ICAM4
feature_name

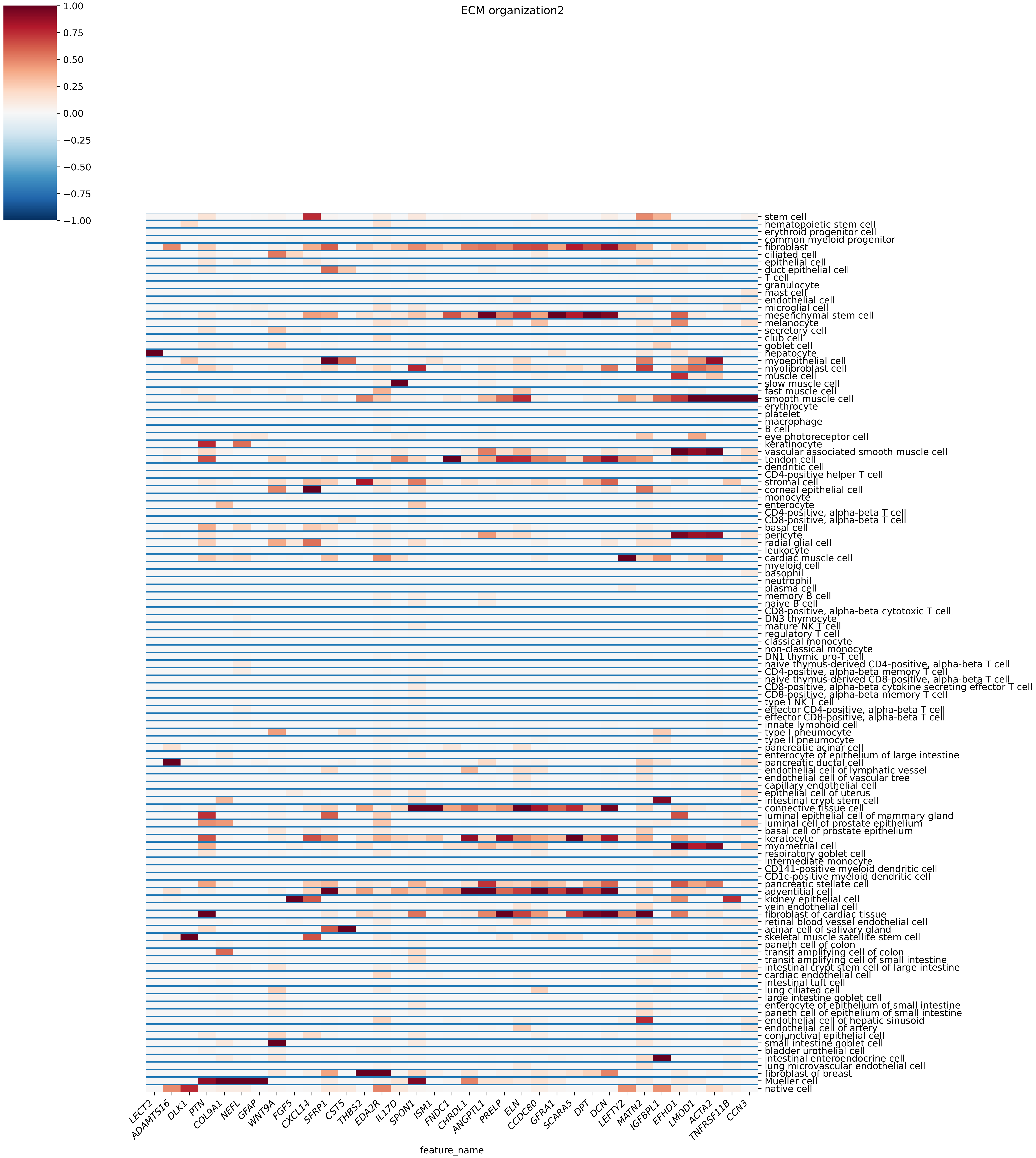
ECM organization2
1.00
0.75
0.50
0.25
0.00
−0.25
−0.50
−0.75
−1.00
stem cell
hematopoietic stem cell
erythroid progenitor cell
common myeloid progenitor
fibroblast
ciliated cell
epithelial cell
duct epithelial cell
T cell
granulocyte
mast cell
endothelial cell
microglial cell
mesenchymal stem cell
melanocyte
secretory cell
club cell
goblet cell
hepatocyte
myoepithelial cell
myofibroblast cell
muscle cell
slow muscle cell
fast muscle cell
smooth muscle cell
erythrocyte
platelet
macrophage
B cell
eye photoreceptor cell
keratinocyte
vascular associated smooth muscle cell
tendon cell
dendritic cell
CD4-positive helper T cell
stromal cell
corneal epithelial cell
monocyte
enterocyte
CD4-positive, alpha-beta T cell
CD8-positive, alpha-beta T cell
basal cell
pericyte
radial glial cell
leukocyte
cardiac muscle cell
myeloid cell
basophil
neutrophil
plasma cell
memory B cell
naive B cell
CD8-positive, alpha-beta cytotoxic T cell
DN3 thymocyte
mature NK T cell
regulatory T cell
classical monocyte
non-classical monocyte
DN1 thymic pro-T cell
naive thymus-derived CD4-positive, alpha-beta T cell
CD4-positive, alpha-beta memory T cell
naive thymus-derived CD8-positive, alpha-beta T cell
CD8-positive, alpha-beta cytokine secreting effector T cell
CD8-positive, alpha-beta memory T cell
type I NK T cell
effector CD4-positive, alpha-beta T cell
effector CD8-positive, alpha-beta T cell
innate lymphoid cell
type I pneumocyte
type II pneumocyte
pancreatic acinar cell
enterocyte of epithelium of large intestine
pancreatic ductal cell
endothelial cell of lymphatic vessel
endothelial cell of vascular tree
capillary endothelial cell
epithelial cell of uterus
intestinal crypt stem cell
connective tissue cell
luminal epithelial cell of mammary gland
luminal cell of prostate epithelium
basal cell of prostate epithelium
keratocyte
myometrial cell
respiratory goblet cell
intermediate monocyte
CD141-positive myeloid dendritic cell
CD1c-positive myeloid dendritic cell
pancreatic stellate cell
adventitial cell
kidney epithelial cell
vein endothelial cell
fibroblast of cardiac tissue
retinal blood vessel endothelial cell
acinar cell of salivary gland
skeletal muscle satellite stem cell
paneth cell of colon
transit amplifying cell of colon
transit amplifying cell of small intestine
intestinal crypt stem cell of large intestine
cardiac endothelial cell
intestinal tuft cell
lung ciliated cell
large intestine goblet cell
enterocyte of epithelium of small intestine
paneth cell of epithelium of small intestine
endothelial cell of hepatic sinusoid
endothelial cell of artery
conjunctival epithelial cell
small intestine goblet cell
bladder urothelial cell
intestinal enteroendocrine cell
lung microvascular endothelial cell
fibroblast of breast
Mueller cell
native cell
LECT2
ADAMTS16
DLK1
PTN
COL9A1
NEFL
GFAP
WNT9A
FGF5
CXCL14
SFRP1
CST5
THBS2
EDA2R
IL17D
SPON1
ISM1
FNDC1
CHRDL1
ANGPTL1
PRELP
ELN
CCDC80
GFRA1
SCARA5
DPT
DCN
LEFTY2
MATN2
IGFBPL1
EFHD1
LMOD1
ACTA2
TNFRSF11B
CCN3
feature_name

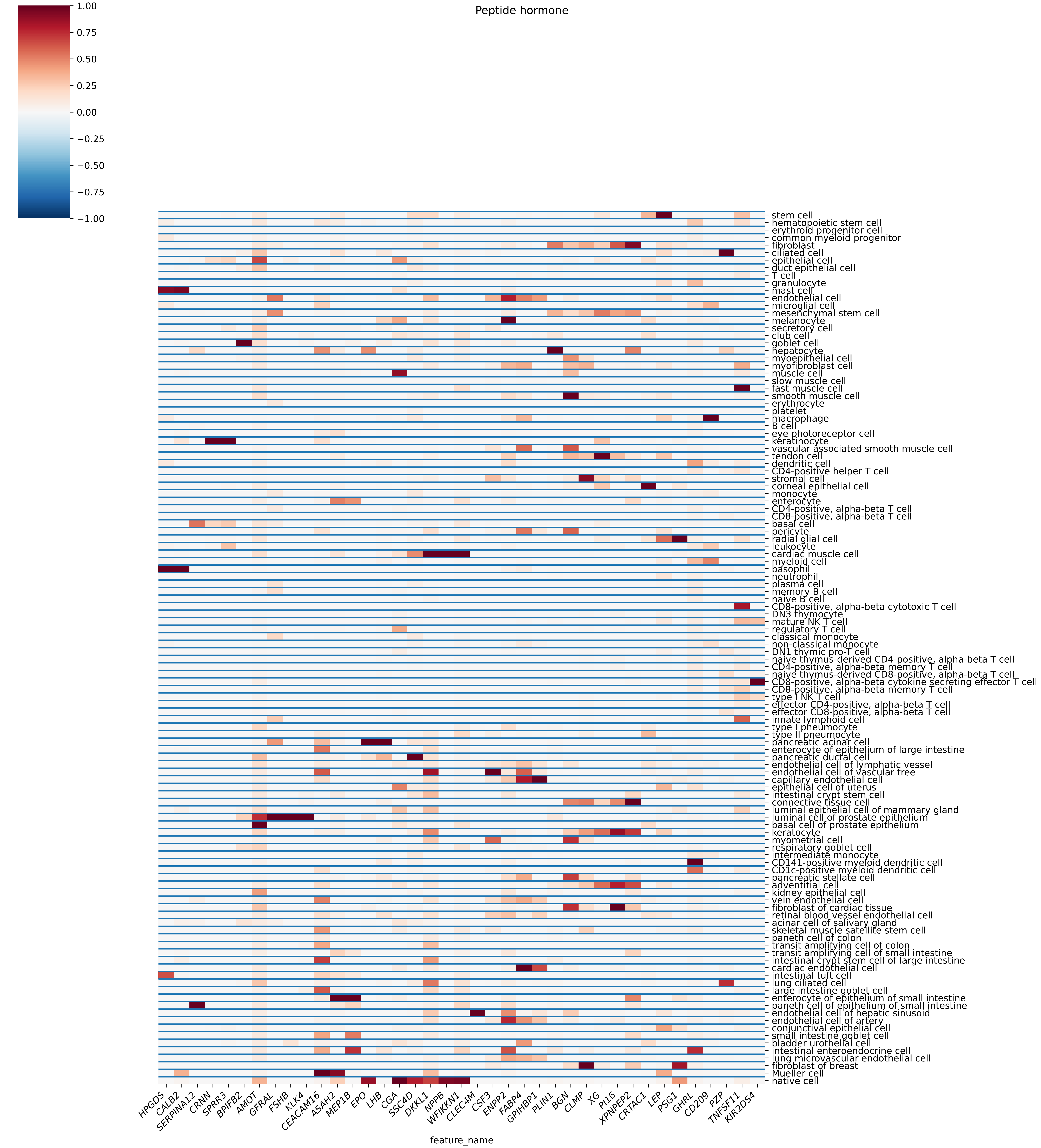

Peptide hormone
1.00
0.75
0.50
0.25
0.00
−0.25
−0.50
−0.75
−1.00
stem cell
hematopoietic stem cell
erythroid progenitor cell
common myeloid progenitor
fibroblast
ciliated cell
epithelial cell
duct epithelial cell
T cell
granulocyte
mast cell
endothelial cell
microglial cell
mesenchymal stem cell
melanocyte
secretory cell
club cell
goblet cell
hepatocyte
myoepithelial cell
myofibroblast cell
muscle cell
slow muscle cell
fast muscle cell
smooth muscle cell
erythrocyte
platelet
macrophage
B cell
eye photoreceptor cell
keratinocyte
vascular associated smooth muscle cell
tendon cell
dendritic cell
CD4-positive helper T cell
stromal cell
corneal epithelial cell
monocyte
enterocyte
CD4-positive, alpha-beta T cell
CD8-positive, alpha-beta T cell
basal cell
pericyte
radial glial cell
leukocyte
cardiac muscle cell
myeloid cell
basophil
neutrophil
plasma cell
memory B cell
naive B cell
CD8-positive, alpha-beta cytotoxic T cell
DN3 thymocyte
mature NK T cell
regulatory T cell
classical monocyte
non-classical monocyte
DN1 thymic pro-T cell
naive thymus-derived CD4-positive, alpha-beta T cell
CD4-positive, alpha-beta memory T cell
naive thymus-derived CD8-positive, alpha-beta T cell
CD8-positive, alpha-beta cytokine secreting effector T cell
CD8-positive, alpha-beta memory T cell
type I NK T cell
effector CD4-positive, alpha-beta T cell
effector CD8-positive, alpha-beta T cell
innate lymphoid cell
type I pneumocyte
type II pneumocyte
pancreatic acinar cell
enterocyte of epithelium of large intestine
pancreatic ductal cell
endothelial cell of lymphatic vessel
endothelial cell of vascular tree
capillary endothelial cell
epithelial cell of uterus
intestinal crypt stem cell
connective tissue cell
luminal epithelial cell of mammary gland
luminal cell of prostate epithelium
basal cell of prostate epithelium
keratocyte
myometrial cell
respiratory goblet cell
intermediate monocyte
CD141-positive myeloid dendritic cell
CD1c-positive myeloid dendritic cell
pancreatic stellate cell
adventitial cell
kidney epithelial cell
vein endothelial cell
fibroblast of cardiac tissue
retinal blood vessel endothelial cell
acinar cell of salivary gland
skeletal muscle satellite stem cell
paneth cell of colon
transit amplifying cell of colon
transit amplifying cell of small intestine
intestinal crypt stem cell of large intestine
cardiac endothelial cell
intestinal tuft cell
lung ciliated cell
large intestine goblet cell
enterocyte of epithelium of small intestine
paneth cell of epithelium of small intestine
endothelial cell of hepatic sinusoid
endothelial cell of artery
conjunctival epithelial cell
small intestine goblet cell
bladder urothelial cell
intestinal enteroendocrine cell
lung microvascular endothelial cell
fibroblast of breast
Mueller cell
native cell
HPGDS
CALB2
SERPINA12
CRNN
SPRR3
BPIFB2
AMOT
GFRAL
FSHB
KLK4
CEACAM16
ASAH2
MEP1B
EPO
LHB
CGA
SSC4D
DKKL1
NPPB
WFIKKN1
CLEC4M
CSF3
ENPP2
FABP4
GPIHBP1
PLIN1
BGN
CLMP
XG
PI16
XPNPEP2
CRTAC1
LEP
PSG1
GHRL
CD209
PZP
TNFSF11
KIR2DS4
feature_name

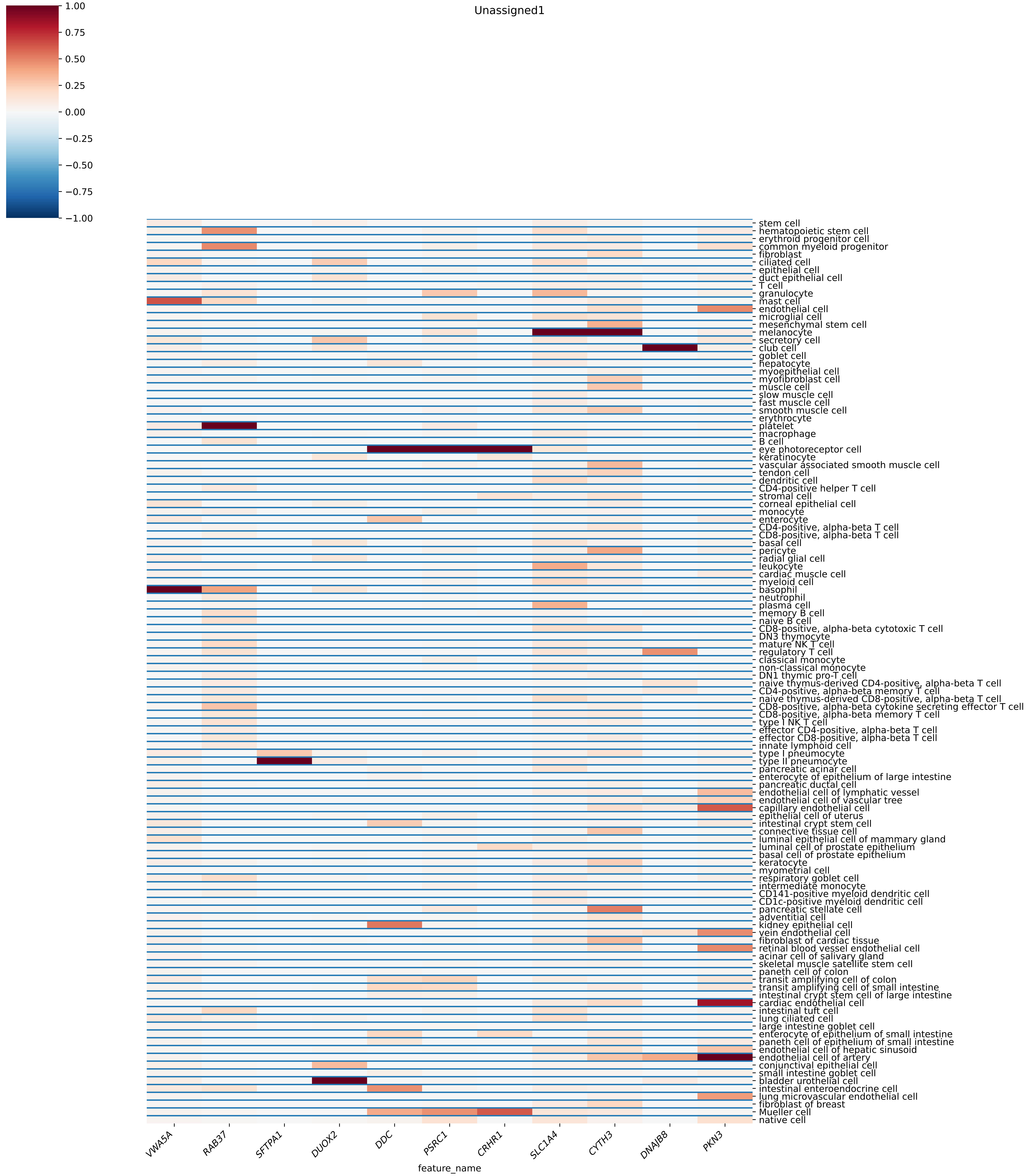

Unassigned1
1.00
0.75
0.50
0.25
0.00
−0.25
−0.50
−0.75
−1.00
stem cell
hematopoietic stem cell
erythroid progenitor cell
common myeloid progenitor
fibroblast
ciliated cell
epithelial cell
duct epithelial cell
T cell
granulocyte
mast cell
endothelial cell
microglial cell
mesenchymal stem cell
melanocyte
secretory cell
club cell
goblet cell
hepatocyte
myoepithelial cell
myofibroblast cell
muscle cell
slow muscle cell
fast muscle cell
smooth muscle cell
erythrocyte
platelet
macrophage
B cell
eye photoreceptor cell
keratinocyte
vascular associated smooth muscle cell
tendon cell
dendritic cell
CD4-positive helper T cell
stromal cell
corneal epithelial cell
monocyte
enterocyte
CD4-positive, alpha-beta T cell
CD8-positive, alpha-beta T cell
basal cell
pericyte
radial glial cell
leukocyte
cardiac muscle cell
myeloid cell
basophil
neutrophil
plasma cell
memory B cell
naive B cell
CD8-positive, alpha-beta cytotoxic T cell
DN3 thymocyte
mature NK T cell
regulatory T cell
classical monocyte
non-classical monocyte
DN1 thymic pro-T cell
naive thymus-derived CD4-positive, alpha-beta T cell
CD4-positive, alpha-beta memory T cell
naive thymus-derived CD8-positive, alpha-beta T cell
CD8-positive, alpha-beta cytokine secreting effector T cell
CD8-positive, alpha-beta memory T cell
type I NK T cell
effector CD4-positive, alpha-beta T cell
effector CD8-positive, alpha-beta T cell
innate lymphoid cell
type I pneumocyte
type II pneumocyte
pancreatic acinar cell
enterocyte of epithelium of large intestine
pancreatic ductal cell
endothelial cell of lymphatic vessel
endothelial cell of vascular tree
capillary endothelial cell
epithelial cell of uterus
intestinal crypt stem cell
connective tissue cell
luminal epithelial cell of mammary gland
luminal cell of prostate epithelium
basal cell of prostate epithelium
keratocyte
myometrial cell
respiratory goblet cell
intermediate monocyte
CD141-positive myeloid dendritic cell
CD1c-positive myeloid dendritic cell
pancreatic stellate cell
adventitial cell
kidney epithelial cell
vein endothelial cell
fibroblast of cardiac tissue
retinal blood vessel endothelial cell
acinar cell of salivary gland
skeletal muscle satellite stem cell
paneth cell of colon
transit amplifying cell of colon
transit amplifying cell of small intestine
intestinal crypt stem cell of large intestine
cardiac endothelial cell
intestinal tuft cell
lung ciliated cell
large intestine goblet cell
enterocyte of epithelium of small intestine
paneth cell of epithelium of small intestine
endothelial cell of hepatic sinusoid
endothelial cell of artery
conjunctival epithelial cell
small intestine goblet cell
bladder urothelial cell
intestinal enteroendocrine cell
lung microvascular endothelial cell
fibroblast of breast
Mueller cell
native cell
VWA5A
RAB37
SFTPA1
DUOX2
DDC
PSRC1
CRHR1
SLC1A4
CYTH3
DNAJB8
PKN3
feature_name

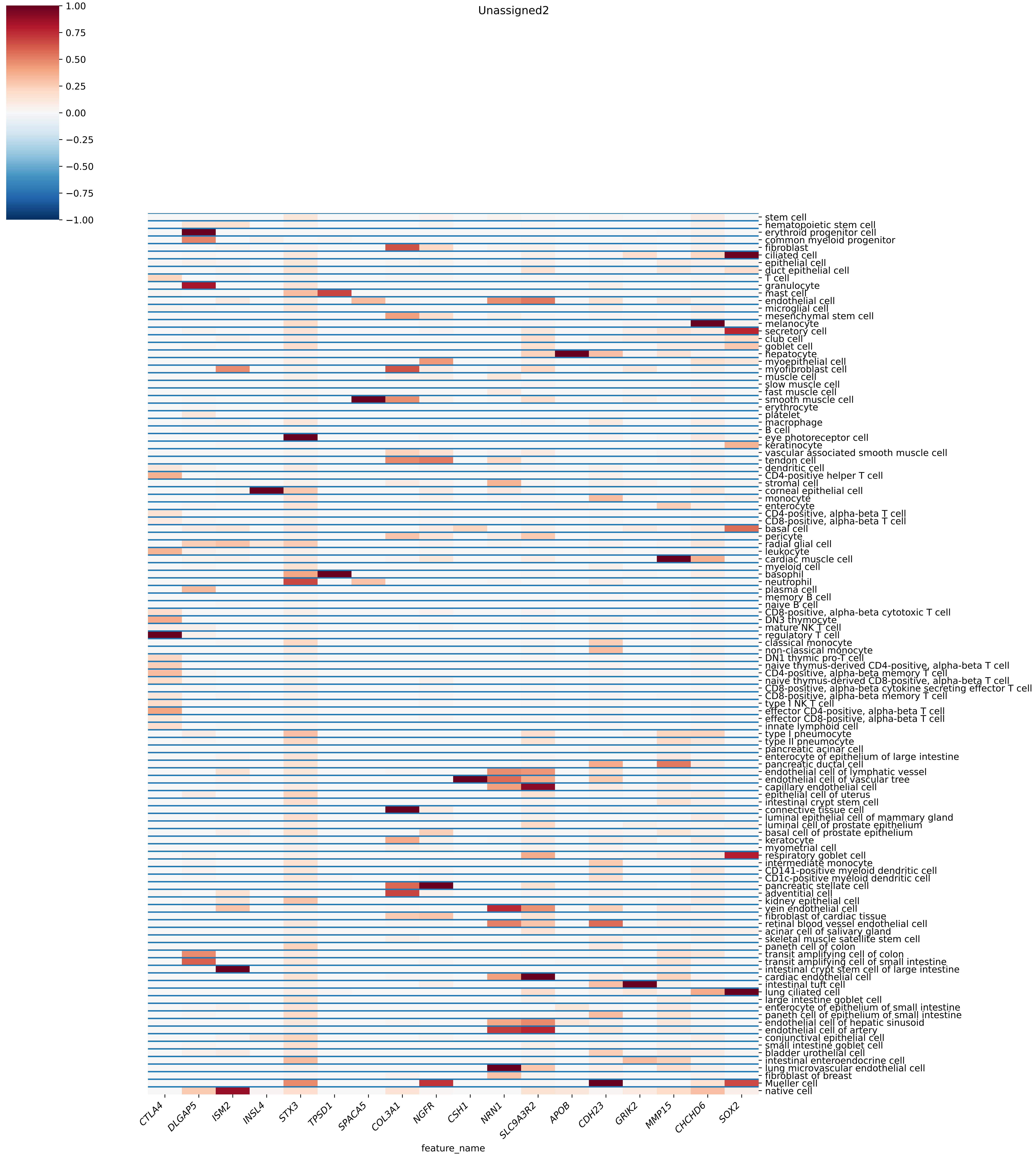
Unassigned2
1.00
0.75
0.50
0.25
0.00
−0.25
−0.50
−0.75
−1.00
stem cell
hematopoietic stem cell
erythroid progenitor cell
common myeloid progenitor
fibroblast
ciliated cell
epithelial cell
duct epithelial cell
T cell
granulocyte
mast cell
endothelial cell
microglial cell
mesenchymal stem cell
melanocyte
secretory cell
club cell
goblet cell
hepatocyte
myoepithelial cell
myofibroblast cell
muscle cell
slow muscle cell
fast muscle cell
smooth muscle cell
erythrocyte
platelet
macrophage
B cell
eye photoreceptor cell
keratinocyte
vascular associated smooth muscle cell
tendon cell
dendritic cell
CD4-positive helper T cell
stromal cell
corneal epithelial cell
monocyte
enterocyte
CD4-positive, alpha-beta T cell
CD8-positive, alpha-beta T cell
basal cell
pericyte
radial glial cell
leukocyte
cardiac muscle cell
myeloid cell
basophil
neutrophil
plasma cell
memory B cell
naive B cell
CD8-positive, alpha-beta cytotoxic T cell
DN3 thymocyte
mature NK T cell
regulatory T cell
classical monocyte
non-classical monocyte
DN1 thymic pro-T cell
naive thymus-derived CD4-positive, alpha-beta T cell
CD4-positive, alpha-beta memory T cell
naive thymus-derived CD8-positive, alpha-beta T cell
CD8-positive, alpha-beta cytokine secreting effector T cell
CD8-positive, alpha-beta memory T cell
type I NK T cell
effector CD4-positive, alpha-beta T cell
effector CD8-positive, alpha-beta T cell
innate lymphoid cell
type I pneumocyte
type II pneumocyte
pancreatic acinar cell
enterocyte of epithelium of large intestine
pancreatic ductal cell
endothelial cell of lymphatic vessel
endothelial cell of vascular tree
capillary endothelial cell
epithelial cell of uterus
intestinal crypt stem cell
connective tissue cell
luminal epithelial cell of mammary gland
luminal cell of prostate epithelium
basal cell of prostate epithelium
keratocyte
myometrial cell
respiratory goblet cell
intermediate monocyte
CD141-positive myeloid dendritic cell
CD1c-positive myeloid dendritic cell
pancreatic stellate cell
adventitial cell
kidney epithelial cell
vein endothelial cell
fibroblast of cardiac tissue
retinal blood vessel endothelial cell
acinar cell of salivary gland
skeletal muscle satellite stem cell
paneth cell of colon
transit amplifying cell of colon
transit amplifying cell of small intestine
intestinal crypt stem cell of large intestine
cardiac endothelial cell
intestinal tuft cell
lung ciliated cell
large intestine goblet cell
enterocyte of epithelium of small intestine
paneth cell of epithelium of small intestine
endothelial cell of hepatic sinusoid
endothelial cell of artery
conjunctival epithelial cell
small intestine goblet cell
bladder urothelial cell
intestinal enteroendocrine cell
lung microvascular endothelial cell
fibroblast of breast
Mueller cell
native cell
CTLA4
DLGAP5
ISM2
INSL4
STX3
TPSD1
SPACA5
COL3A1
NGFR
CSH1
NRN1
SLC9A3R2
APOB
CDH23
GRIK2
MMP15
CHCHD6
SOX2
feature_name

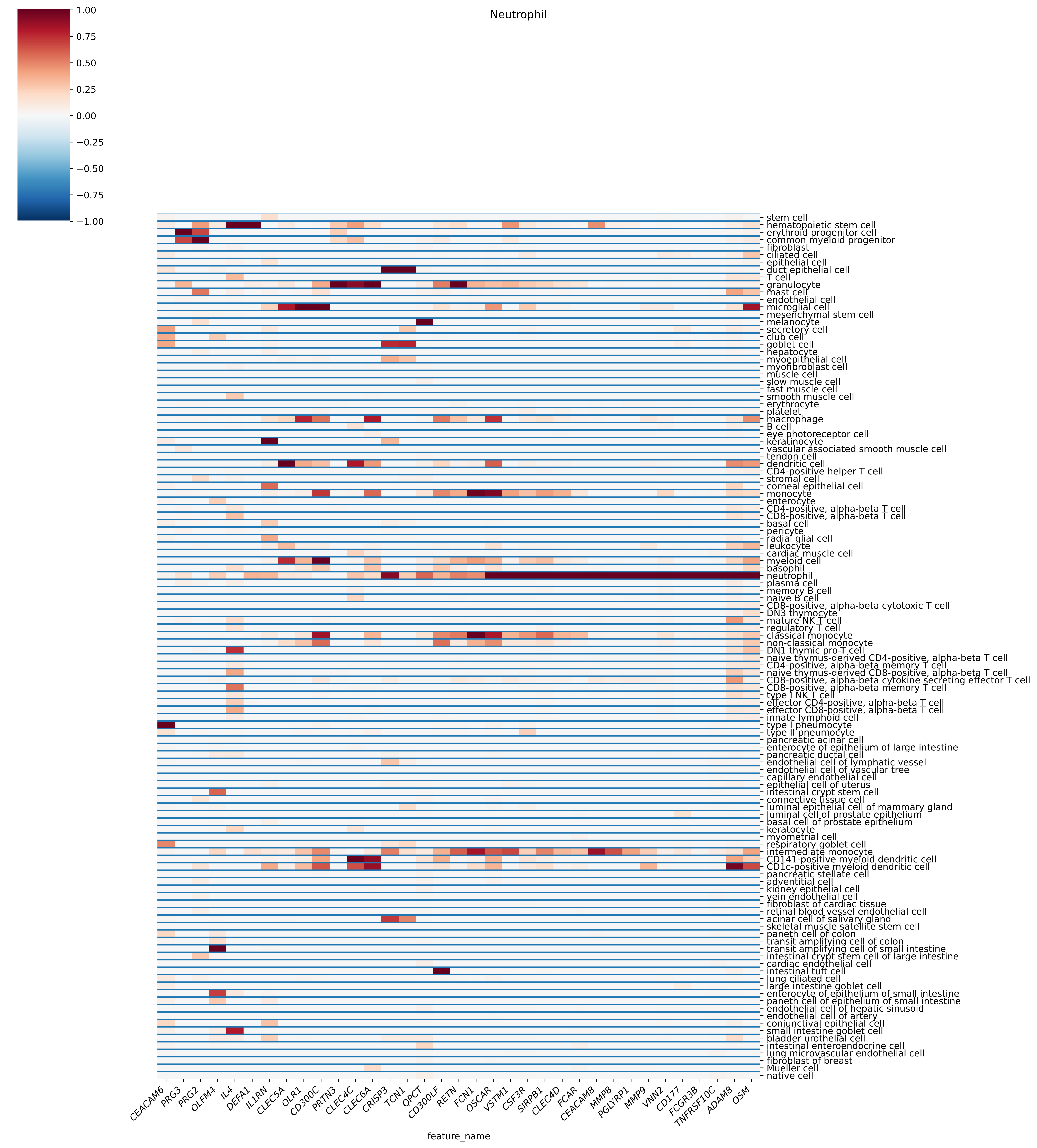
Neutrophil
1.00
0.75
0.50
0.25
0.00
−0.25
−0.50
−0.75
−1.00
stem cell
hematopoietic stem cell
erythroid progenitor cell
common myeloid progenitor
fibroblast
ciliated cell
epithelial cell
duct epithelial cell
T cell
granulocyte
mast cell
endothelial cell
microglial cell
mesenchymal stem cell
melanocyte
secretory cell
club cell
goblet cell
hepatocyte
myoepithelial cell
myofibroblast cell
muscle cell
slow muscle cell
fast muscle cell
smooth muscle cell
erythrocyte
platelet
macrophage
B cell
eye photoreceptor cell
keratinocyte
vascular associated smooth muscle cell
tendon cell
dendritic cell
CD4-positive helper T cell
stromal cell
corneal epithelial cell
monocyte
enterocyte
CD4-positive, alpha-beta T cell
CD8-positive, alpha-beta T cell
basal cell
pericyte
radial glial cell
leukocyte
cardiac muscle cell
myeloid cell
basophil
neutrophil
plasma cell
memory B cell
naive B cell
CD8-positive, alpha-beta cytotoxic T cell
DN3 thymocyte
mature NK T cell
regulatory T cell
classical monocyte
non-classical monocyte
DN1 thymic pro-T cell
naive thymus-derived CD4-positive, alpha-beta T cell
CD4-positive, alpha-beta memory T cell
naive thymus-derived CD8-positive, alpha-beta T cell
CD8-positive, alpha-beta cytokine secreting effector T cell
CD8-positive, alpha-beta memory T cell
type I NK T cell
effector CD4-positive, alpha-beta T cell
effector CD8-positive, alpha-beta T cell
innate lymphoid cell
type I pneumocyte
type II pneumocyte
pancreatic acinar cell
enterocyte of epithelium of large intestine
pancreatic ductal cell
endothelial cell of lymphatic vessel
endothelial cell of vascular tree
capillary endothelial cell
epithelial cell of uterus
intestinal crypt stem cell
connective tissue cell
luminal epithelial cell of mammary gland
luminal cell of prostate epithelium
basal cell of prostate epithelium
keratocyte
myometrial cell
respiratory goblet cell
intermediate monocyte
CD141-positive myeloid dendritic cell
CD1c-positive myeloid dendritic cell
pancreatic stellate cell
adventitial cell
kidney epithelial cell
vein endothelial cell
fibroblast of cardiac tissue
retinal blood vessel endothelial cell
acinar cell of salivary gland
skeletal muscle satellite stem cell
paneth cell of colon
transit amplifying cell of colon
transit amplifying cell of small intestine
intestinal crypt stem cell of large intestine
cardiac endothelial cell
intestinal tuft cell
lung ciliated cell
large intestine goblet cell
enterocyte of epithelium of small intestine
paneth cell of epithelium of small intestine
endothelial cell of hepatic sinusoid
endothelial cell of artery
conjunctival epithelial cell
small intestine goblet cell
bladder urothelial cell
intestinal enteroendocrine cell
lung microvascular endothelial cell
fibroblast of breast
Mueller cell
native cell
CEACAM6
PRG3
PRG2
OLFM4
IL4
DEFA1
IL1RN
CLEC5A
OLR1
CD300C
PRTN3
CLEC4C
CLEC6A
CRISP3
TCN1
QPCT
CD300LF
RETN
FCN1
OSCAR
VSTM1
CSF3R
SIRPB1
CLEC4D
FCAR
CEACAM8
MMP8
PGLYRP1
MMP9
VNN2
CD177
FCGR3B
TNFRSF10C
ADAM8
OSM
feature_name

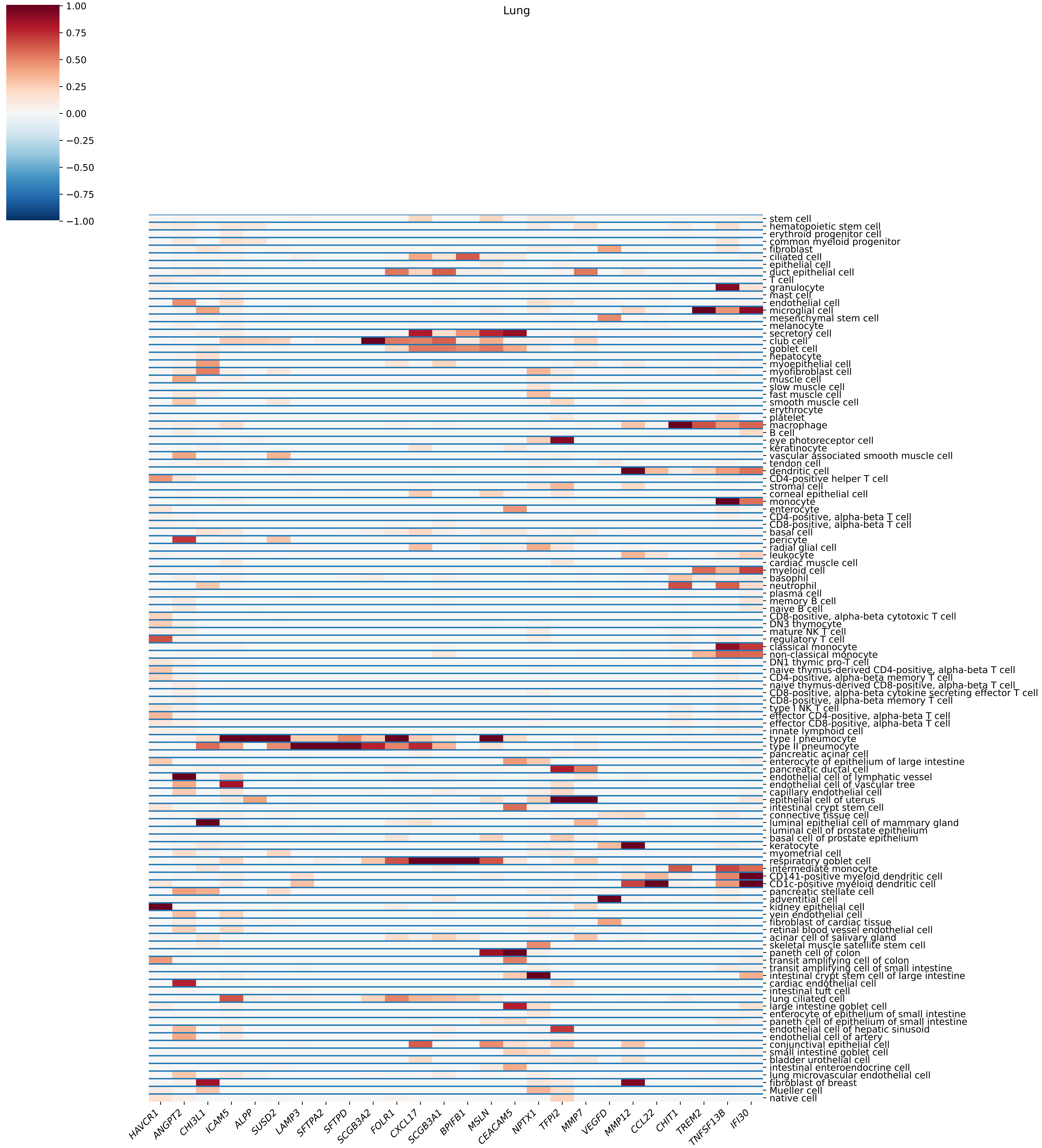

Lung
1.00
0.75
0.50
0.25
0.00
−0.25
−0.50
−0.75
−1.00
stem cell
hematopoietic stem cell
erythroid progenitor cell
common myeloid progenitor
fibroblast
ciliated cell
epithelial cell
duct epithelial cell
T cell
granulocyte
mast cell
endothelial cell
microglial cell
mesenchymal stem cell
melanocyte
secretory cell
club cell
goblet cell
hepatocyte
myoepithelial cell
myofibroblast cell
muscle cell
slow muscle cell
fast muscle cell
smooth muscle cell
erythrocyte
platelet
macrophage
B cell
eye photoreceptor cell
keratinocyte
vascular associated smooth muscle cell
tendon cell
dendritic cell
CD4-positive helper T cell
stromal cell
corneal epithelial cell
monocyte
enterocyte
CD4-positive, alpha-beta T cell
CD8-positive, alpha-beta T cell
basal cell
pericyte
radial glial cell
leukocyte
cardiac muscle cell
myeloid cell
basophil
neutrophil
plasma cell
memory B cell
naive B cell
CD8-positive, alpha-beta cytotoxic T cell
DN3 thymocyte
mature NK T cell
regulatory T cell
classical monocyte
non-classical monocyte
DN1 thymic pro-T cell
naive thymus-derived CD4-positive, alpha-beta T cell
CD4-positive, alpha-beta memory T cell
naive thymus-derived CD8-positive, alpha-beta T cell
CD8-positive, alpha-beta cytokine secreting effector T cell
CD8-positive, alpha-beta memory T cell
type I NK T cell
effector CD4-positive, alpha-beta T cell
effector CD8-positive, alpha-beta T cell
innate lymphoid cell
type I pneumocyte
type II pneumocyte
pancreatic acinar cell
enterocyte of epithelium of large intestine
pancreatic ductal cell
endothelial cell of lymphatic vessel
endothelial cell of vascular tree
capillary endothelial cell
epithelial cell of uterus
intestinal crypt stem cell
connective tissue cell
luminal epithelial cell of mammary gland
luminal cell of prostate epithelium
basal cell of prostate epithelium
keratocyte
myometrial cell
respiratory goblet cell
intermediate monocyte
CD141-positive myeloid dendritic cell
CD1c-positive myeloid dendritic cell
pancreatic stellate cell
adventitial cell
kidney epithelial cell
vein endothelial cell
fibroblast of cardiac tissue
retinal blood vessel endothelial cell
acinar cell of salivary gland
skeletal muscle satellite stem cell
paneth cell of colon
transit amplifying cell of colon
transit amplifying cell of small intestine
intestinal crypt stem cell of large intestine
cardiac endothelial cell
intestinal tuft cell
lung ciliated cell
large intestine goblet cell
enterocyte of epithelium of small intestine
paneth cell of epithelium of small intestine
endothelial cell of hepatic sinusoid
endothelial cell of artery
conjunctival epithelial cell
small intestine goblet cell
bladder urothelial cell
intestinal enteroendocrine cell
lung microvascular endothelial cell
fibroblast of breast
Mueller cell
native cell
HAVCR1
ANGPT2
CHI3L1
ICAM5
ALPP
SUSD2
LAMP3
SFTPA2
SFTPD
SCGB3A2
FOLR1
CXCL17
SCGB3A1
BPIFB1
MSLN
CEACAM5
NPTX1
TFPI2
MMP7
VEGFD
MMP12
CCL22
CHIT1
TREM2
TNFSF13B
IFI30
feature_name

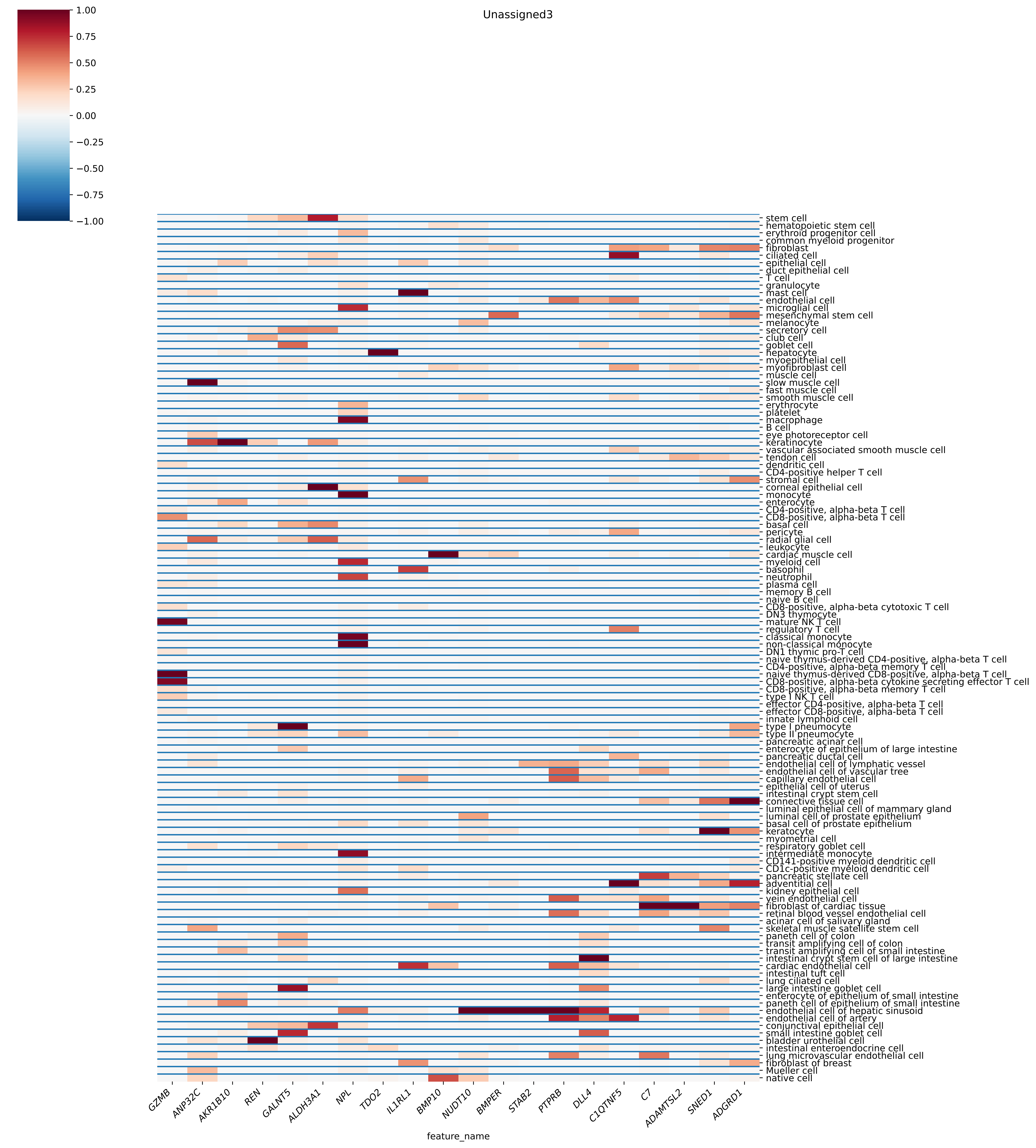

Unassigned3
1.00
0.75
0.50
0.25
0.00
−0.25
−0.50
−0.75
−1.00
stem cell
hematopoietic stem cell
erythroid progenitor cell
common myeloid progenitor
fibroblast
ciliated cell
epithelial cell
duct epithelial cell
T cell
granulocyte
mast cell
endothelial cell
microglial cell
mesenchymal stem cell
melanocyte
secretory cell
club cell
goblet cell
hepatocyte
myoepithelial cell
myofibroblast cell
muscle cell
slow muscle cell
fast muscle cell
smooth muscle cell
erythrocyte
platelet
macrophage
B cell
eye photoreceptor cell
keratinocyte
vascular associated smooth muscle cell
tendon cell
dendritic cell
CD4-positive helper T cell
stromal cell
corneal epithelial cell
monocyte
enterocyte
CD4-positive, alpha-beta T cell
CD8-positive, alpha-beta T cell
basal cell
pericyte
radial glial cell
leukocyte
cardiac muscle cell
myeloid cell
basophil
neutrophil
plasma cell
memory B cell
naive B cell
CD8-positive, alpha-beta cytotoxic T cell
DN3 thymocyte
mature NK T cell
regulatory T cell
classical monocyte
non-classical monocyte
DN1 thymic pro-T cell
naive thymus-derived CD4-positive, alpha-beta T cell
CD4-positive, alpha-beta memory T cell
naive thymus-derived CD8-positive, alpha-beta T cell
CD8-positive, alpha-beta cytokine secreting effector T cell
CD8-positive, alpha-beta memory T cell
type I NK T cell
effector CD4-positive, alpha-beta T cell
effector CD8-positive, alpha-beta T cell
innate lymphoid cell
type I pneumocyte
type II pneumocyte
pancreatic acinar cell
enterocyte of epithelium of large intestine
pancreatic ductal cell
endothelial cell of lymphatic vessel
endothelial cell of vascular tree
capillary endothelial cell
epithelial cell of uterus
intestinal crypt stem cell
connective tissue cell
luminal epithelial cell of mammary gland
luminal cell of prostate epithelium
basal cell of prostate epithelium
keratocyte
myometrial cell
respiratory goblet cell
intermediate monocyte
CD141-positive myeloid dendritic cell
CD1c-positive myeloid dendritic cell
pancreatic stellate cell
adventitial cell
kidney epithelial cell
vein endothelial cell
fibroblast of cardiac tissue
retinal blood vessel endothelial cell
acinar cell of salivary gland
skeletal muscle satellite stem cell
paneth cell of colon
transit amplifying cell of colon
transit amplifying cell of small intestine
intestinal crypt stem cell of large intestine
cardiac endothelial cell
intestinal tuft cell
lung ciliated cell
large intestine goblet cell
enterocyte of epithelium of small intestine
paneth cell of epithelium of small intestine
endothelial cell of hepatic sinusoid
endothelial cell of artery
conjunctival epithelial cell
small intestine goblet cell
bladder urothelial cell
intestinal enteroendocrine cell
lung microvascular endothelial cell
fibroblast of breast
Mueller cell
native cell
GZMB
ANP32C
AKR1B10
REN
GALNT5
ALDH3A1
NPL
TDO2
IL1RL1
BMP10
NUDT10
BMPER
STAB2
PTPRB
DLL4
C1QTNF5
C7
ADAMTSL2
SNED1
ADGRD1
feature_name

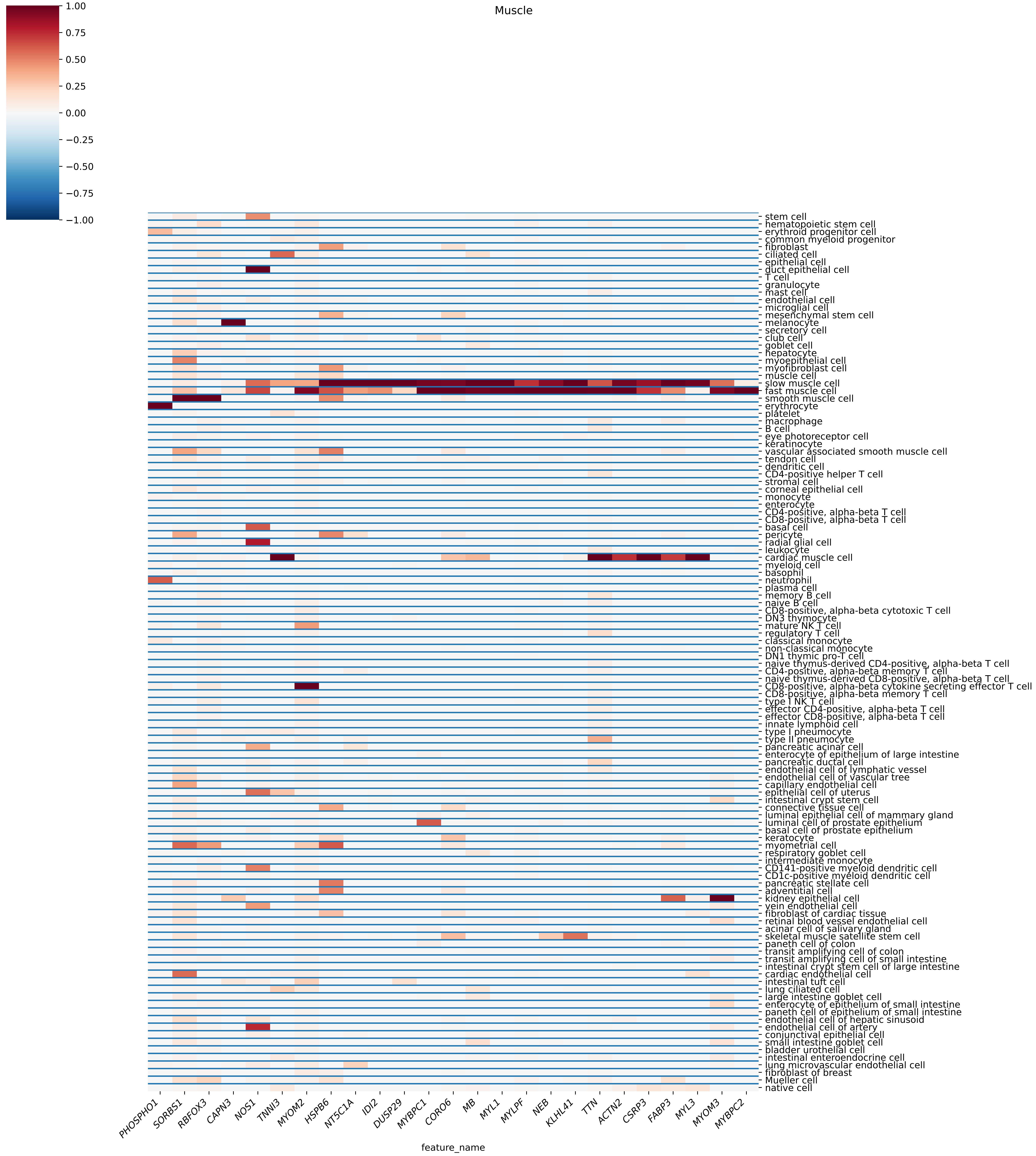
Muscle
1.00
0.75
0.50
0.25
0.00
−0.25
−0.50
−0.75
−1.00
stem cell
hematopoietic stem cell
erythroid progenitor cell
common myeloid progenitor
fibroblast
ciliated cell
epithelial cell
duct epithelial cell
T cell
granulocyte
mast cell
endothelial cell
microglial cell
mesenchymal stem cell
melanocyte
secretory cell
club cell
goblet cell
hepatocyte
myoepithelial cell
myofibroblast cell
muscle cell
slow muscle cell
fast muscle cell
smooth muscle cell
erythrocyte
platelet
macrophage
B cell
eye photoreceptor cell
keratinocyte
vascular associated smooth muscle cell
tendon cell
dendritic cell
CD4-positive helper T cell
stromal cell
corneal epithelial cell
monocyte
enterocyte
CD4-positive, alpha-beta T cell
CD8-positive, alpha-beta T cell
basal cell
pericyte
radial glial cell
leukocyte
cardiac muscle cell
myeloid cell
basophil
neutrophil
plasma cell
memory B cell
naive B cell
CD8-positive, alpha-beta cytotoxic T cell
DN3 thymocyte
mature NK T cell
regulatory T cell
classical monocyte
non-classical monocyte
DN1 thymic pro-T cell
naive thymus-derived CD4-positive, alpha-beta T cell
CD4-positive, alpha-beta memory T cell
naive thymus-derived CD8-positive, alpha-beta T cell
CD8-positive, alpha-beta cytokine secreting effector T cell
CD8-positive, alpha-beta memory T cell
type I NK T cell
effector CD4-positive, alpha-beta T cell
effector CD8-positive, alpha-beta T cell
innate lymphoid cell
type I pneumocyte
type II pneumocyte
pancreatic acinar cell
enterocyte of epithelium of large intestine
pancreatic ductal cell
endothelial cell of lymphatic vessel
endothelial cell of vascular tree
capillary endothelial cell
epithelial cell of uterus
intestinal crypt stem cell
connective tissue cell
luminal epithelial cell of mammary gland
luminal cell of prostate epithelium
basal cell of prostate epithelium
keratocyte
myometrial cell
respiratory goblet cell
intermediate monocyte
CD141-positive myeloid dendritic cell
CD1c-positive myeloid dendritic cell
pancreatic stellate cell
adventitial cell
kidney epithelial cell
vein endothelial cell
fibroblast of cardiac tissue
retinal blood vessel endothelial cell
acinar cell of salivary gland
skeletal muscle satellite stem cell
paneth cell of colon
transit amplifying cell of colon
transit amplifying cell of small intestine
intestinal crypt stem cell of large intestine
cardiac endothelial cell
intestinal tuft cell
lung ciliated cell
large intestine goblet cell
enterocyte of epithelium of small intestine
paneth cell of epithelium of small intestine
endothelial cell of hepatic sinusoid
endothelial cell of artery
conjunctival epithelial cell
small intestine goblet cell
bladder urothelial cell
intestinal enteroendocrine cell
lung microvascular endothelial cell
fibroblast of breast
Mueller cell
native cell
PHOSPHO1
SORBS1
RBFOX3
CAPN3
NOS1
TNNI3
MYOM2
HSPB6
NT5C1A
IDI2
DUSP29
MYBPC1
CORO6
MB
MYL1
MYLPF
NEB
KLHL41
TTN
ACTN2
CSRP3
FABP3
MYL3
MYOM3
MYBPC2
feature_name

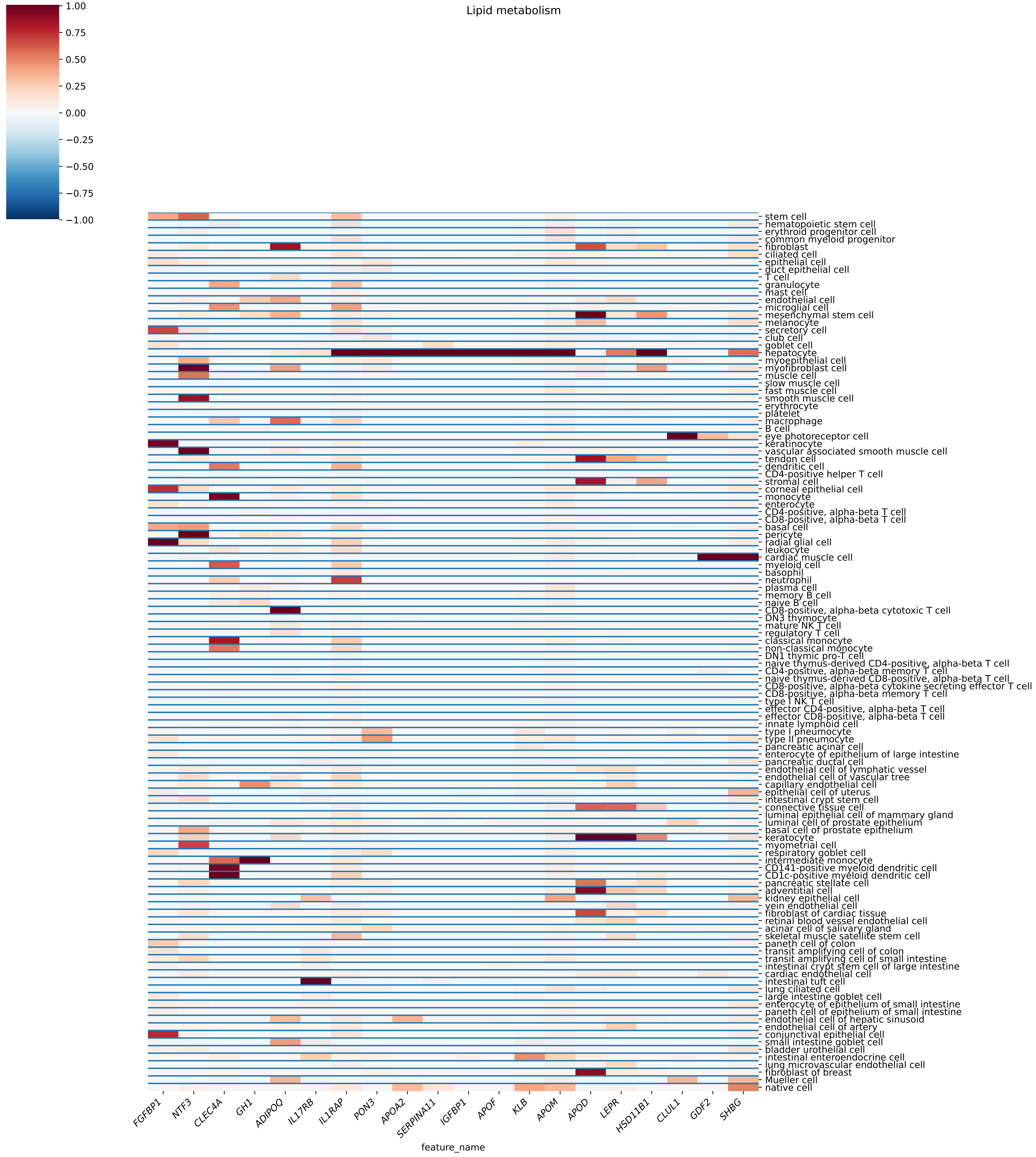
Lipid metabolism
1.00
0.75
0.50
0.25
0.00
−0.25
−0.50
−0.75
−1.00
stem cell
hematopoietic stem cell
erythroid progenitor cell
common myeloid progenitor
fibroblast
ciliated cell
epithelial cell
duct epithelial cell
T cell
granulocyte
mast cell
endothelial cell
microglial cell
mesenchymal stem cell
melanocyte
secretory cell
club cell
goblet cell
hepatocyte
myoepithelial cell
myofibroblast cell
muscle cell
slow muscle cell
fast muscle cell
smooth muscle cell
erythrocyte
platelet
macrophage
B cell
eye photoreceptor cell
keratinocyte
vascular associated smooth muscle cell
tendon cell
dendritic cell
CD4-positive helper T cell
stromal cell
corneal epithelial cell
monocyte
enterocyte
CD4-positive, alpha-beta T cell
CD8-positive, alpha-beta T cell
basal cell
pericyte
radial glial cell
leukocyte
cardiac muscle cell
myeloid cell
basophil
neutrophil
plasma cell
memory B cell
naive B cell
CD8-positive, alpha-beta cytotoxic T cell
DN3 thymocyte
mature NK T cell
regulatory T cell
classical monocyte
non-classical monocyte
DN1 thymic pro-T cell
naive thymus-derived CD4-positive, alpha-beta T cell
CD4-positive, alpha-beta memory T cell
naive thymus-derived CD8-positive, alpha-beta T cell
CD8-positive, alpha-beta cytokine secreting effector T cell
CD8-positive, alpha-beta memory T cell
type I NK T cell
effector CD4-positive, alpha-beta T cell
effector CD8-positive, alpha-beta T cell
innate lymphoid cell
type I pneumocyte
type II pneumocyte
pancreatic acinar cell
enterocyte of epithelium of large intestine
pancreatic ductal cell
endothelial cell of lymphatic vessel
endothelial cell of vascular tree
capillary endothelial cell
epithelial cell of uterus
intestinal crypt stem cell
connective tissue cell
luminal epithelial cell of mammary gland
luminal cell of prostate epithelium
basal cell of prostate epithelium
keratocyte
myometrial cell
respiratory goblet cell
intermediate monocyte
CD141-positive myeloid dendritic cell
CD1c-positive myeloid dendritic cell
pancreatic stellate cell
adventitial cell
kidney epithelial cell
vein endothelial cell
fibroblast of cardiac tissue
retinal blood vessel endothelial cell
acinar cell of salivary gland
skeletal muscle satellite stem cell
paneth cell of colon
transit amplifying cell of colon
transit amplifying cell of small intestine
intestinal crypt stem cell of large intestine
cardiac endothelial cell
intestinal tuft cell
lung ciliated cell
large intestine goblet cell
enterocyte of epithelium of small intestine
paneth cell of epithelium of small intestine
endothelial cell of hepatic sinusoid
endothelial cell of artery
conjunctival epithelial cell
small intestine goblet cell
bladder urothelial cell
intestinal enteroendocrine cell
lung microvascular endothelial cell
fibroblast of breast
Mueller cell
native cell
FGFBP1
NTF3
CLEC4A
GH1
ADIPOQ
IL17RB
IL1RAP
PON3
APOA2
SERPINA11
IGFBP1
APOF
KLB
APOM
APOD
LEPR
HSD11B1
CLUL1
GDF2
SHBG
feature_name

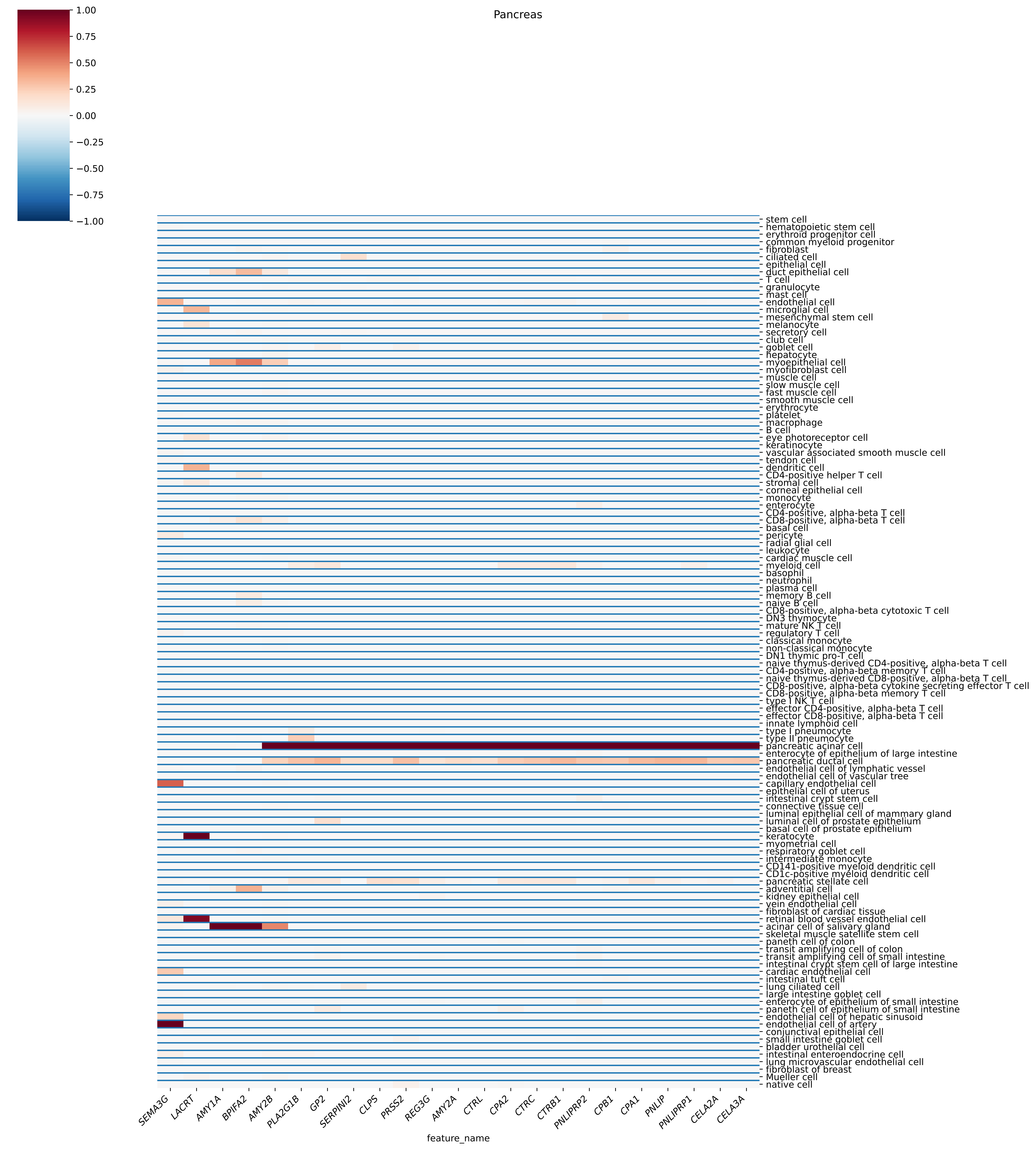

Pancreas
1.00
0.75
0.50
0.25
0.00
−0.25
−0.50
−0.75
−1.00
stem cell
hematopoietic stem cell
erythroid progenitor cell
common myeloid progenitor
fibroblast
ciliated cell
epithelial cell
duct epithelial cell
T cell
granulocyte
mast cell
endothelial cell
microglial cell
mesenchymal stem cell
melanocyte
secretory cell
club cell
goblet cell
hepatocyte
myoepithelial cell
myofibroblast cell
muscle cell
slow muscle cell
fast muscle cell
smooth muscle cell
erythrocyte
platelet
macrophage
B cell
eye photoreceptor cell
keratinocyte
vascular associated smooth muscle cell
tendon cell
dendritic cell
CD4-positive helper T cell
stromal cell
corneal epithelial cell
monocyte
enterocyte
CD4-positive, alpha-beta T cell
CD8-positive, alpha-beta T cell
basal cell
pericyte
radial glial cell
leukocyte
cardiac muscle cell
myeloid cell
basophil
neutrophil
plasma cell
memory B cell
naive B cell
CD8-positive, alpha-beta cytotoxic T cell
DN3 thymocyte
mature NK T cell
regulatory T cell
classical monocyte
non-classical monocyte
DN1 thymic pro-T cell
naive thymus-derived CD4-positive, alpha-beta T cell
CD4-positive, alpha-beta memory T cell
naive thymus-derived CD8-positive, alpha-beta T cell
CD8-positive, alpha-beta cytokine secreting effector T cell
CD8-positive, alpha-beta memory T cell
type I NK T cell
effector CD4-positive, alpha-beta T cell
effector CD8-positive, alpha-beta T cell
innate lymphoid cell
type I pneumocyte
type II pneumocyte
pancreatic acinar cell
enterocyte of epithelium of large intestine
pancreatic ductal cell
endothelial cell of lymphatic vessel
endothelial cell of vascular tree
capillary endothelial cell
epithelial cell of uterus
intestinal crypt stem cell
connective tissue cell
luminal epithelial cell of mammary gland
luminal cell of prostate epithelium
basal cell of prostate epithelium
keratocyte
myometrial cell
respiratory goblet cell
intermediate monocyte
CD141-positive myeloid dendritic cell
CD1c-positive myeloid dendritic cell
pancreatic stellate cell
adventitial cell
kidney epithelial cell
vein endothelial cell
fibroblast of cardiac tissue
retinal blood vessel endothelial cell
acinar cell of salivary gland
skeletal muscle satellite stem cell
paneth cell of colon
transit amplifying cell of colon
transit amplifying cell of small intestine
intestinal crypt stem cell of large intestine
cardiac endothelial cell
intestinal tuft cell
lung ciliated cell
large intestine goblet cell
enterocyte of epithelium of small intestine
paneth cell of epithelium of small intestine
endothelial cell of hepatic sinusoid
endothelial cell of artery
conjunctival epithelial cell
small intestine goblet cell
bladder urothelial cell
intestinal enteroendocrine cell
lung microvascular endothelial cell
fibroblast of breast
Mueller cell
native cell
SEMA3G
LACRT
AMY1A
BPIFA2
AMY2B
PLA2G1B
GP2
SERPINI2
CLPS
PRSS2
REG3G
AMY2A
CTRL
CPA2
CTRC
CTRB1
PNLIPRP2
CPB1
CPA1
PNLIP
PNLIPRP1
CELA2A
CELA3A
feature_name

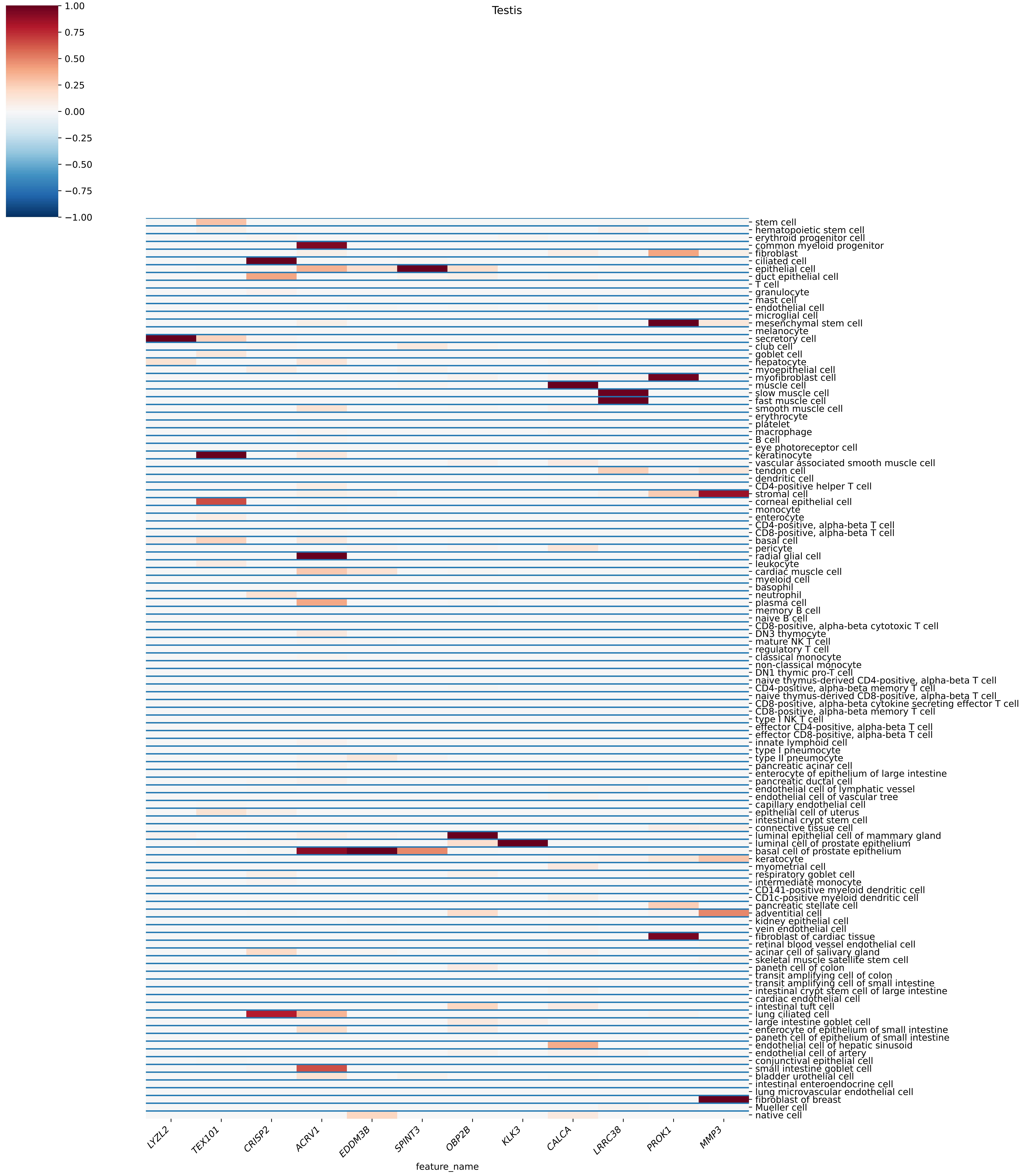

Testis
1.00
0.75
0.50
0.25
0.00
−0.25
−0.50
−0.75
−1.00
stem cell
hematopoietic stem cell
erythroid progenitor cell
common myeloid progenitor
fibroblast
ciliated cell
epithelial cell
duct epithelial cell
T cell
granulocyte
mast cell
endothelial cell
microglial cell
mesenchymal stem cell
melanocyte
secretory cell
club cell
goblet cell
hepatocyte
myoepithelial cell
myofibroblast cell
muscle cell
slow muscle cell
fast muscle cell
smooth muscle cell
erythrocyte
platelet
macrophage
B cell
eye photoreceptor cell
keratinocyte
vascular associated smooth muscle cell
tendon cell
dendritic cell
CD4-positive helper T cell
stromal cell
corneal epithelial cell
monocyte
enterocyte
CD4-positive, alpha-beta T cell
CD8-positive, alpha-beta T cell
basal cell
pericyte
radial glial cell
leukocyte
cardiac muscle cell
myeloid cell
basophil
neutrophil
plasma cell
memory B cell
naive B cell
CD8-positive, alpha-beta cytotoxic T cell
DN3 thymocyte
mature NK T cell
regulatory T cell
classical monocyte
non-classical monocyte
DN1 thymic pro-T cell
naive thymus-derived CD4-positive, alpha-beta T cell
CD4-positive, alpha-beta memory T cell
naive thymus-derived CD8-positive, alpha-beta T cell
CD8-positive, alpha-beta cytokine secreting effector T cell
CD8-positive, alpha-beta memory T cell
type I NK T cell
effector CD4-positive, alpha-beta T cell
effector CD8-positive, alpha-beta T cell
innate lymphoid cell
type I pneumocyte
type II pneumocyte
pancreatic acinar cell
enterocyte of epithelium of large intestine
pancreatic ductal cell
endothelial cell of lymphatic vessel
endothelial cell of vascular tree
capillary endothelial cell
epithelial cell of uterus
intestinal crypt stem cell
connective tissue cell
luminal epithelial cell of mammary gland
luminal cell of prostate epithelium
basal cell of prostate epithelium
keratocyte
myometrial cell
respiratory goblet cell
intermediate monocyte
CD141-positive myeloid dendritic cell
CD1c-positive myeloid dendritic cell
pancreatic stellate cell
adventitial cell
kidney epithelial cell
vein endothelial cell
fibroblast of cardiac tissue
retinal blood vessel endothelial cell
acinar cell of salivary gland
skeletal muscle satellite stem cell
paneth cell of colon
transit amplifying cell of colon
transit amplifying cell of small intestine
intestinal crypt stem cell of large intestine
cardiac endothelial cell
intestinal tuft cell
lung ciliated cell
large intestine goblet cell
enterocyte of epithelium of small intestine
paneth cell of epithelium of small intestine
endothelial cell of hepatic sinusoid
endothelial cell of artery
conjunctival epithelial cell
small intestine goblet cell
bladder urothelial cell
intestinal enteroendocrine cell
lung microvascular endothelial cell
fibroblast of breast
Mueller cell
native cell
LYZL2
TEX101
CRISP2
ACRV1
EDDM3B
SPINT3
OBP2B
KLK3
CALCA
LRRC38
PROK1
MMP3
feature_name
