## Supplementary material for "Representation learning based on proteomic profiles uncovers key cell types and biological processes contributing to the plasma proteome": Supplementary material.docx

**Supplementary Materials:**

**Figure S1.** Expression enrichment of PPP proteins across 54 human organs/tissues from GTEx. Each page corresponds to one protein module. For each organ/tissue, median Transcripts Per Million (TPM) values across samples are used (bulk-gex_v8_rna-seq_GTEx_Analysis_2017-06-05_v8_RNASeQCv1.1.9_gene_median_tpm.gct.gz). Each column (gene) is normalized by being divided by its maximum value. Genes with TPMs <= 10 in all organ/tissues are excluded. Genes that are expressed with TPMs exceeding 20% of the maximum in more than 10 organ/tissues are excluded. Protein modules with fewer than 8 genes left after filtering are not shown.

**Figure S2.** Expression enrichment of PPP proteins across 116 human cell types from Tabular Sapiens that have more than 100 UMIs. Processed Tabular Sapiens H5AD file is downloaded from cellxgene. Each page corresponds to one protein module. For each organ/tissue, mean Counts Per Million (CPM) values across cells are used. Each column (gene) is normalized by being divided by its maximum value. Genes with CPMs <= 0.001 in all organ/tissues are excluded. Genes that are expressed with TPMs exceeding 20% of the maximum in more than 15 cell types are excluded. Protein modules with fewer than 8 genes left after filtering are not shown.

**Figure S3.** Heatmap depicting correlations between 1^st^ Principal Component of each protein module and demographic information and blood test results.

**Figure S4.** Volcano plot showing results of module-variant associations. X-axis represents the mean differences in normalized effect size while y-axis represents -log10 of false discovery rate from the z-test. To make the plot legible we only included the associations with FDR < 1e-10. LD clumping within 500kb window was carried out before plotting, therefore each dot represents one locus.

**Table S1.** Module membership of each of the 2,935 plasma proteins.

**Table S2.** Pathway enrichment analysis results for inferred protein modules.

**Table S3.** Significant overlaps between protein modules and proteins dysregulated in diseases.

**Table S4.** Significant associations between protein modules and genetic variants.
