## Supplementary figures and images for "Representation learning based on proteomic profiles uncovers key cell types and biological processes contributing to the plasma proteome"

### FigureS3.pdf

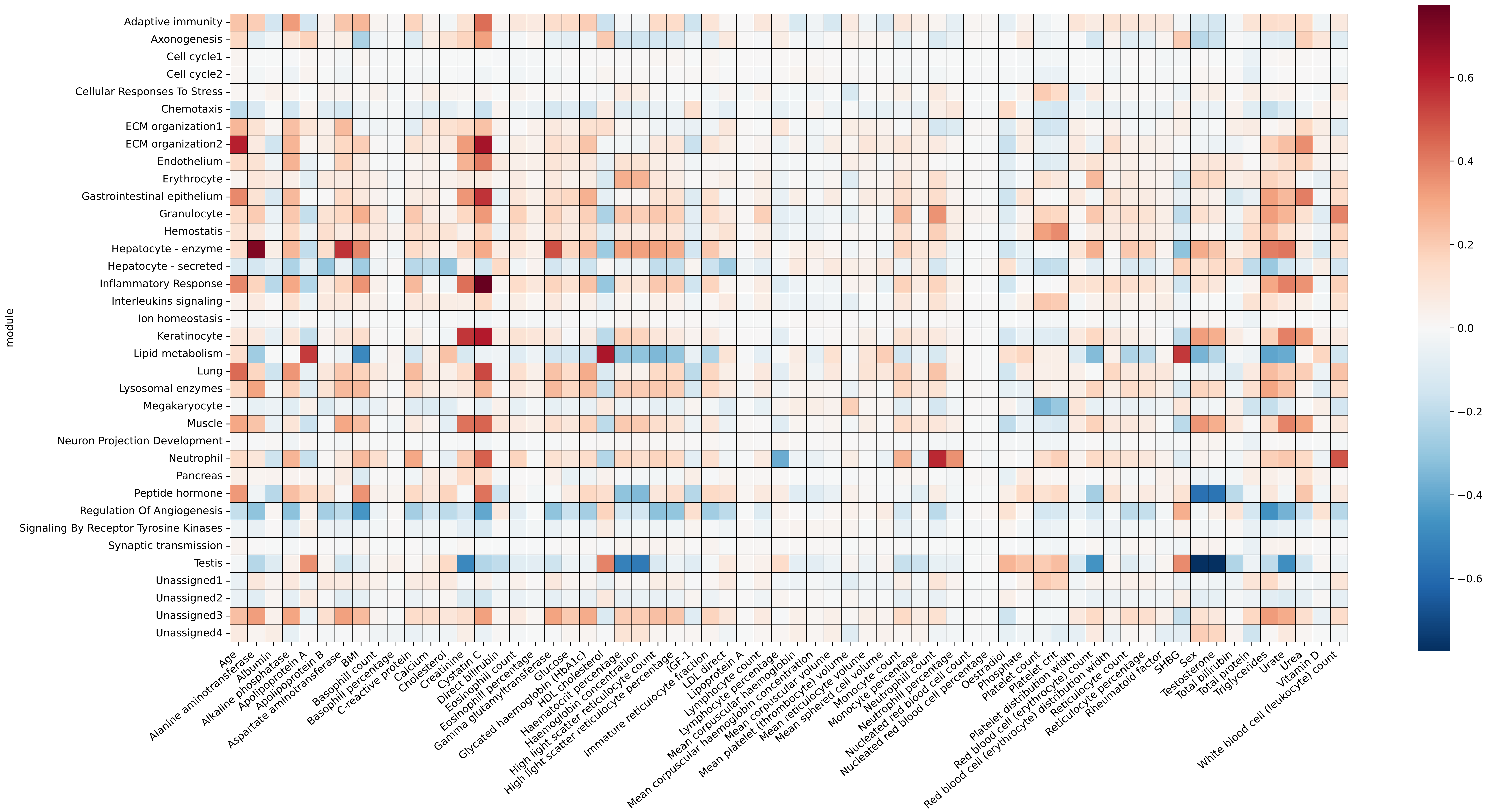

### FigureS4.png

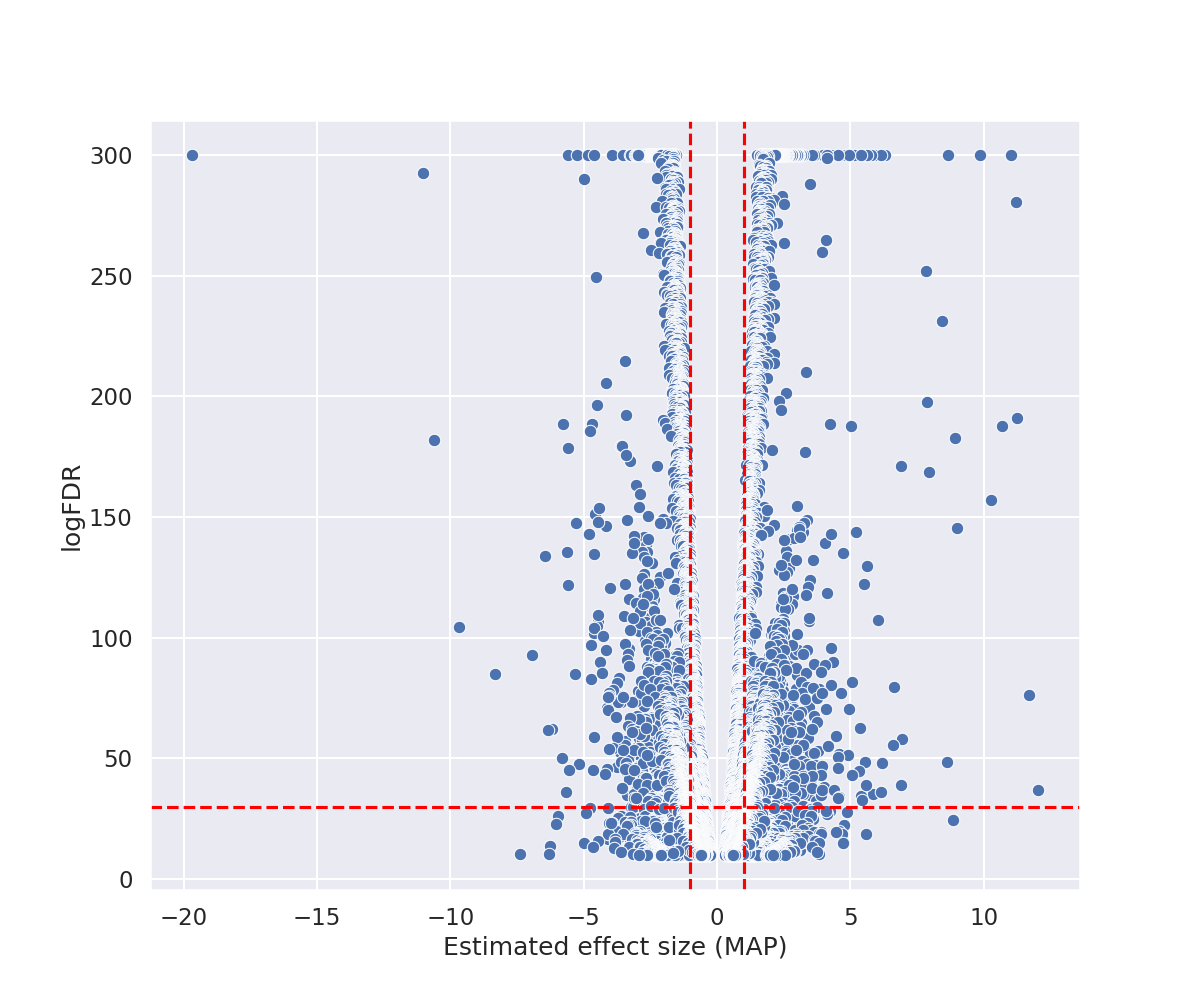
